# A spatially resolved genomic and molecular atlas of human white matter

**DOI:** 10.64898/2026.07.06.26357381

**Authors:** Alec Reinhardt, Zhiwen Jiang, Tengfei Li, Borui Tang, Xinjie Qian, Shuai Huang, Joseph G. Ibrahim, Yun Li, Bingxin Zhao, Patrick Sullivan, Jason Stein, Hongtu Zhu

**Affiliations:** Department of Biostatistics, University of North Carolina at Chapel Hill; Biomedical Research Imaging Center, UNC School of Medicine; Department of Radiology, UNC School of Medicine; Department of Genetics, University of North Carolina at Chapel Hill; Department of Statistics and Data Science, The Wharton School, University of Pennsylvania; Department of Psychiatry, University of North Carolina at Chapel Hill

## Abstract

Genetic variation, circulating metabolism and brain disease are each linked to white matter, but whether they converge on shared tract segments is unknown. Here we map genetic effects at 609,000 locations along 6,090 fiber pathways in 56,313 UK Biobank participants and integrate circulating proteomic and metabolomic associations. We identify 315, 238 and 345 loci for fractional anisotropy, mean and axial diffusivity; 227 fractional-anisotropy loci have no white-matter association in the GWAS Catalog. Footprints range from single locations to multi-tract patterns, concentrating in capsular and limbic tracts. Nearly all loci replicate at the locus level in 15,872 held-out adults, and 47% of fractional-anisotropy footprints recur in 7,403 children. Colocalization and Mendelian randomization link localized effects to brain and cardiometabolic disease, with shared causal variants at SPI1 for Alzheimer’s disease, APOE/APOC1 for coronary atherosclerosis and SREBF1/PEMT for stroke. Testing whether colocalized loci act where disease genetic correlation concentrates recovers the APOE/APOC1 link to Alzheimer’s disease but shows that shared variants rarely imply shared anatomy. Circulating fatty acids, led by linoleic acid, show the broadest associations, overlapping genetic hotspots in the corpus callosum. The atlas places inherited risk and lipid metabolism on shared white-matter segments and nominates lipid and lysophosphatidic-acid-signalling genes for follow-up.

## Main

White-matter tracts connect cortical and subcortical systems and support communication across the human brain [1, 2]. Their microstructure contributes to cognition and behaviour and is altered in many neuropsychiatric and neurodegenerative disorders [3, 4]. Diffusion magnetic resonance imaging (MRI) provides indirect indices of tissue organization, including fractional anisotropy (FA), mean diffusivity (MD) and axial diffusivity (AD), which are sensitive to axonal geometry, myelination and other tissue properties [5–9]. Twin and family studies have established substantial heritability of these measures [10–12], and large-scale genome-wide association studies (GWAS) have revealed a highly polygenic architecture with genetic relationships to cognitive and psychiatric traits [13–16].

Where along a tract these genetic effects act remains largely unknown. Tract averages, tract-specific functional principal components, structural connectivity phenotypes and latent representations of diffusion maps have expanded genetic discovery [14, 17–19], but each summarizes white matter at a scale broader than a position along a pathway and cannot show how the effect of an individual locus is distributed within a tract. An anatomical reference organized along pathways would allow the location and extent of each locus’s association footprint to be compared across cohorts and with molecular and disease maps.

Circulating proteins and metabolites provide complementary information about systemic processes, including lipid metabolism, inflammation and extracellular signalling. Previous multi-omic interpretation of white-matter GWAS has relied on functional annotation, enrichment analysis and predicted interaction networks [19]. Directly measured circulating molecules add a different view: their associations with diffusion phenotypes can be mapped onto the same anatomy as genetic associations, identifying tract segments where inherited variation and systemic molecular state coincide. An analogous question arises for disease genetics. Colocalization can support a shared causal variant between an imaging phenotype and a disease, but not where in the brain that variant acts. A complementary test asks whether a locus’s association footprint coincides with the white-matter regions where the disease’s genetic correlation concentrates, distinguishing genomic sharing from anatomical correspondence, which tract averages cannot resolve.

Here we constructed a molecular-genetic atlas of human white matter by combining connectome-based spatial statistics [20] with efficient genome-wide analysis of high-dimensional imaging phenotypes [21] (Fig. 1). Using 56,313 UK Biobank (UKB) participants for discovery, we mapped genetic associations along 6,090 atlas-aligned pathways sampled at 609,000 locations, evaluated replication in 15,872 held-out UK Biobank participants and in 7,403 children from the Adolescent Brain Cognitive Development (ABCD) study, and integrated circulating proteomic and metabolomic associations from available subsets.

**Figure 1:**
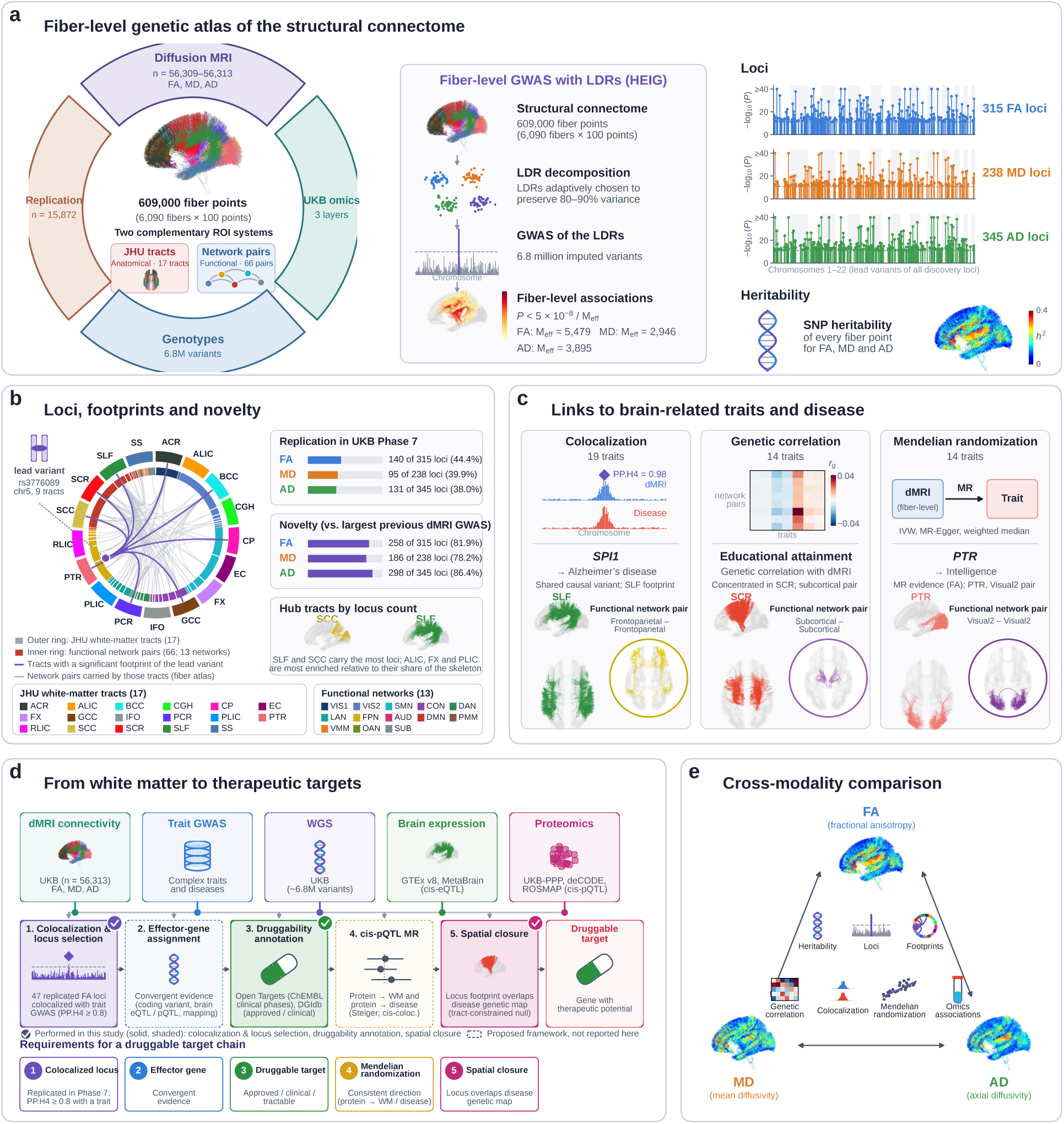
A fiber-level genetic and molecular atlas of the human structural connectome. **a**, Study design. Diffusion MRI from 56,313 UK Biobank participants (discovery) and 15,872 held-out participants (replication) was mapped onto a connectome atlas of 6,090 fiber pathways sampled at 100 points each (609,000 locations) and analysed with 6.8 million variants and three circulating omics layers, organized by 17 JHU tracts and 66 functional network pairs. Centre: signals in each of 430 fiber clusters are compressed into low-dimensional representations (LDRs), GWAS is run on the LDRs and statistics are reconstructed at every fiber point. Right: lead variants of all discovery loci (315 FA, 238 MD and 345 AD) and SNP heritability at every fiber point. **b**, Loci, footprints and novelty. Circos plot of the FA lead variant rs3776089, whose footprint is enriched in nine tracts (outer ring) and their network pairs (inner ring). Bars give spatial replication in UKB Phase 7 (*>*50% footprint overlap: 140, 95 and 131 loci) and the number of loci not reported by the largest previous diffusion-MRI GWAS [14] (258, 186 and 298). The SLF and splenium carry the most loci by count; enrichment relative to tract size is highest in the ALIC, fornix and PLIC (Fig. 5a). **c**, Links to brain-related traits and disease, one exemplar per analysis: colocalization across 19 traits (*SPI1* locus at 11p11.2 with Alzheimer’s disease, PP.H4 = 0.98), genetic correlation across 14 traits at the network-pair level (educational attainment) and Mendelian randomization on 14 outcomes (MR evidence for FA in the PTR on intelligence; Visual2–Visual2 pair). **d**, From white-matter loci to therapeutic targets: a five-step chain from colocalization and locus selection through effector-gene assignment, druggability annotation (Open Targets with ChEMBL-derived clinical phases, and DGIdb), cis-pQTL MR and spatial closure. Steps 1, 3 and 5 (solid, check marks) are performed here; steps 2 and 4 (dashed) are proposed. **e**, All analyses are run for FA, MD and AD. Tract abbreviations are defined in the Methods.

We find that genetic effects on white matter are not uniform along tracts: loci act on segments ranging from single locations to patterns spanning several tracts, with the highest density in compact capsular and limbic pathways, a distribution that tracks tract-level heritability for FA but not for MD or AD. Most loci were not detected by tract-level GWAS; nearly all replicate in held-out adults, and nearly half of FA footprints recur in children. Shared causal variants link these loci to brain and cardiometabolic disease, and circulating fatty acids co-locate with genetic hotspots in the corpus callosum, pointing to lipid and membrane metabolism as a candidate biological axis. Spatial tests further show that shared genetic signal seldom implies shared anatomy: of 56 colocalized locus–trait pairs with a disease map, only one, in the major histocompatibility complex, overlapped the map of its own trait. Together, these findings provide a common anatomical framework for relating inherited variation, circulating molecular state and disease susceptibility.

### Representation learning and quality control of location-level diffusion metrics

To enable fine-grained genetic analysis of white-matter microstructure at biobank scale while preserving anatomical interpretability, we used the connectome-based spatial statistics (CBSS) framework [20]. CBSS encodes diffusion MRI signals along atlas-aligned fiber trajectories rather than isolated voxels, maintaining tract topology and functional network organization (Fig. 1a; Supplementary Fig. 1). The white-matter skeleton was reconstructed into 6,090 fiber bundles, each sampled at 100 along-tract nodes, yielding 609,000 ordered fiber-grid locations. To reduce redundancy and ensure computational efficiency for genome-wide testing, these locations were hierarchically aggregated into 430 anatomically coherent clusters (Supplementary Table 1), constrained by functional network compartments and local neighborhoods. This preserves biological organization while allowing full reconstruction of location-level signals.

Within each cluster, we applied functional principal component analysis (fPCA) to derive low-dimensional representations (LDRs) [21]. Conceptually, fPCA extracts smooth spatial modes of microstructural variation along fiber trajectories, identifying coherent patterns rather than voxelwise noise. Graphically, each LDR can be viewed as a compact “basis function” describing a canonical spatial motif (e.g., a gradient or focal change), and each participant’s tract profile is expressed as a weighted combination of these motifs. Dimensionality was selected by an adaptive rule targeting 80–90% of variance per cluster, yielding on average 20.6 LDRs (8,868 total phenotypes; Supplementary Table 1). Reconstruction fidelity was high (correlation = 0.83–0.99; Supplementary Fig. 2c), and leading components showed anatomically smooth, biologically plausible patterns (Supplementary Fig. 3a,c). Wild-bootstrap null simulations demonstrated well-calibrated type-I error across tracts (Supplementary Fig. 3b), supporting the validity of these representations for downstream inference. Because every location-level statistic is a linear combination of the LDR statistics of its cluster, the effective spatial resolution of all location-level maps is set by the 8,868 LDRs and the effective number of independent locations (*M*_eff_ = 5,479; Methods), not by the 609,000 sampling points themselves.

### GWAS and heritability of white matter microstructure

Using the UK Biobank discovery cohort (Phase 1–6; n = 56,313, white British ancestry), we per-formed GWAS on 8,868 low-dimensional FA representations spanning 430 fiber clusters (609,000 locations). Projecting summary statistics back into image space revealed extensive genetic influence on white-matter microstructure: after correction for the effective number of voxel-level tests (*P <* 9.12 *×* 10*^−^*^6^), 90.7% of locations were significantly heritable in FA, with 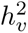 ranging from 0 to 39.3% (mean = 16.4%) (Fig. 2a,b; Supplementary Fig. 4; Supplementary Table 3). Aggregated estimates confirmed robust heritability across all 17 major tracts (mean = 19.3%) and 66 functional network pairs (mean = 17.3%) (Fig. 2a; Supplementary Table 3). The same analysis for MD and AD gave similarly pervasive heritability (94.2% and 93.4% of locations significant). Tract-level heritability was organized similarly across the three measures, but the within-tract spatial patterns only partly overlapped (median within-tract correlation of location-level *h*^2^: FA–MD *r* = 0.53, FA–AD 0.64, MD–AD 0.73; Fig. 2a,b; Supplementary Results §2.3).

**Figure 2:**
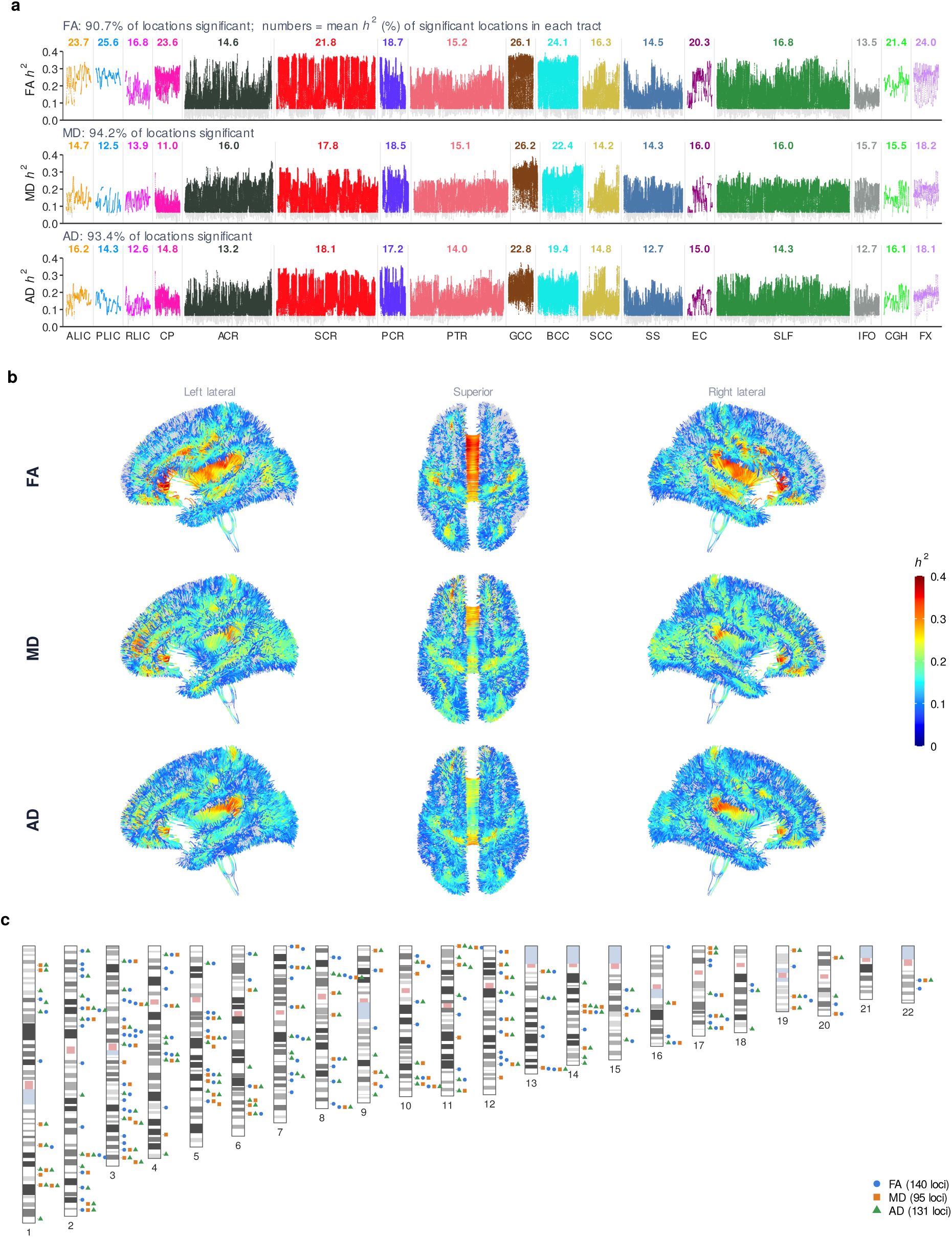
SNP heritability of white-matter microstructure at every fiber location, and replicated loci across the genome. **a**, Location-level SNP heritability (*h*^2^) of FA (top), MD (middle) and AD (bottom) in the UK Biobank discovery cohort (n = 56,313), plotted for every fiber location grouped by JHU tract (colours) and ordered along each tract. Grey points are locations that are not significantly heritable after correction for the effective number of independent locations (*P <* 9.12 *×* 10*^−^*^6^); 90.7%, 94.2% and 93.4% of locations are significant for FA, MD and AD. The number above each tract is the mean *h*^2^ (%) of its significant locations. Heritability is highest in the genu of the corpus callosum (FA 26.1%, MD 26.2%, AD 22.8%) and in the internal capsule, and is most uniform within compact projection tracts (for example PLIC, FA SD = 2.7%) and most variable within large association and callosal systems (for example SCR and GCC, SD *≈* 10%). **b**, The same estimates projected onto the fiber atlas (left lateral, superior and right lateral views; colour scale *h*^2^ 0–0.4). For all three measures, heritability peaks along the body and genu of the corpus callosum, the internal capsule and the sagittal stratum, and declines toward the cortical terminations of superficial fibers; the MD and AD maps are more spatially patchy than the FA map. **c**, Chromosomal ideogram of the loci that replicated spatially in UKB Phase 7 (*>*50% overlap of the discovery footprint; n = 15,872): 140 FA loci (blue circles), 95 MD loci (orange squares) and 131 AD loci (green triangles) placed at the position of the discovery lead variant. Replicated loci are distributed across all 22 autosomes, and most are specific to one or two measures. Tract- and network-pair summaries of the location-level estimates for FA, MD and AD (mean, SD, proportion of significant locations and ACAT *P*-values) are given in Supplementary Table 3; FA–MD–AD comparisons are in Supplementary Fig. 4 and Supplementary Results §2.3.

Heritability patterns exhibited clear anatomical organization. The genu of the corpus callosum (GCC) carried the highest mean heritability over its significant locations (26.1%; 25.1% over all GCC locations, Supplementary Table 3), and the posterior limb of the internal capsule (PLIC) the most uniform (mean = 25.6%, SD = 2.7%), consistent with strong genetic regulation of tightly packed projection pathways. In contrast, association and callosal systems such as the superior corona radiata (SCR) (mean = 21.8% over significant locations; SD = 10.0%) and the GCC (SD = 9.5%) displayed greater within-tract variability, reflecting more heterogeneous microstructural organization (Fig. 2a). The brain maps make the anatomy explicit: for all three measures, heritability peaks along the body and genu of the corpus callosum and in the internal capsule and sagittal stratum, and is lowest at the cortical terminations of superficial fibers (Fig. 2b). These spatial patterns were highly reproducible in the UKB Phase 7 cohort (n = 15,872; r = 0.72 location-wise; r = 0.93 cluster-wise) (Supplementary Table 3).

Applying both genome-wide significance (*P <* 5 *×* 10*^−^*^8^; voxel-adjusted *P <* 9.12 *×* 10*^−^*^12^) and empirical spatial screening identified 1,185 tract-level loci, which consolidated into 315 distinct loci; parallel analyses of MD and AD identified 238 and 345 distinct loci, respectively (Fig. 1a). The FA loci spatial footprints ranged from focal (single-location) to highly distributed (up to 185,624 locations; median = 118), and nearly half (47.6%) influenced more than two tracts, indicating a mixture of pathway-specific and cross-system genetic effects (Supplementary Table 5). Replication in the held-out UKB Phase 7 cohort (*n* = 15,872; same scanners, protocol and ancestry as discovery) was assessed at the locus level and at the footprint level. At the locus level, 311 of 315 (98.7%) FA loci had a variant within *±*250 kb of the discovery lead that passed the same spatial screen at *P <* 10*^−^*^4^ in at least one tract (a threshold-based criterion that does not test the direction of effect at each locus); replication effect sizes scaled with discovery effect sizes (slope = 0.68, SE = 0.05, *P <* 2 *×* 10*^−^*^16^; the slope below 1 is consistent with winner’s curse in discovery), and 44% (140 loci) showed *>*50% spatial overlap of the discovery footprint (Fig. 1b; Fig. 2c). Locus-level replication was similarly high for MD (233/238, 97.9%) and AD (339/345, 98.3%), with 40% (95 loci) and 38% (131 loci) showing *>*50% spatial overlap, respectively. The replicated loci are distributed across all 22 autosomes, and the three measures only partly share them (Fig. 2c). Relative to the largest prior GWAS of tract-level white-matter microstructure [14], 57, 52 and 46 of the FA, MD and AD loci fall within *±*500 kb of a previously reported locus, and 258, 186 and 298 (81.9%, 78.2% and 86.4%) do not (Fig. 1b); 115 of the 258 FA loci not reported by Zhao et al. [14] also showed *>*50% spatial replication.

A broader GWAS Catalog lookup indicated that 88 of 315 FA loci (27.9%) had been previously linked to FA or other white-matter phenotypes in any study, so that 227 FA loci (72.1%) are novel with respect to all catalogued white-matter GWAS. Beyond white matter, the loci showed broad pleiotropy: 293 loci (93.0%) overlapped regions associated with neuropsychiatric or cognitive traits (395 traits total), including educational attainment (120 loci), schizophrenia (27), Alzheimer’s disease (20), major depressive disorder (18), bipolar disorder (9), and Parkinson’s disease (7). More than 70% also overlapped loci for non-neurological traits, such as bone density (83.4%), cardiac function (81.4%) and refractive error (71.2%), highlighting the cross-organ pleiotropic architecture underlying white-matter biology.

Location-level spatial atlases clarified this architecture. Many loci produced focal, tract-specific signatures concentrated in projection and callosal pathways: for example, the 13q14.11 locus (rs1747223) was confined to 51 locations in the genu of the corpus callosum, posterior corona radiata and posterior thalamic radiation (Supplementary Fig. 37), and the 18p11.21 locus (rs2306811), detected for MD and AD but not FA, was restricted to the body and genu of the corpus callosum and the superior corona radiata (Supplementary Fig. 38). Others showed broad, distributed signatures spanning multiple association fibers and callosal tracts (Supplementary Figs. 35–36 and 39); the 12q23.3 locus (rs12146713) illustrates this pattern, with associations spanning 12 FA tracts (including the corpus callosum, internal capsule, corona radiata and superior longitudinal fasciculus) and 71.8% location-level spatial overlap in the replication cohort (Supplementary Figs. 25–26). Hotspot maps converged on the corpus callosum, corticospinal tract and fronto-occipital fasciculus, and locus density relative to each tract’s share of the fiber skeleton was highest in compact projection and limbic tracts, with odds ratios (OR) of 54 for the anterior limb of the internal capsule (ALIC), 42 for the fornix (FX) and 37 for the PLIC (Fig. 1b; Fig. 5a; Supplementary Figs. 27–29). Locus enrichment was coupled to tract-level heritability for FA but not for MD or AD. Across the 17 tracts, locus enrichment and mean *h*^2^ were positively rank-correlated for FA (Spearman *ρ* = 0.55, *P* = 0.023), because the compact capsular and limbic tracts are both strongly heritable and strongly enriched, but uncorrelated for MD (*ρ* = *−*0.24, *P* = 0.37) and AD (*ρ* = 0.17, *P* = 0.52); even for FA, the most heritable tract, the GCC (Fig. 2a), ranked only tenth of 17 in FA enrichment (OR = 6.9; Fig. 5a). Because the odds ratios are normalized to each tract’s share of the skeleton, compact tracts attain large values from few loci, so these ranks describe locus density rather than the number of loci. Together, these results indicate that projection systems carry a larger share of localized effects, whereas association and callosal networks are shaped by broader, multi-tract influences.

### Validation on ABCD

To assess whether the adult loci are detectable earlier in development, we examined FA in the Adolescent Brain Cognitive Development (ABCD) cohort (n = 7,403 children aged 9–10 years; multi-site and multi-ancestry), adjusting for acquisition site. Because ABCD is more than seven-fold smaller than the discovery cohort, we did not expect genome-wide significance and instead asked whether the discovery footprint of each locus was reproduced at a nominal threshold (*P <* 1 *×* 10*^−^*^2^) with *>*50% spatial overlap. Across the 315 FA loci, 149 (47.3%) met this criterion, and 72 (22.9%) met it in both UKB Phase 7 and ABCD (Supplementary Fig. 24; Supplementary Table 5).

The loci reproduced in ABCD were spread across all 17 tracts. Relative to the number of loci each tract carries, they were most over-represented in the sagittal stratum (49 of its 79 loci; odds ratio 2.2, nominal Fisher’s exact *P* = 0.003), and no other tract was nominally enriched. Despite differences in cohort composition and developmental stage, part of the localized adult architecture is therefore detectable in late childhood.

### Links between white matter microstructure and diseases

We quantified shared polygenic architecture between FA and 14 neuropsychiatric, neurodegenerative and cognitive traits using location-level genetic correlation (Methods) [22]. Nine of the 14 external GWAS include UK Biobank participants (Supplementary Table 2), so these estimates, and the Mendelian randomization estimates below, may be inflated by sample overlap; traits from UKB-free GWAS (schizophrenia, bipolar disorder, major depressive disorder, ADHD and autism) are unaffected. Location-wise genetic correlations (*r_g_*) revealed distributed, network-structured patterns across the white-matter skeleton (Fig. 3c). Schizophrenia, educational attainment, and bipolar disorder showed the broadest signatures, each with significant correlations in *∼*1% of locations after correction (*P <* 9.12 *×* 10*^−^*^6^; Supplementary Table 4). Averaged across the brain, Alzheimer’s disease exhibited the strongest positive mean correlation (mean of location-level *r_g_* = 0.083) (Fig. 3c; Supplementary Fig. 16), while depression showed the strongest negative correlation (mean *r_g_* = *−*0.053) (Fig. 3c; Supplementary Fig. 12), with major depressive disorder at *−*0.036 (Supplementary Fig. 14). The positive sign for Alzheimer’s disease (higher genetic liability associated with higher FA) runs counter to the lower FA seen in manifest disease, and should be interpreted with caution because the Alzheimer’s disease GWAS includes UKB parental-history proxy cases and because FA–age relationships differ between the imaging cohort and the ages at which the disease manifests. At the tract level, each tract–trait cell was classified from two complementary tests on its location-level estimates (Methods): a signed, precision-weighted aggregate that asks whether the correlation is consistently of one sign across the tract, and a dependence-robust Cauchy combination that is sensitive to signal of either sign. Of the 238 FA cells, 145 were significant after FDR correction across all cells (Fig. 3a; Supplementary Table 4), 112 (77.2%) by the signed test, indicating a consistent direction across the tract’s fiber clusters, and 33 by the Cauchy test only, indicating mixed-sign signal. Because the signed aggregate treats fiber clusters as independent, its standard error is anti-conservative and we report it only as a classification statistic; the magnitude of a tract-level correlation is given by the mean of its location-level *r_g_* values (for example 0.113 for the SCR and Alzheimer’s disease), and 70 of the 145 cells were also significant under the Cauchy test. The most prominent associations mapped onto the superior corona radiata and inferior fronto-occipital fasciculus (IFO) for Alzheimer’s disease, and the posterior limb of the internal capsule for depression (Supplementary Table 4); for Alzheimer’s disease, however, only the IFO cell passed the Cauchy test and no single FA location passed the location-level threshold, so its relationship with FA is best described as small but consistently positive across tracts rather than locally significant. Aggregation to the 66 functional network pairs showed the same organization with a clear sign structure (Fig. 3b): Alzheimer’s disease, cognitive performance and educational attainment were positively correlated with FA across many network pairs, internalizing traits (depression, depressed affect, neuroticism, worry and insomnia) were consistently negatively correlated, and schizophrenia and bipolar disorder showed many significant pairs, more than half of them mixed in sign (as were most of the significant pairs for major depressive disorder), consistent with their broad but non-uniform location-level signatures.

**Figure 3:**
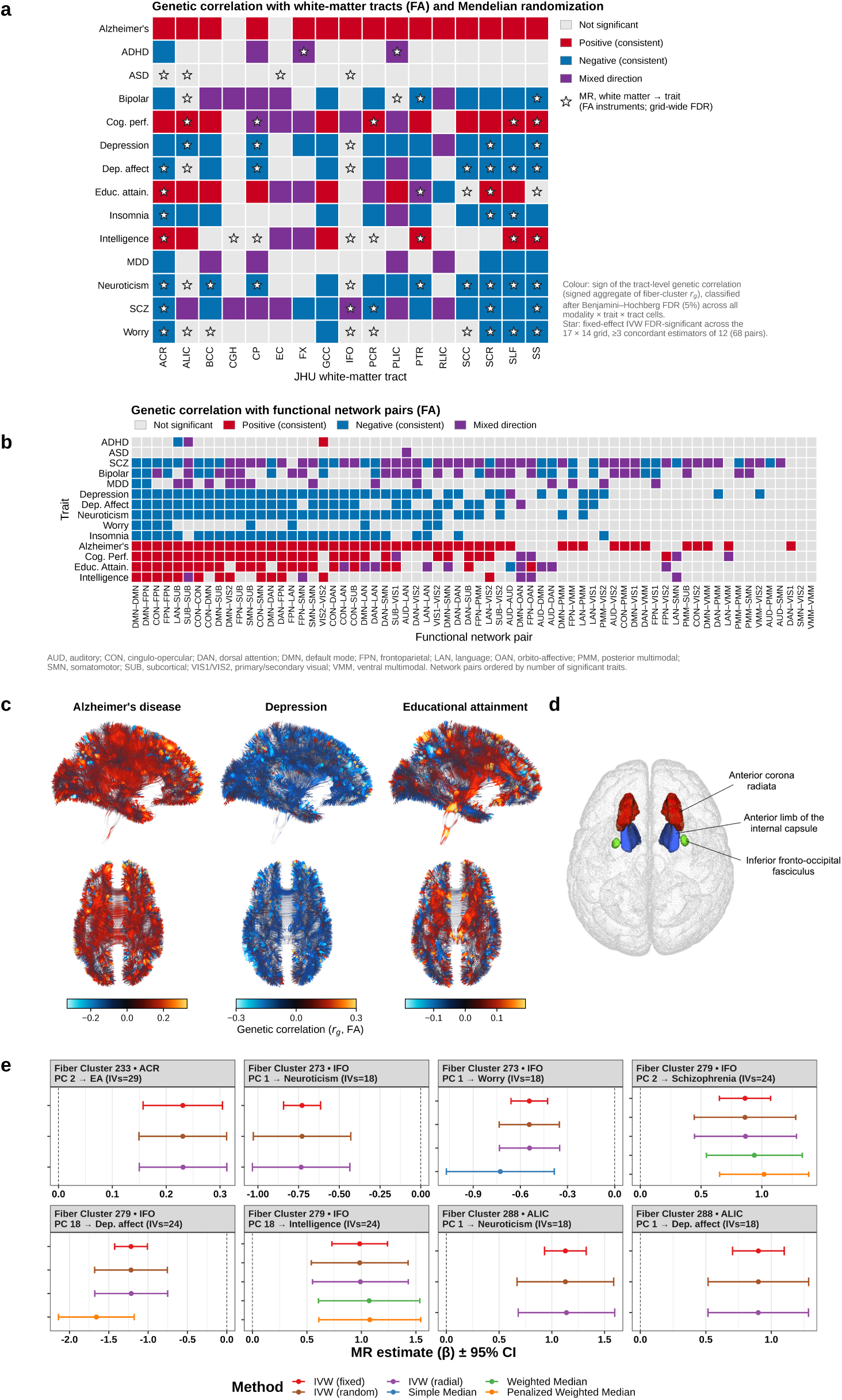
Genetic correlation and Mendelian randomization between white-matter microstructure and brain-related traits. **a**, Tract-level genetic correlation (GC) between FA and 14 traits, with Mendelian randomization (MR) overlaid. Cells (JHU tract *×* trait) are coloured by the classification of the tract-level genetic correlation after FDR correction across all measure *×* trait *×* tract cells: signed aggregate significant and positive (red) or negative (blue), Cauchy test significant only, that is, mixed in sign (purple), or not significant (grey); 145 of 238 FA cells are significant, 70 of them also by the Cauchy test (Supplementary Table 4). Stars mark the 68 pairs with MR evidence (FA fiber-cluster instruments; fixed-effect IVW FDR-significant across the 17 tracts *×* 14 outcomes, and at least three concordant estimators of twelve, IVW included; Supplementary Table 9). Neuroticism has MR evidence in 10 tracts, schizophrenia in 5, and Alzheimer’s disease and major depressive disorder in none. **b**, Genetic correlation between FA and the same traits at the 66 functional network pairs (columns ordered by number of significant traits), coloured as in **a**. AUD, auditory; CON, cingulo-opercular; DAN, dorsal attention; DMN, default mode; FPN, frontoparietal; LAN, language; OAN, orbito-affective; PMM, posterior multimodal; SMN, somatomotor; SUB, subcortical; VIS1/VIS2, primary/secondary visual; VMM, ventral multimodal. **c**, Location-level genetic correlation maps (lateral and superior views) for Alzheimer’s disease (mean location-level *r_g_* = 0.083), depression (mean *r_g_* = *−*0.053) and educational attainment; colour scales give *r_g_* for FA. **d**, Tracts hosting the fiber-cluster-level MR exemplars in **e**, superior view: anterior corona radiata (red), anterior limb of the internal capsule (blue) and inferior fronto-occipital fasciculus (green). **e**, Forest plots of eight exemplar fiber-cluster–trait pairs (IVs, number of instruments): MR estimate (*β*) *±* 95% CI for the estimators that are FDR-significant for that pair. Because the sign of an LDR is arbitrary, the sign of *β* does not indicate whether higher FA raises or lowers the outcome. ADHD, attention-deficit/hyperactivity disorder; ASD, autism spectrum disorder; EA, educational attainment; MDD, major depressive disorder; SCZ, schizophrenia.

To test whether these correlations reflected shared causal variants, we performed Bayesian colocalization at the 140 spatially replicated FA loci against a panel of 19 external traits: Alzheimer’s disease, ADHD, autism, bipolar disorder, cognitive performance, educational attainment, intelligence, insomnia, major depressive disorder, schizophrenia, Parkinson’s disease, reaction time, drinks per week, number of sexual partners, coronary atherosclerosis, stroke, type 2 diabetes, glomerular filtration rate and liver disease (Supplementary Table 2). Four of the 2,660 locus–trait pairs could not be evaluated (at the 17q21.31 locus, too few shared variants for cognitive performance and educational attainment and a failed fit for reaction time; missing sample sizes for one locus with bipolar disorder; see the notes to Supplementary Table 8) and were treated as not colocalized. Applying stringent thresholds (PP.H4 *≥* 0.8 and PP.H4 *>* PP.H3), 47 loci colocalized with at least one of 17 of these traits (Supplementary Table 8). Of these, 35 were not reported by Zhao et al. [14], indicating that newly resolved FA loci intersect established neuropsychiatric and cardiometabolic risk regions at the level of shared causal variants. Colocalization spanned both brain-specific pathways (e.g., schizophrenia: 8 loci; educational attainment: 7 loci) and systemic physiology (e.g., glomerular filtration rate: 9; type 2 diabetes: 7). For prioritized loci, aligned association peaks across FA and external traits supported shared local genetic drivers (Supplementary Figs. 30–34). Several of these loci implicate genes with established roles in lipid and membrane metabolism: at 19q13.32, a locus spanning APOE and the neighboring APOC1/APOC2 apolipoproteins colocalized with coronary atherosclerosis (PP.H4 = 0.995); at 17p11.2, a locus containing SREBF1, a master transcriptional regulator of cholesterol and fatty-acid synthesis [23], and PEMT, which catalyzes phosphatidylcholine biosynthesis [24], colocalized with stroke (PP.H4 = 0.986); and at 11p11.2, a locus containing the myeloid transcription factor SPI1 [25] colocalized with Alzheimer’s disease (PP.H4 = 0.982); the lipid-sensing nuclear receptor NR1H3 [26] lies 136 kb away at a separate MD/AD lead (rs10838681, *r*^2^ = 0.12 with the FA lead) that did not colocalize with any trait. Two additional loci linked white-matter microstructure to type 2 diabetes (5q13.3, PP.H4 = 0.997) and glomerular filtration rate (6p25.1, PP.H4 = 0.992), indicating that shared variants extend to systemic metabolic and vascular physiology beyond lipid pathways. Colocalization at a single representative location with default priors is sensitive to the prior on a shared variant and to the presence of multiple causal variants, and the PP.H4 values should be read with this in mind; in particular, the 6p21.33 locus lies in the extended major histocompatibility complex (MHC), where single-causal-variant colocalization is unreliable.

We next asked whether genetically predicted white-matter microstructure is associated with each trait using two-sample Mendelian randomization (MR) with fiber-cluster LDRs as exposures [27]. The 14 MR outcomes are the same GWAS as the genetic-correlation panel, with the APOE region removed from the Alzheimer’s disease outcome (Supplementary Table 2). Requiring fixed-effect inverse-variance-weighted (IVW) significance after FDR correction across the full tract *×* trait grid and at least three concordant estimators (fixed-effect IVW included), 68 tract–trait pairs showed consistent MR evidence (Fig. 3a, stars; Supplementary Table 9). These signals were trait-dependent: neuroticism, worry, depressed affect and intelligence showed MR evidence in 10, 9, 8 and 8 of 17 tracts; cognitive performance, educational attainment, schizophrenia and depression in 5 each; autism spectrum disorder and bipolar disorder in 4 each; insomnia in 3 and ADHD in 2. Neither major depressive disorder nor Alzheimer’s disease met the criteria in any tract. For Alzheimer’s disease this contrasts with a significant genetic correlation in 15 of 17 tracts: with the APOE region removed there is no evidence for a directional effect of white-matter microstructure on Alzheimer’s disease risk, and the genetic correlation is more consistent with shared genetic architecture. Three caveats apply to all MR results. First, the outcome GWAS for Alzheimer’s disease, insomnia, the cognitive traits and the neuroticism-family traits include UK Biobank participants, so these estimates are subject to sample-overlap bias toward the observational association. Second, the sign of an LDR is arbitrary, so the MR results show whether an association is consistent across estimators, not whether higher FA raises or lowers the outcome. Third, the reverse direction (trait to white matter) was not tested, and the many estimators are not independent, and every reported pair is supported by the three IVW estimators only (Methods); the MR results therefore nominate rather than establish directional effects.

Across traits, MR evidence concentrated in the sagittal stratum, the anterior and superior corona radiata, the anterior limb of the internal capsule and the inferior fronto-occipital fasciculus (7–9 traits each; Fig. 3a). At the fiber-cluster level, the clearest effects arose from clusters in the inferior fronto-occipital fasciculus, anterior limb of the internal capsule and anterior corona radiata (Fig. 3d), where single clusters carried consistent effects on several traits (for example, IFO cluster 279 on depressed affect, intelligence and schizophrenia; IFO cluster 273 and ALIC cluster 288 on neuroticism; Fig. 3e). Across the 68 significant tract–trait pairs, a median of 5.5 of the 12 estimators were FDR-significant (range 3–10), but the estimators that were also directionally concordant were, in every pair, the three IVW estimators (Methods).

Together, these results show that white-matter microstructure shares extensive network-level polygenic architecture with brain-related traits, with MR evidence that is broadest for internalizing phenotypes and intelligence, narrower for the other cognitive and psychiatric traits and absent for Alzheimer’s disease and major depressive disorder.

### Multi-omics association atlases

We next examined molecular correlates of white-matter microstructure by performing location-level phenome-wide association analyses (PheWAS) between FA and three circulating molecular modalities (proteomics, metabolomics and clinical laboratory biomarkers) measured from peripheral venous blood in UK Biobank Phases 1–6. For each modality, location-wise association statistics were obtained from linear models adjusted for the same demographic and genetic covariates used in the GWAS, with sample sizes of 5,614 (proteomics), 26,412 (metabolomics), and 18,689 (clinical biomarkers); blood samples were collected at the baseline visit, several years before imaging, and the models did not adjust for body-mass index, lipid-lowering medication, smoking or fasting time, so the associations describe systemic state rather than isolated molecular effects. Multiplicity correction accounted for the effective number of independent locations (*M*_eff_ = 5,479) and of omics features (25 of 2,923 proteins, 5 of 502 metabolomic measures and 13 of 57 biomarkers, from the eigenspectrum of each feature correlation matrix), and spatial screening was performed using a wild-bootstrap null model. This analysis identified 7 proteomic, 137 metabolomic, and 8 clinical-biomarker features with significant location-level associations with FA (Supplementary Table 10); the 137 metabolomic features are highly inter-correlated (the Nightingale fatty-acid fractions in particular are near-deterministic functions of one another), and correspond to far fewer independent signals. In the held-out Phase 7 cohort, 108 of the 137 metabolomic features (79%), but only 1 of 7 proteomic and 2 of 8 biomarker features, reproduced *≥*10% of their discovery footprint (Supplementary Results §2.7; Supplementary Fig. 41), so the proteomic and biomarker layers should be regarded as exploratory. Metabolomic features showed the broadest and most anatomically coherent associations, with extensive clusters of significant voxels across major projection and callosal pathways, most prominently the corpus callosum (Fig. 4a,b,d). The largest footprints belonged to fatty-acid composition measures (linoleic acid, omega-6, polyunsaturated, saturated and monounsaturated fatty acids as fractions of total fatty acids), each significant at tens of thousands of locations and overlapping the genomic hotspot mask at 90% or more of their extent, followed by the glucose/lactate ratio and large-HDL lipid measures (Fig. 4a). The strongest associations therefore involved linoleic acid and the other fatty-acid fractions; phosphatidylcholines ranked 23rd of the 137 metabolomic features by extent (9,732 locations, 66% of them reproduced in Phase 7; Supplementary Table 10), and their regional effects concentrated in commissural and corticospinal tracts (Fig. 4d, left). Regions showing significant metabolomic associations overlapped the genomic association hotspots (Dice = 0.67; Fig. 4b). Because the corpus callosum is the largest and most heritable structure in the skeleton, part of this overlap is expected from the size and location of the two masks alone, and the Dice value is reported as a descriptive measure of co-location rather than as evidence of a shared cause. Proteomic associations were focal: the most extensive, BAIAP2, NPY, SYT1 and ADGRG1, covered tens to hundreds of locations, and only ADGRG1 (GPR56, a receptor required for oligodendrocyte development) fell entirely within the genomic hotspot mask (Fig. 4a,d, middle). Clinical-biomarker associations were the sparsest and most spatially scattered of the three molecular layers, with albumin-adjusted calcium, free thyroxine and iron showing the largest extents (Fig. 4a,d, right). The seven proteomic features are BAIAP2, NPY, SYT1, AD-GRG1, MOCS2, CEACAM18 and CALCB, the last significant at a single location (Fig. 4a).

**Figure 4:**
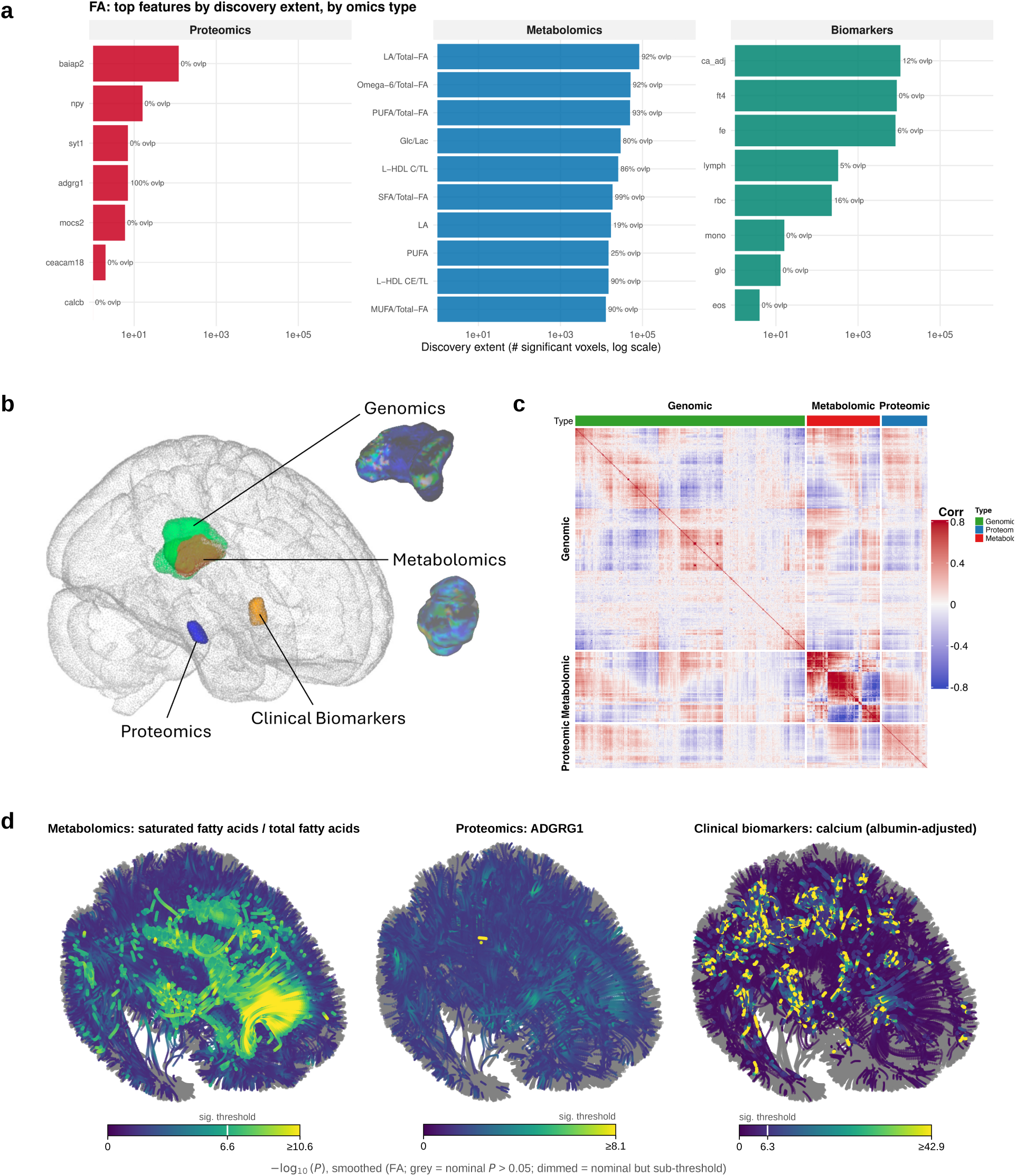
Multi-omics association atlases of FA. **a**, Circulating features with the largest discovery extent (number of significantly associated fiber locations, log scale) in location-level phenome-wide association analyses of FA against plasma proteomics (n = 5,614; red), NMR metabolomics (n = 26,412; blue) and clinical biomarkers (n = 18,689; green). The label at the end of each bar gives the fraction of the footprint lying within the genomic hotspot mask. Fatty-acid fractions, the glucose/lactate ratio and large-HDL measures have footprints of 10^4^–10^5^ locations, 80–99% within the hotspots; proteomic and biomarker footprints are one to three orders of magnitude smaller. **b**, Union of significant locations for each layer within a glass brain: genomics (green), metabolomics (orange; Dice = 0.67 with the genomic hotspots, largely within the corpus callosum and corona radiata), proteomics (blue) and clinical biomarkers (yellow). Insets isolate the genomic and metabolomic regions. **c**, Pairwise Pearson correlations between location-wise Z-score maps of all significant genomic (lead variants), metabolomic and proteomic features, ordered by hierarchical clustering within omics type (colour scale *−*0.8 to 0.8). Maps cluster by omics type, with the highest within-type concordance among metabolites (median *|r|* = 0.6). **d**, Example atlases (*−* log_10_ *P*, smoothed; left lateral view), one per layer: saturated-fatty-acid fraction (metabolomics), ADGRG1/GPR56 (proteomics) and albumin-adjusted calcium (clinical biomarkers). The white tick on each colour bar marks the significance threshold; grey locations have nominal *P >* 0.05. Full feature lists are in Supplementary Table 10 and feature correlations in Supplementary Fig. 40.

To assess cross-omic structure, we computed pairwise Pearson correlations of location-wise Z-score maps (Fig. 4c). Effect-map clustering occurred strongly by omics type, with metabolomic features exhibiting the highest within-type concordance (median *|r|* = 0.6), and the genomic block (lead variants of the FA loci) forming a largely independent set of spatial patterns with sparse off-diagonal structure (Fig. 4c). We also observed feature pairs with minimal phenotypic correlation (*|r| <* 0.1) but concordant spatial association patterns: circulating calcitonin gene-related peptide beta (CALCB) showed weak phenotypic correlations with LCAT (r = *−*0.008) and BID (r = *−*0.036), yet their FA effect maps were aligned (r = 0.51 and 0.58), and omega-6 fatty acid percentage and glutamine were weakly correlated phenotypically (r = 0.058) but spatially concordant (r = 0.78). Conversely, some phenotypically correlated features showed little overlap in their FA effects. Because all effect maps are reconstructed from the same fPCA bases and inherit the same spatial covariance, the null expectation for the correlation between two unrelated effect maps is not zero; these dissociations therefore indicate that marginal feature correlations do not predict shared white-matter signatures, rather than establishing convergent biology (Supplementary Fig. 40).

Finally, we quantified the overlap of statistically significant locations using Dice scores after FDR correction, identifying 12,663 of 36,585 feature pairs (34.6%) with substantial spatial overlap (Dice *≥* 0.1). Of these, 3,147 (24.8%) crossed omics-type boundaries, most commonly metabolomic–proteomic pairs (2,972), with fewer biomarker–metabolomic (128) and biomarker–proteomic (47) pairs. The strongest cross-omics overlaps involved metabolite–protein pairs, for example triglycerides in HDL with the inflammatory chemokines CCL7 (Dice = 0.55) and CCL3 (Dice = 0.46), and glucose with the lipid-metabolism enzyme LCAT (Dice = 0.51) and with ADGRG1 (Dice = 0.49), indicating that metabolic, inflammatory and neuronal-signaling features map onto shared white-matter pathways (Supplementary Table 10).

This anatomical co-location parallels, but does not by itself establish, a shared mechanism with the colocalized loci described above: PEMT (17p11.2), which colocalizes with stroke risk, is the biosynthetic enzyme for phosphatidylcholine, a metabolite class associated with FA in our PheWAS (23rd of 137 metabolomic features by extent). Whether this reflects a shared genetic driver between the locus and the metabolite was not tested here, and represents a natural target for future work integrating genetic, metabolomic, and disease colocalization evidence within a unified mediation framework.

### Biological Enrichment Analysis

To place the location-level genetic and omics associations into a broader biological framework, we performed multi-layered enrichment analyses. From our GWAS results, we identified tracts with significantly enriched signals and compared these tract-level profiles across FA, MD, and AD (Fig. 5a). Using both locus-based enrichment (Fisher’s exact test) and size-normalized enrichment relative to tract-specific fiber counts, we observed consistent overrepresentation of signals in compact projection and limbic pathways, including the anterior and posterior limbs of the internal capsule (ALIC: OR = 40–56; PLIC: OR = 31–44), the fornix (OR = 27–42), the external capsule (OR = 13–35), and the cingulum (hippocampal; OR = 22–37), indicating a shared anatomical substrate of genetic effects across diffusion modalities. Because these odds ratios are normalized to each tract’s share of the fiber skeleton, the large association tracts that carry the most loci in absolute terms (SLF: 135 FA loci; SCR: 121; ACR: 99) have the lowest enrichment, whereas the compact capsular and limbic tracts have the highest (Fig. 5a). Despite this overall concordance, notable modalityspecific differences emerged. FA-associated loci showed stronger enrichment in the cerebral peduncle (FA: OR = 13.1 versus MD: OR = 9.7, AD: OR = 9.8) and external capsule (FA: OR = 35.4 versus MD: OR = 23.0, AD: OR = 13.1). In contrast, MD showed markedly greater enrichment in the body of the corpus callosum (MD: OR = 9.0 versus FA: OR = 4.7, AD: OR = 6.1), and the posterior thalamic radiation was entirely absent from MD-associated loci (0 loci; FA: OR = 2.6, AD: OR = 2.4), suggesting differential sensitivity of diffusivity metrics to callosal and thalamic projection microstructure. Cross-modality comparison of enrichment profiles revealed high concordance (FA–MD: r = 0.96, FA–AD: r = 0.96, MD–AD: r = 0.98), supporting largely shared but non-identical genetic architectures across modalities.

**Figure 5:**
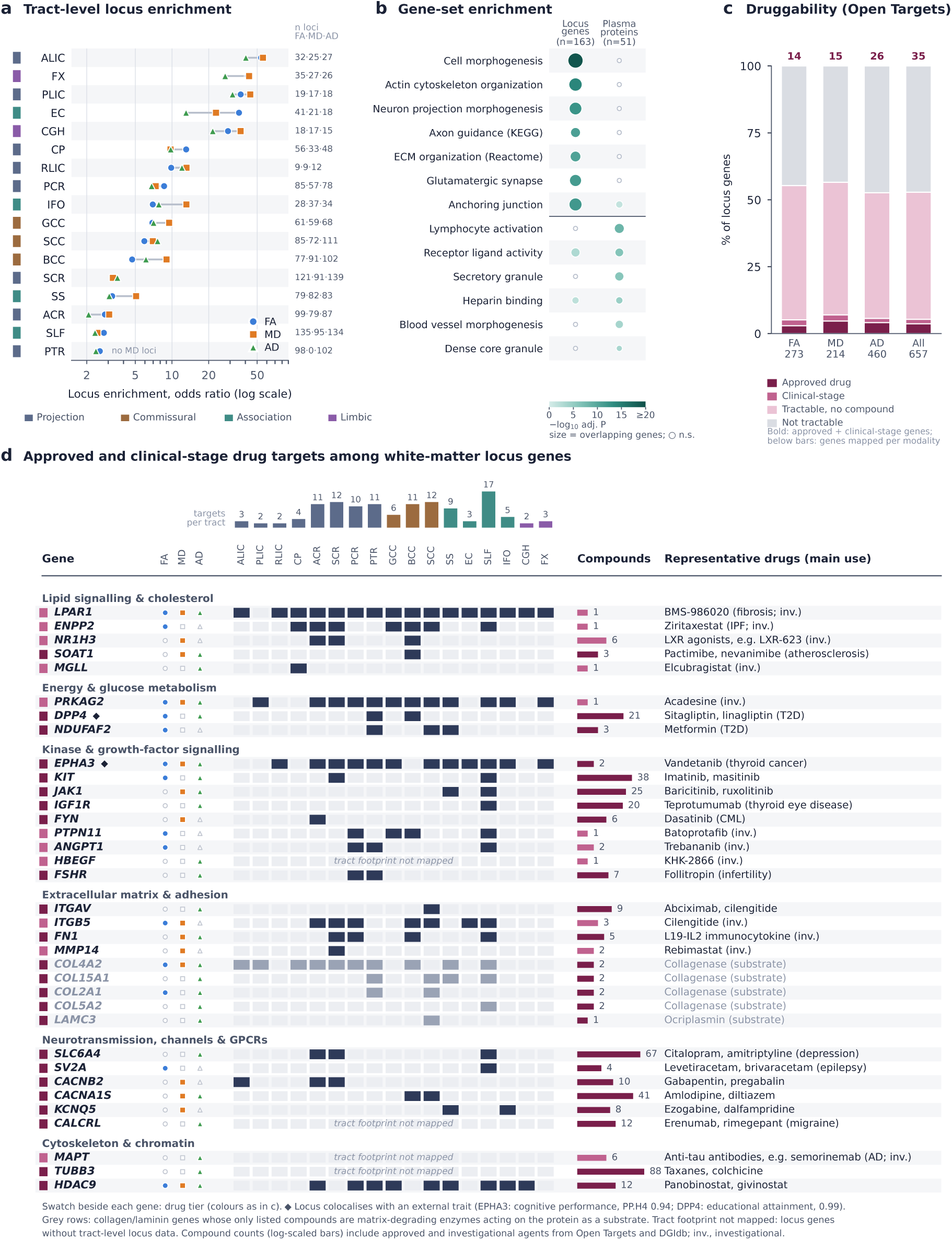
Anatomical and functional enrichment of white-matter loci, and the druggable subset of their genes. **a**, Tract-level locus enrichment for FA (blue circles), MD (orange squares) and AD (green triangles): odds ratio (log scale) normalized to each tract’s share of the fiber skeleton, with loci per tract (FA*·*MD*·*AD) at right and tract class at left. Compact capsular and limbic tracts (ALIC, FX, PLIC, EC, CGH) are the most enriched; the large association tracts that carry the most loci (SLF, SCR, ACR) are the least; MD has no loci in the PTR. **b**, Gene-set enrichment (g:Profiler; GO, KEGG and Reactome; adjusted *−* log_10_ *P*, colour; dot size, overlapping genes; open circles, not significant) for the annotated genes at FA loci (n = 163 of 273) and for the FA-associated plasma proteins (n = 51). Locus genes are enriched for cell and neuron-projection morphogenesis, actin-cytoskeleton organization, axon guidance and extracellular-matrix organization; plasma proteins for lymphocyte activation, receptor–ligand activity, secretory granules and heparin binding. **c**, Druggability tiers (Open Targets, with ChEMBL-derived clinical phases) of the 657 unique genes at the 898 loci (273, 214 and 460 per measure): approved-drug target (24 genes), clinical-stage target (11), tractable without a reported compound (312) or not currently tractable (310). Numbers above bars give approved plus clinical-stage genes per set (14 FA, 15 MD, 26 AD; 35 in total). **d**, The 35 approved- or clinical-stage targets, grouped by mechanism. For each gene: tier (swatch), measures with a lead variant (FA/MD/AD symbols), tracts in which the footprint is enriched (dark cells; bar chart above gives targets per tract), number of compounds (log bar) and representative drugs with indication. Diamonds mark loci colocalized with a trait (EPHA3, cognitive performance; DPP4, educational attainment). Grey rows are collagens and laminins whose only listed compounds are matrix-degrading enzymes. Gene-level annotations are in Supplementary Table 12.

We then mapped the 315 FA lead variants from the discovery GWAS to 273 genes using FUMA [28], of which 163 had annotations in the tested gene-set databases. Gene-set enrichment with g:Profiler [29] revealed significant over-representation of pathways related to neurogenesis, cyto-plasmic processes, negative chemotaxis, and actin binding (adjusted *P <* 0.001) (Fig. 5b; Supplementary Table 11). The locus genes were most strongly enriched for cell and neuron-projection morphogenesis, actin cytoskeleton organization, axon guidance, extracellular-matrix organization, the glutamatergic synapse and anchoring junctions (Fig. 5b). Many of these pathways play central roles in neural connectivity, axonal remodeling, and cytoskeletal dynamics, biological processes fundamental to white-matter integrity and consistent with earlier large-scale imaging-genetics studies [14, 30]. When enrichment was repeated for the genes at loci assigned to each tract, the superior longitudinal fasciculus (SLF) and anterior corona radiata (ACR) showed the greatest number of enriched pathways (Supplementary Fig. 44), as expected for the tracts that carry the most loci (Fig. 5a).

The plasma proteins associated with FA occupied a complementary functional space (Fig. 5b). Whereas the locus genes were enriched for morphogenetic and cytoskeletal terms, the proteomic features were enriched for lymphocyte activation, receptor–ligand activity, secretory and dense-core granules, heparin binding and blood-vessel morphogenesis, with receptor–ligand activity, heparin binding and anchoring junctions shared between the two sets. To examine functional relationships among implicated genes more directly, we constructed protein–protein interaction networks using STRING v12.0 [31]. The resulting networks showed over-representation of extracellular and signaling-related functions (Supplementary Fig. 45). FA-associated proteins were enriched for protein-binding and signaling-receptor-binding functions (FDR *<* 0.05) and were preferentially localized to extracellular regions and vesicular compartments, including dense-core granules involved in neuropeptide release. These findings highlight an integrated molecular landscape dominated by secreted and receptor-binding proteins, suggesting that intercellular communication, neurovascular interactions, and neuroimmune signaling contribute to the maintenance of white-matter microstructure. Enrichment of secreted and signal-related UniProt keywords, together with blood-expressed proteins, further implies that circulating signaling molecules may influence white matter through peripheral-to-central pathways.

To place the genetic, proteomic and metabolic layers in one network, we constructed a gene–metabolite–disease interaction network using MetaboAnalyst [32] (Supplementary Fig. 46). FA-associated genetic and proteomic features were mapped to known metabolic pathways and linked to significantly associated metabolites through curated gene–metabolite relationships. The resulting network contained interconnected modules involving neuronal maintenance, oxidative stress and lipid metabolism, and Alzheimer’s disease and schizophrenia appeared as nodes connected to multiple genes and metabolites. Because disease annotations in this database are curated from the literature, the network summarizes prior knowledge about the implicated genes rather than providing independent evidence of shared mechanism.

Collectively, these enrichment analyses indicate that the genes at white-matter loci are enriched for cytoskeletal and morphogenetic programs, whereas the circulating proteins associated with white matter are enriched for secreted and receptor-binding functions, with lipid metabolism recurring across layers.

### Pharmacological annotation of white-matter locus genes

To ask whether the genes at the loci identified here are pharmacologically tractable, we assigned genes to the 898 FA, MD and AD loci and annotated each gene with its tractability and known compounds using the Open Targets Platform (release 26.06, which carries ChEMBL-derived clinical phases) [33, 34] and DGIdb [35] (Fig. 5c,d; Supplementary Table 12). Human genetic support increases the probability that a target succeeds in clinical development [36, 37], but positional assignment alone does not establish that a gene is the effector at its locus, so this annotation is a prioritization rather than a causal claim. The three locus sets mapped to 657 unique genes (273 for FA, 214 for MD and 460 for AD, consistent with the FUMA mapping above), of which 61 carry a lead variant in all three measures. By druggability tier, 24 genes (3.7%) are targets of approved drugs, 11 (1.7%) are targets of compounds in clinical development, 312 (47.5%) are tractable by structural or antibody criteria but have no reported compound, and 310 (47.2%) are not currently tractable (Fig. 5c); the proportions were nearly identical within each measure (14, 15 and 26 approved or clinical-stage targets among the FA, MD and AD genes). The 35 genes with an existing compound (Fig. 5d) include 31 at loci whose spatial footprint is enriched in at least one JHU tract (mean 3.5 tracts, median 2), and five (EPHA3, COL4A2, HDAC9, LPAR1 and PRKAG2) carry independent leads in all three diffusion measures with footprints spanning 9–16 tracts.

Footprints of the 35 genes most often involved the superior longitudinal fasciculus (17 genes), superior corona radiata and splenium of the corpus callosum (12 each), anterior corona radiata, body of the corpus callosum and posterior thalamic radiation (11 each), posterior corona radiata (10) and sagittal stratum (9) (Fig. 5d). These are the tracts that carry the most loci overall (Fig. 5a), so the recurrence of druggable-gene footprints in them is expected rather than a property of the druggable subset; 18 of the 35 overlapped at least one of the five tracts in which MR evidence concentrated (SS, ACR, ALIC, IFO, SCR). The corona radiata, including the SCR, and the SLF are also among the tracts most consistently reported with reduced FA in schizophrenia [38] and with MR support for post-traumatic stress disorder and schizophrenia in prior tract-level analyses [39].

Two druggable genes combine footprint breadth, multi-measure support and a shared causal variant with a trait (Fig. 5d), and a third sits beside one. EPHA3 (ephrin type-A receptor 3; vandetanib, KB-004), whose locus spans 12 tracts in all three measures, colocalizes with cognitive performance (PP.H4 = 0.94). DPP4 (dipeptidyl peptidase 4; gliptins) colocalizes with educational attainment (PP.H4 = 0.99). NR1H3 (liver X receptor *α*; LXR agonists in clinical development) is the lipid-sensing receptor at an MD/AD lead (rs10838681) in 11p11.2, 136 kb from and in weak linkage disequilibrium (*r*^2^ = 0.12) with the FA lead whose locus colocalizes with Alzheimer’s disease (above); its own footprint lies in the anterior and superior corona radiata and body of the corpus callosum, and it carries no colocalization of its own, so its link to Alzheimer’s disease is positional only.

Beyond individual genes, the compound-bearing set includes two pathways also implicated by the colocalization and metabolomic results (Fig. 5d). First, lysophosphatidic-acid (LPA) signalling is represented by two unlinked loci: ENPP2 (autotaxin, the enzyme that generates LPA; ziritaxe-stat) at an FA locus spanning seven tracts, and LPAR1 (LPA receptor 1; BMS-986020) at a locus spanning 16 tracts in all three measures. Autotaxin–LPA–LPAR1 signalling regulates oligoden-drocyte maturation and myelination [40], and both compounds were developed for fibrotic disease, so the direction of any effect on white matter is untested. Second, lipid and membrane metabolism is represented by NR1H3 (positionally), SOAT1 (acyl-CoA:cholesterol acyltransferase 1), MGLL (monoacylglycerol lipase) and PRKAG2 (AMP-activated protein kinase *γ*2), echoing the APOE/APOC1 and SREBF1/PEMT colocalizations and the phosphatidylcholine and linoleic-acid associations in the corpus callosum. A third group comprises approved neuropsychiatric drugs whose targets sit on white-matter loci: SLC6A4 (serotonin transporter; selective serotonin reup-take inhibitors and tricyclics, with footprints in ACR, SCR and SLF), SV2A (levetiracetam; SLF), CACNB2 (gabapentinoids; ACR, ALIC, SCR), KCNQ5 (Kv7 openers; IFO, SS) and CALCRL (CGRP-receptor antagonists). MAPT, the target of clinical-stage anti-tau antibodies, also lies at an AD-modality locus, but within the 17q21.31 inversion and without an enriched tract, and is not prioritized here.

Gene assignment was positional and did not use effector-gene evidence; tractability annotations are unadjusted for locus size or gene density; and two steps of the target chain set out in Fig. 1d were not performed: effector-gene assignment and cis-protein quantitative trait locus (cis-pQTL) Mendelian randomization from protein to white matter and to disease. Several approved-drug targets at white-matter loci are oncology kinases (KIT, JAK1, IGF1R), collagens whose only listed compounds are degrading enzymes, or tubulin (TUBB3), and their relevance to microstructure is unclear. The value of the annotation is to list, for each tractable locus gene, the tract, the trait and the compound in one frame for follow-up. The final step of the chain, spatial closure, asks whether the gene acts where the disease acts, and is reported next.

### Spatial closure between locus footprints and disease genetic-correlation maps

Because locus footprints and disease genetic-correlation maps lie on the same 609,000-location grid, we could ask, for each of the 47 colocalized FA loci, whether its footprint (locations at *P <* 9.12 *×* 10*^−^*^12^) coincides with each of the 14 disease maps (locations at *P <* 9.12 *×* 10*^−^*^6^) more than expected under a null that rotates each fiber’s footprint along its own 100 points and permutes fibers within their JHU tract, which preserves footprint size, along-fiber clustering and tract membership (Methods; Supplementary Results §2.8; Supplementary Table 13; Supplementary Fig. 47). The result is largely negative. Of 828 testable locus–map pairs across the three measures, 16 pairs at 13 loci exceeded the null (Benjamini–Hochberg *q <* 0.05), 12 of them at the resolution floor of the null (*P* = 10*^−^*^4^ at 10,000 draws), and the closures concentrated in the genu of the corpus callosum and the corona radiata, where the bipolar disorder, schizophrenia and educational attainment maps overlap one another; we read them as one hotspot rather than as independent results. The test does recover a known relationship: the APOE/APOC1 locus (19q13.32), used as a positive control, has an MD footprint that contains all 105 locations of the Alzheimer’s disease map (16-fold, the maximum attainable for a footprint of this size; *P* = 10*^−^*^4^). Because the Alzheimer’s disease GWAS behind this map includes the APOE region, the locus contributes to the map it is tested against and part of this coincidence is expected by construction; it shows that the test detects a strong known signal rather than validating it independently. Of the 56 locus–trait pairs in which the colocalized trait has a genetic-correlation map, however, only one passed, the 6p21.33 locus in the MHC on the bipolar disorder map (27-fold; *q* = 0.006), and because single-variant colocalization in the MHC is unreliable we do not treat it as a positive finding; none of the compound-bearing colocalized loci (EPHA3, DPP4, SPI1, PDE8B, TNIK) closed on its own trait.

## Discussion

By testing genetic association at 609,000 locations along 6,090 atlas-aligned fiber pathways rather than at the level of whole tracts, this study resolves the spatial footprint of each locus (Fig. 1). Three features of the resulting map are, to our knowledge, new. First, inherited effects on white matter are not whole-tract properties: footprints span five orders of magnitude in size, from single locations to 185,624 locations, and nearly half of the FA loci cross more than two tracts, so that the same tract can host both a focal callosal-segment locus and a brain-wide one. Second, this resolution substantially expands the known genetic architecture of each measure (258, 186 and 298 loci for FA, MD and AD not reported by the largest previous tract-level GWAS [14], and 227 FA loci not linked to any white-matter phenotype in the GWAS Catalog), with 98–99% locus-level replication in a held-out UK Biobank cohort and nominal spatial reproduction of nearly half of the FA loci in children aged 9–10 in ABCD, consistent with part of the localized genetic architecture being in place before adolescence. Third, the regions where loci concentrate are not explained by tract-level heritability: locus density is enriched up to 50-fold in compact capsular and limbic tracts relative to their share of the fiber skeleton, whereas the corpus callosum carries the highest heritability, and the rank correlation between enrichment and *h*^2^ is positive for FA but absent for MD and AD.

Across genetic, proteomic, and metabolomic layers, our results repeatedly point to lipid and membrane metabolism as a recurring biological theme linking white-matter microstructure to systemic and brain disease. Loci that colocalize with cardiovascular, metabolic, and neurodegenerative disease risk (APOE/APOC1 and SREBF1/PEMT, with NR1H3 positionally adjacent to the SPI1 locus) sit within lipid-regulatory pathways, and, independently, the circulating metabolites most strongly associated with white-matter microstructure in our PheWAS are linoleic acid and the other fatty-acid fractions, with phosphatidylcholines among the significant features, all central to lipid and membrane biology. We assessed each of these layers separately (genetic colocalization with disease, and phenotypic association between circulating metabolites and white-matter microstructure), and their co-location on shared anatomy and recurrence in shared pathways nominate lipid and membrane metabolism as a candidate integrative axis rather than a demonstrated one. Testing this directly, for instance whether these same loci associate with circulating levels of the implicated metabolites and whether any such association mediates disease risk, is a natural direction for future genetically informed mediation and colocalization analyses.

The pharmacological annotation extends this axis toward intervention only as a prioritization. About one in twenty genes at white-matter loci already has an approved or clinical-stage compound, and the compound-bearing genes recur in the superior longitudinal fasciculus and corona radiata largely because those tracts carry the most loci. What the annotation adds is a short list in which the tract, the colocalized trait and the compound can be read together, with lipid metabolism and lysophosphatidic-acid signalling (two unlinked loci encoding autotaxin and its receptor LPAR1) recurring in it. The spatial-closure test adds a link that no tract-level study can supply: whether a locus acts where its disease’s genetic correlation with white matter concentrates. Tested against a null that preserves footprint size, along-fiber clustering and tract membership, the answer is largely negative: single-variant footprints are focal whereas polygenic genetic-correlation maps are tract-wide, and the few closures that remain cluster in the genu of the corpus callosum and corona radiata, where several disease maps overlap. The APOE/APOC1 footprint containing the entire Alzheimer’s disease map in MD, built without the APOE region, shows that the test recovers a known relationship; the single own-trait closure, at the MHC locus on the bipolar disorder map, lies in a region where colocalization itself is unreliable and is not counted as a new finding. None of the druggable genes closes on its own trait, so none has yet been shown to act on white matter where a compound would need to move it; establishing effector genes and protein-level MR is the necessary next step, and the map now says, for each candidate, which link is missing.

The spatially organized and reproducible heritability patterns indicate that white-matter microstructure is under stable polygenic control, strongest in the long-range projection and callosal pathways, with loci enriched for axon-guidance, cytoskeletal and extracellular-matrix programs. The polygenic overlap between FA and traits such as Alzheimer’s disease, depression and cognitive performance places white-matter microstructure among the substrates linking genetic risk for neurodegenerative and psychiatric conditions to large-scale communication networks. The sign of the Alzheimer’s disease relationship deserves comment: genetic liability to Alzheimer’s disease was positively correlated with FA, yet manifest Alzheimer’s disease is characterized by reduced FA, and MR with FA as exposure gave no consistent signal in any tract once the APOE region was removed. Possible explanations for the positive correlation include sample overlap with UK Biobank proxy cases in the Alzheimer’s disease GWAS, survival and selection effects in an imaging cohort of healthy middle-aged adults, and age-dependent genetic effects; the present data cannot distinguish these, and we therefore do not interpret the genetic correlation as evidence that higher FA causally increases or decreases Alzheimer’s disease risk.

The overlap of genetic, proteomic and metabolomic associations on the same anatomy provides molecular context for these patterns. Enrichment of lipid-metabolic, cytoskeletal and extracellular-signaling pathways points to membrane maintenance, axonal transport and intercellular communication as candidate molecular determinants of white-matter organization, and the co-location of circulating lipid-metabolite associations and genetic hotspots in the corpus callosum is consistent with models in which neurovascular and neuroimmune pathways couple peripheral metabolic state to central tissue maintenance. These associations are cross-sectional and were not adjusted for body composition or medication, so they describe where systemic and genetic signals coincide rather than how one influences the other.

Several limitations remain. First, the effective resolution of the atlas is set by the 8,868 LDRs rather than by the 609,000 sampling points, and the size and shape of a footprint depend on the fPCA basis and the variance cut-off used to define it; footprints should be read as cluster-level patterns rather than as point-wise localization. Second, the Phase 7 cohort is a held-out split of the same biobank, scanners and ancestry, so replication there guards against sampling error but not against cohort-specific confounding, and the ABCD comparison was limited to FA and used a nominal threshold without an empirical null. Third, most external GWAS for genetic correlation and MR include UK Biobank participants, MR exposures were LDR scores, and reverse-direction MR was not performed, so the MR results are hypothesis-generating. Fourth, the sample consists primarily of middle-aged adults of white British ancestry, and extending analyses across ancestries, developmental periods and aging trajectories will be essential. Fifth, FA, MD and AD are sensitive but not specific indices of microstructure, and the three measures implicated largely distinct locations even where they shared a locus; advanced diffusion models and myelin-sensitive MRI would refine interpretation. Sixth, the multi-omics associations were constrained by modality-specific sample sizes, limited covariate adjustment and the years between blood sampling and imaging, and only the metabolomic layer replicated well. Finally, the pharmacological annotation is positional and unadjusted and nominates genes without establishing effector genes or the direction of drug effect, and the spatial-closure test has limits of its own: the rotation null shifts fibers independently and so may under-represent the clustering of real footprints across neighbouring fibers, making it slightly permissive; its resolution floor (*P* = 10*^−^*^4^ at 10,000 draws) ties the ordering of the strongest pairs; disease maps are available only for the 14 brain-related traits, so loci colocalized with cardiometabolic traits can be tested only against other diseases; and the Alzheimer’s disease map is empty at threshold in FA, so closure on it could be assessed only in MD and AD.

## Methods

### Cohorts and imaging

We analysed diffusion MRI (dMRI) and genotype data from 72,185 UK Biobank (UKB) participants [41, 42] who passed imaging and genetic quality control and were unrelated and of British ancestry by genetic principal components and self-report [43]. Discovery used 56,313 participants from imaging Phases 1–6 and replication an independent 15,872 from Phase 7. The discovery sample was 53.0% female, with a mean age at UK Biobank recruitment of 54.5 years (SD 7.5; range 40–70), and the replication sample 54.3% female, with a mean age at recruitment of 54.5 years (SD 7.6; range 40–70); imaging took place at a later visit. UKB dMRI was acquired on 3T Siemens Skyra scanners at 2 mm isotropic resolution with 50 directions at each of *b* = 1,000 and 2,000 s/mm^2^, and preprocessed centrally (*topup*, *eddy*, gradient-distortion correction) [44]; fractional anisotropy (FA), mean diffusivity (MD) and axial diffusivity (AD) were derived from a tensor fit to the *b* = 1,000 shell. Developmental generalization was tested in the Adolescent Brain Cognitive Development (ABCD) study [45], a multi-site, multi-ancestry cohort of children aged 9–10 years; after quality control 7,403 participants with usable dMRI, genotypes and covariates (47.8% female; mean age 9.9 years, SD 0.6) entered the analysis, which was restricted to FA and processed with an analogous pipeline and tensor fit; association models were adjusted for sex, age, age^2^, age *×* sex, age^2^ *×* sex, 40 genetic principal components and acquisition site (categorical), with leave-one-chromosome-out polygenic predictions to account for relatedness. Full acquisition and preprocessing details are given in Supplementary Methods §1.1.

### Fiber-level atlas and projection

The connectome-based spatial statistics (CBSS) framework [20] maps each participant’s diffusion measures onto a brain function–aware structural connectome atlas. The atlas was built from whole-brain tractography (TractoFlow) of 1,042 Human Connectome Project Young Adult participants, clustered by functional-network-aware Tract Dictionary Learning into 6,090 fiber pathways, each represented by its centroid fiber and labelled by the 12 canonical Cole–Anticevic functional networks and the subcortical system that its endpoints connect. Each participant’s FA image is registered nonlinearly to 1 *×* 1 *×* 1 mm^3^ MNI152 space; a modified tract-based spatial statistics projection [46] assigns the maximum-FA voxel perpendicular to the local tract direction to each skeleton point and extracts MD and AD at the same coordinates; and an elastic registration in the Fisher–Rao square-root slope function framework aligns and refines along-tract profiles across participants. Each pathway carries 100 ordered points, giving 609,000 location-level measurements per participant per measure. For genome-wide testing, two-stage Ward clustering, first by functional network pair and then by centroid proximity, grouped the pathways into 430 clusters of 1–136 fibers (mean 14.2, median 9, interquartile range 4–18). Clusters were assigned to 17 canonical white-matter tracts of the JHU ICBM-DTI-81 atlas [47, 48] by nearest-voxel distance below 10 mm (399 of 430 clusters assigned) and to 66 functional network pairs (Supplementary Methods §1.2). All 6,090 pathways were retained, including the 367 that fall below the test–retest reliability threshold of the released CBSS atlas (5,723 pathways).

### Low-dimensional representations and the effective number of tests

Within each cluster, functional principal component analysis (fPCA) with local linear smoothing [21] compressed the location-level signal into low-dimensional representations (LDRs), with the number of components chosen by an adaptive rule targeting 80–90% of variance (achieved range 70–92%; 8,868 LDRs for FA; Supplementary Table 1). The eigenvalue spectrum defines the effective number of independent locations per cluster, 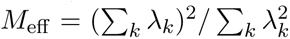, which sums to 5,479.48 for FA (2,946 for MD and 3,895 for AD) and calibrates every location-level multiple-testing correction below.

### Genotypes and GWAS

Imputed genotypes were filtered in PLINK [49, 50] to exclude participants with *>*10% missingness and variants with minor allele frequency *<*0.01, missingness *>*10%, Hardy–Weinberg *P <* 10*^−^*^7^ or imputation INFO *<*0.8, leaving *∼*7.8 million variants, of which *∼*6.8 million non-strand-ambiguous variants were tested. GWAS of every LDR was run with the Representation learning-based Voxel-level Genetic Analysis (RVGA) framework in HEIG [21], adjusting for age, sex, their interactions and squares, 40 genetic principal components and assessment centre, with a two-step ridge regression and leave-one-chromosome-out polygenic offsets to control relatedness and polygenic background [51]. LDR-level summary statistics were projected back to the 609,000 locations through the fPCA bases and LDR covariance, yielding location-level effect sizes and test statistics without a direct voxel-wise GWAS. The same pipeline was run for MD and AD.

### Spatial screening, locus definition and replication

Variants significant at *P <* 5 *×* 10*^−^*^8^*/M*_eff_ (9.12 *×* 10*^−^*^12^ for FA) were reconstructed at location level (*P <* 10*^−^*^5^) and retained when the number of significant locations (the cluster size) exceeded a threshold from an empirical null distribution of cluster size obtained by wild bootstrap [21]: in each of *B* = 50 bootstrap replicates the LDR residuals of every participant were multiplied by an independent standard-normal weight, the LDR GWAS was rerun for *∼*150,000 independent genotyped variants (*r*^2^ *<* 0.1, MAF *>*0.01) and the location-level statistics were reconstructed, giving *∼*7.5 million null cluster sizes at *P <* 10*^−^*^5^; a variant was retained when its cluster size exceeded the 1 *−* 0.05*/*(*L × M*_eff_) quantile of this distribution, where *L* is the number of loci before screening. Because this target quantile (*≈* 1 *−* 10*^−^*^8^) lies beyond the resolution of *∼*7.5 million draws, the realised threshold is the maximum null cluster size. Three-stage LD clumping (cluster-level independent SNPs at *r*^2^ *>* 0.6 within 1 Mb; cluster-level lead SNPs at *r*^2^ *>* 0.1 within 500 kb; merging across clusters) defined tract-level and distinct loci. In Phase 7, a locus was replicated at the locus level when a SNP within *±*250 kb of the discovery lead passed the same screening at *P <* 10*^−^*^4^; the direction of effect was not part of this criterion and was assessed only in aggregate, by regressing replication on discovery effect sizes at the lead variants, and its spatial overlap was the proportion of the discovery footprint (the union of significant locations across SNPs in the window) that was significant in replication, and a locus was counted as spatially replicated when this proportion exceeded 50%. In ABCD (FA only), associations of the discovery candidate variants at *P <* 10*^−^*^2^ were used, without a cluster-size screen. Over-representation of ABCD-reproduced loci in each tract was assessed with Fisher’s exact test against the loci not carried by that tract. Overlap with prior findings was assessed against the loci of Zhao et al. [14] within *±*500 kb and, for FA, against all white-matter entries in the GWAS Catalog within the same window.

### Heritability

RVGA estimated SNP heritability at every location by estimating genetic covariance in LDR space and projecting it through the fPCA bases with analytic standard errors [21]; estimates outside [0, 1] or with inflated standard errors were removed, and significance used a Bonferroni correction over *M*_eff_. Proportions of significant locations use each measure’s own *M*_eff_ over its non-missing locations. Location-level estimates were aggregated to the 17 tracts and 66 network pairs, with ROI-level *P*-values from the aggregated Cauchy association test (ACAT) [52] (Supplementary Table 3). Tract means quoted in the text and in Fig. 2a are means over the significant locations of each tract; Supplementary Table 3 reports means and SDs over all locations of each tract, and Supplementary Fig. 6 medians.

### Genetic correlation, Mendelian randomization and colocalization

Genetic correlations between every LDR and 14 external traits (ADHD, bipolar disorder, major depressive disorder, insomnia, autism, schizophrenia, Alzheimer’s disease, cognitive performance, educational attainment, intelligence, neuroticism, depressed affect, worry and depression; Supplementary Table 2) were estimated with the two-sample framework in HEIG and projected to location level with analytic standard errors. Location-level estimates were aggregated to the 17 tracts and 66 network pairs with two tests computed on all locations of each unit: a signed aggregate, the precision-weighted generalized-least-squares combination of the location-level *r_g_* estimates, in which the correlation among locations within a fiber cluster is reconstructed from the fPCA basis and LDR covariance (shrunk 10% toward the identity) and clusters are treated as independent; and a Cauchy (ACAT) combination of the location-level *P*-values, which is robust to unknown dependence and insensitive to sign. Benjamini–Hochberg FDR (5%) was applied once across all measure *×* trait *×* unit cells of each level (714 tract cells; 2,772 network-pair cells), separately for the two tests, and each cell was classified as consistent-direction (signed test significant), mixed-direction (Cauchy test significant only) or not significant. Because cross-cluster dependence is not modelled, the standard error of the signed aggregate is anti-conservative; it is therefore used only for classification, Supplementary Table 4 reports the mean location-level *r_g_* of each unit as its effect size and gives the Cauchy *q*-value alongside, and the signed aggregate and its working *z* are labelled as such. The GWAS for educational attainment, cognitive performance, intelligence, neuroticism, depressed affect, worry, depression, insomnia and Alzheimer’s disease include UK Biobank participants (Supplementary Table 2), and no correction for sample overlap was applied. Two-sample MR treated each LDR as the exposure, with instruments at *P <* 5 *×* 10*^−^*^8^ in the LDR-level GWAS clumped at *r*^2^ *<* 0.01 within 10 Mb against 1000 Genomes Europeans, harmonized in TwoSampleMR [53] against the same 14 GWAS used for genetic correlation (Supplementary Table 2), taken from the published summary statistics and matched to the instruments by GRCh37 position and alleles (odds ratios were log-transformed; for GWAS reporting only *z*-scores, effects were derived from *z*, sample size and allele frequency; variants in the APOE region, chr19:44.4–46.5 Mb, were removed from the Alzheimer’s disease outcome), and analysed with twelve estimators (fixed-effect IVW, random-effects IVW, radial IVW, MR-Egger [54], simple, weighted and penalised weighted medians [55], simple and weighted modes with and without the NOME assumption, and MR-GRIP). LDR-level results (fits with at least five instruments) were combined to tracts with ACAT for each estimator, FDR-controlled at 5% across the full tract *×* outcome grid for each estimator (17 tracts *×* 14 out-comes; the same flat correction applied to the genetic-correlation cells), filtered for sign consistency with the fixed-effect IVW estimate of the same LDR, and reported when the fixed-effect IVW estimate passed FDR *≤* 0.05 and at least three estimators in total, fixed-effect IVW included, were FDR-significant and sign-concordant (68 pairs; Supplementary Table 9). In all 68 pairs the three concordant estimators are the fixed-effect, random-effects and radial IVW estimators, which are close to concordant by construction; no other estimator met the sign-concordance criterion in any cell of the grid, so the reported pairs are in effect those in which the random-effects IVW estimate is also FDR-significant (Supplementary Methods §1.11). The sign of an LDR is arbitrary and no sign alignment was applied. Reverse-direction MR was not performed. Colocalization used *coloc.abf* [56] with default priors at the representative location of each FA locus (minimum *P* with the lead SNP), reconstructing genome-wide statistics at that location and testing them against each of 19 external traits (Supplementary Table 2); analyses were restricted to loci with *>*50% spatial replication, and strong colocalization required PP.H4 *≥* 0.8 and PP.H4 *>* PP.H3 (Supplementary Table 8). The Alzheimer’s disease summary statistics used (GCST90027158 [57]) exclude the APOE region, so colocalization with Alzheimer’s disease could not be assessed at 19q13.32. Genetic correlation and MR used a different Alzheimer’s disease GWAS (Supplementary Table 2), which includes the APOE region.

### Spatial closure

For each of the 47 colocalized FA loci we tested, in each diffusion measure, whether the location-level footprint of the lead variant coincides with each of the 14 disease genetic-correlation maps more than expected. The footprint was the set of locations at the atlas threshold (*P <* 9.12 *×* 10*^−^*^12^) and the disease map the set of locations at *P <* 9.12 *×* 10*^−^*^6^ (0.05*/M*_eff_); locations with a reconstructed *P* of exactly zero (1,833 locations on 19 fibers, identical across variants in the MD and AD maps, and attributable to the aggregation step) were set to missing on both sides before thresholding. The fold enrichment was the observed overlap divided by *n*_footprint_ *× n*_map_*/n*_valid_, and is bounded above by *n*_valid_*/n*_footprint_. Its null distribution was obtained from 10,000 draws in which every fiber’s footprint pattern was circularly shifted along the fiber’s 100 points by an independent random offset and fibers were then permuted within their JHU tract (clusters without a tract assignment were permuted among themselves), which preserves the number of footprint locations, their along-fiber runs and their distribution across tracts; *P* = (1 + #*{*null *≥* observed*}*)*/*(1 + 10,000). A pair was testable when both sets were non-empty in that measure (828 pairs). Benjamini–Hochberg correction was applied across all testable pairs and, separately, across the 56 pairs in which the disease is the trait the locus colocalizes with (Alzheimer’s disease, schizophrenia, bipolar disorder, educational attainment, cognitive performance, intelligence, ADHD, insomnia, major depressive disorder and autism have maps; type 2 diabetes, coronary atherosclerosis, glomerular filtration rate, stroke, liver disease, reaction time and the remaining non-brain traits do not). Footprint definitions by Benjamini–Hochberg FDR over locations, and threshold-free versions using the top 500, 2,000 and 10,000 locations by *−* log_10_ *P* against the same null, are reported as sensitivity columns in Supplementary Table 13; the full results are given in Supplementary Results §2.8.

### Multi-omics association atlases

Plasma proteomics from the UKB Pharma Proteomics Project (*n* = 5,614) [58], Nightingale NMR metabolomics (*n* = 26,412) [59] and clinical laboratory biomarkers (*n* = 18,689) were each regressed against every FA LDR with the GWAS covariates, and the shared fPCA bases reconstructed location-level statistics. Multiplicity used the product of the effective number of features (from the feature correlation eigenspectrum: 25 proteins, 5 metabolomic measures, 13 biomarkers) and *M*_eff_, followed by the same wild-bootstrap spatial-extent screen as the GWAS (Supplementary Table 10). Because the eigenspectrum estimator is dominated by the leading eigenvalue, it can under-count independent tests among highly collinear features, so these thresholds are more permissive than a per-feature Bonferroni correction; this is a limitation of the present analysis. Models were adjusted for the GWAS covariates only. Cross-feature concordance was quantified by Pearson correlation of location-level Z-score maps and by Dice overlap of FDR-thresholded significance masks (Supplementary Methods §1.13–1.14).

### Enrichment and network analyses

FUMA v1.5.7 [28] annotated genome-wide statistics at each locus’s representative location and mapped significant SNPs to genes positionally and through GTEx v8 brain cis-eQTLs (FDR *<* 0.05); proteomic features were mapped to their encoding genes. g:Profiler v0.2.2 [29] tested GO, KEGG and Reactome enrichment with g:SCS correction; STRING v12.0 [31] built protein–protein interaction networks at confidence *>* 0.4; and the MetaboAnalyst 5.0 network module [32] linked genes, HMDB metabolites and disease annotations (Supplementary Table 11). Tract-level locus enrichment used Fisher’s exact tests on lead-SNP tract assignments and a size-normalized enrichment relative to each tract’s fiber count.

### Druggability annotation

Genes at the 898 FA, MD and AD loci were taken from the FUMA mapping above (657 unique genes) and annotated with Open Targets Platform tractability evidence (small-molecule, antibody and PROTAC buckets; release 26.06, queried through the GraphQL API on 2026-09-22) [33], with compounds and their highest clinical phase, mechanism and indication from the ChEMBL-derived drug records in the same release [34], and with drug–gene interactions from DGIdb (export of 202609-22) [35]. Druggability tiers (Supplementary Table 12) follow Open Targets; DGIdb interactions are reported separately, because DGIdb lists compounds for some genes that Open Targets does not tier as tractable (for example PDE8B and TNIK). Genes were placed in four tiers: approved-drug target, clinical-stage target, tractable without a reported compound, and not currently tractable (Supplementary Table 12). For the 35 genes in the first two tiers, the tracts in which the locus footprint was enriched were taken from the tract-level locus assignments, and recurrence across genes was counted per tract (Fig. 5c,d). Tract–disease literature links were curated from ENIGMA case–control dMRI meta-analyses [38] and tract-level MR studies [39]. No adjustment was made for locus size or gene density, and no effector-gene prioritization or cis-pQTL MR was performed; the annotation is intended as a prioritization.

### Statistics and reproducibility

All tests were two-sided. Location-level GWAS significance used *P <* 5 *×* 10*^−^*^8^*/M*_eff_ and location-level heritability and genetic-correlation significance *P <* 0.05*/M*_eff_, with *M*_eff_ the effective number of independent locations per measure (5,479, 2,946 and 3,895); tract- and network-pair-level tests, Mendelian randomization and feature-pair overlaps were controlled at a Benjamini–Hochberg false-discovery rate of 5% across the full grid of tests stated in each section; spatial extents were screened against wild-bootstrap null distributions; and spatial closure used a 10,000-draw rotation null. Sample sizes are 56,313 (discovery), 15,872 (replication) and 7,403 (ABCD) participants, and 5,614, 26,412 and 18,689 for proteomics, metabolomics and clinical biomarkers; no data were excluded after quality control and no randomization or blinding applied to this observational study.

## Data Availability

The UK Biobank data analyzed in this study are available from the UK Biobank (https://www.ukbiobank.ac.uk/) upon application and approval (Application 22783). The Adolescent Brain Cognitive Development (ABCD) Study data are available through the NIMH Data Archive (https://nda.nih.gov/abcd) upon approval by the NIH Data Access Committee. The derived summary statistics and analysis code generated for this study will be made available from the corresponding author upon reasonable request.

https://github.com/aereinh/CBSS_GWAS

## Ethics

UK Biobank has ethical approval from the North West Multi-centre Research Ethics Committee and participants gave written informed consent; data were accessed under application 22783. ABCD procedures were approved by the study’s centralized and site institutional review boards with parental consent and child assent.

## Data availability

Location-level summary statistics for FA, MD and AD, heritability and genetic-correlation maps, the locus tables and the druggability annotation are available through the browsable atlas at https://bigkp.org. UK Biobank data are available to approved researchers through https://www.ukbiobank.ac.uk, and ABCD data through the NIH Brain Development Cohorts (NBDC) Data Sharing Platform (formerly distributed through the NIMH Data Archive). External GWAS summary statistics are listed in Supplementary Table 2.

## Code availability

Analysis code is available at https://github.com/aereinh/CBSS_GWAS; the HEIG software is available at https://github.com/Zhiwen-Owen-Jiang/heig/.

## Acknowledgements

We thank research assistants in the UNC BIG-S2 team for downloading and preprocessing raw images. We thank the individuals represented in the UK Biobank study for their participation and the research teams for their work in collecting, processing, and disseminating these datasets for analysis. We thank University of North Carolina at Chapel Hill and the Research Computing groups for providing computational resources and support that have contributed to the research results. This research has been conducted using the UK Biobank resource (application number 22783), subject to a data transfer agreement. Data used in the preparation of this article were obtained from the Adolescent Brain Cognitive Development^TM^ (ABCD) Study (https://abcdstudy.org/), held in the NIH Brain Development Cohorts Data Sharing Platform (https://www.nbdc-datahub.org/). This is a multisite, longitudinal study designed to recruit more than 10,000 children aged 9–10 and follow them over 10 years into early adulthood. The ABCD Study^®^is supported by the National Institutes of Health and additional federal partners under award numbers: U01DA041048, U01DA050989, U01DA051016, U01DA041022, U01DA051018, U01DA051037, U01DA050987, U01DA041174, U01DA041106, U01DA041117, U01DA041028, U01DA041134, U01DA050988, U01DA051039, U01DA041156, U01DA041025, U01DA041120, U01DA051038, U01DA041148, U01DA041093, U01DA041089, U24DA041123, U24DA041147. A full list of supporters is available at https://abcdstudy.org/federal-partners.html. ABCD Consortium investigators designed and implemented the study and/or provided data but did not necessarily participate in the analysis or writing of this report. This manuscript reflects the views of the authors and may not reflect the opinions or views of the NIH or ABCD Consortium investigators. The ABCD data repository grows and changes over time. The ABCD genotype data used in this report came from NIMH Data Archive Digital Object Identifier (DOI) 10.15154/1504431 (Data Release 2.0.1); raw baseline diffusion MRI data were obtained from the ABCD imaging data shared in NIMH Data Archive collection 2573.

## Author contributions

A.R., Z.J. and H.Z. conceived and designed the study. Z.J. developed the statistical methodology. A.R., Z.J. and T.L. developed the analytical software. T.L. constructed the white-matter atlas. A.R., T.L., B.T., X.Q. and S.H. performed data preprocessing. A.R. conducted the primary analyses, with contributions from Z.J. A.R. generated the figures and drafted the manuscript, with contributions from T.L. A.R., Z.J., T.L., J.G.I., Y.L., B.Z., P.S., J.S. and H.Z. contributed to the interpretation of results. J.G.I. and H.Z. supervised the study. All authors reviewed and revised the manuscript and approved the final version.

## Additional information

Supplementary Information is available for this paper. Correspondence and requests for materials should be addressed to Hongtu Zhu.

