## Supplementary Information for "A spatially resolved genomic and molecular atlas of human white matter"

**3 References** **28**

**4 Supplementary Figures** **33**
Supplementary Fig. 1: CBSS white-matter atlas used for fiber-level analyses 33
Supplementary Fig. 2: Properties of low-dimensional representations (LDRs) 34
Supplementary Fig. 3: Quality control and calibration of voxel-level GWAS 35
Supplementary Fig. 4: Voxel-wise heritability across diffusion MRI measures 36
Supplementary Fig. 33: Colocalization at 6p25.1 (Glomerular filtration rate) 65
Supplementary Fig. 34: Colocalization at 19q13.32 (Coronary atherosclerosis) 66
Supplementary Fig. 35: Voxel GWAS association map at rs9262135 (6p21) . 67
Supplementary Fig. 36: Voxel GWAS association map at rs6062264 (20q13.12) 68
Supplementary Fig. 37: Voxel GWAS association map at rs1747223 (13q14.11) 69
Supplementary Fig. 38: Voxel GWAS association map at rs2306811 (18p11.21) 70
Supplementary Fig. 39: Voxel GWAS association map at rs78903072 (17q21.31) 71

Supplementary Fig. 40: Cross-omics phenotypic and spatial correlation maps 72
Supplementary Fig. 41: Cross-omics top features associated with FA . . . . 73
Supplementary Fig. 42: Cross-omics top features associated with MD . . . . 74
Supplementary Fig. 43: Cross-omics top features associated with AD . . . . 75
Supplementary Fig. 44: Tract-level gene-set enrichment of FA locus genes . 76
Supplementary Fig. 45: STRING protein–protein interaction network of FA-associated plasma proteins . . . . . . . . . . . . . . . . . . . . . . . . . 77
Supplementary Fig. 46: MetaboAnalyst gene–metabolite–disease network . . 78
Supplementary Fig. 47: Spatial closure of locus footprints on disease genetic-correlation maps . . . . . . . . . . . . . . . . . . . . . . . . . . . . . . 79
Supplementary Fig. 48: ADGRG1 (proteomics) . . . . . . . . . . . . . . . . 80
Supplementary Fig. 49: BAIAP2 (proteomics) . . . . . . . . . . . . . . . . . 81
Supplementary Fig. 50: MOCS2 (proteomics) . . . . . . . . . . . . . . . . . 82
Supplementary Fig. 51: NPY (proteomics) . . . . . . . . . . . . . . . . . . . 83
Supplementary Fig. 52: SYT1 (proteomics) . . . . . . . . . . . . . . . . . . 84
Supplementary Fig. 53: Ratio of saturated fatty acids to total fatty acids (metabolomics) . . . . . . . . . . . . . . . . . . . . . . . . . . . . . . 85
Supplementary Fig. 54: Lactate (metabolomics) . . . . . . . . . . . . . . . . 86
Supplementary Fig. 55: Cholesteryl esters to total lipids ratio in very large VLDL (metabolomics) . . . . . . . . . . . . . . . . . . . . . . . . . . 87
Supplementary Fig. 56: Phospholipids to total lipids ratio in large HDL (metabolomics) . . . . . . . . . . . . . . . . . . . . . . . . . . . . . . 88
Supplementary Fig. 57: Cholesteryl esters to total lipids ratio in medium HDL (metabolomics) . . . . . . . . . . . . . . . . . . . . . . . . . . . 89
Supplementary Fig. 58: Calcium, albumin-adjusted (biomarkers) . . . . . . 90
Supplementary Fig. 59: Iron (biomarkers) . . . . . . . . . . . . . . . . . . . 91
Supplementary Fig. 60: Free thyroxine, FT4 (biomarkers) . . . . . . . . . . 92
Supplementary Fig. 61: Lymphocyte count (biomarkers) . . . . . . . . . . . 93
Supplementary Fig. 62: Red blood cell count (biomarkers) . . . . . . . . . . 94

### 1 Materials and Methods

This section follows the analysis pipeline in order. Sections 1.1–1.2 describe the cohorts and the CBSS pipeline that converts diffusion images into 609,000 location-level measurements on a common atlas. Section 1.3 compresses these locations into 8,868 low-dimensional representations (LDRs) and derives the effective number of independent locations that calibrates every multiple-testing correction below. Sections 1.4–1.8 cover the genetic data and the core GWAS machinery: quality control, LDR-level association testing, reconstruction of location-level effects, spatial screening and locus definition, and replication; Section 1.9 estimates heritability. Sections 1.10–1.12 then relate white-matter genetic architecture to external traits through three complementary lenses: shared polygenic architecture (genetic correlation), putative directional effects (Mendelian randomization), and shared causal variants (colocalization). Sections 1.13–1.15 add the measured molecular layers and their integration, Section 1.16 annotates locus genes for pharmacological tractability and tests the spatial closure between locus footprints and disease genetic-correlation maps, Section 1.17 extends all analyses to the MD and AD modalities, and Section 1.18 reports sensitivity analyses.

#### 1.1 Study cohorts and imaging data

This study draws on diffusion magnetic resonance imaging (dMRI) and genetic data from the UK Biobank (UKB), a population-based cohort of more than 500,000 participants aged 40–69 years at recruitment [60]. The UKB imaging substudy acquired multimodal brain MRI under standardized protocols across multiple assessment centers [61]. The primary analyses include 72,185 individuals of European ancestry from UKB Phases 1–7 who passed imaging and genetic quality control. To minimize population stratification and ensure homogeneity, we restricted the sample to unrelated individuals of British ancestry, defined by UKB genetic principal components and self-reported ethnicity [62], and we excluded participants with missing imaging data, poor image quality, or failed genetic quality control. Discovery and replication use non-overlapping subsets of the UKB imaging cohort: 56,313 individuals from Phases 1–6 for discovery and 15,872 individuals from Phase 7 for replication, so the replication cohort tests every association in an independent sample.

A standardized protocol on 3T Siemens Skyra scanners with a 32-channel head coil acquired the diffusion data at $2 \times 2 \times 2$ mm$^3$ spatial resolution with anterior-to-posterior phase encoding and a multiband acceleration factor of three, using a monopolar Stejskal–Tanner pulse sequence (echo time 92 ms, repetition time 3,600 ms) with 50 diffusion-encoding directions at each of two nonzero b-values ($b = 1{,}000$ and 2,000 s/mm$^2$) [61]. The UKB brain-imaging team preprocessed the dMRI data centrally [63]: the *topup* tool estimates the $B_0$ fieldmap from the highest-quality pair of opposite-phase-encoded $b = 0$ volumes, the *eddy* tool corrects eddy-current distortions, head motion, and outlier slices, and gradient-distortion correction yields the final corrected 4D data; an automated classifier scores every T1-weighted image and removes unusable scans together with their companion dMRI. Diffusion tensor models fitted to the $b = 1{,}000$ shell yield the fractional anisotropy (FA), mean diffusivity

(MD), and axial diffusivity (AD) maps analyzed here. The connectome-based spatial statistics (CBSS) pipeline [64] projects these measures onto a white-matter skeleton and builds a high-resolution representation of fiber architecture across subjects. An analogous pipeline processed the Adolescent Brain Cognitive Development (ABCD) study, the external developmental cohort used in this work [65].

The ABCD study, used here to test developmental generalization of the identified loci, is a longitudinal cohort of 11,878 children aged 9–10 years at baseline, recruited at 21 sites across the United States [65]. Its dMRI protocol is harmonized across 3T Siemens Prisma, GE MR750, and Philips scanners: a multi-shell acquisition at 1.7 mm isotropic resolution with multiband acceleration, comprising 96 diffusion-encoding directions distributed over four shells ($b$ = 500, 1,000, 2,000, and 3,000 s/mm$^2$), with reverse phase-encoded $b = 0$ volumes acquired for susceptibility-distortion correction [65]. All ABCD procedures were approved by the study's centralized and site-level institutional review boards; parents or guardians provided written informed consent and children provided assent. Diffusion tensor maps for the present analyses were derived with processing analogous to the UKB pipeline and mapped onto the BFSC atlas through the same CBSS projection and elastic-registration stage described in §1.2. As with UKB, the ABCD tensor model was fitted to the $b = 1{,}000$ s/mm$^2$ shell. After quality control, 7,403 participants with usable diffusion data, genotypes and covariates entered the ABCD analysis, which was restricted to FA; the sample was multi-ancestry and unrestricted by genetic ancestry, consistent with the developmental-generalization aim of this validation cohort. The ABCD association models were adjusted for sex, age, age$^2$, age $\times$ sex, age$^2$ $\times$ sex, the first 40 genetic principal components and acquisition site as a categorical covariate, with leave-one-chromosome-out polygenic predictions to account for relatedness.

#### 1.2 The CBSS pipeline: atlas construction, projection, and preprocessing

The CBSS framework operates in three stages [64]: (i) whole-brain tractography and alignment to standard space, (ii) construction of a brain function–aware structural connectome (BFSC) atlas, and (iii) atlas-based mapping of diffusion measures from new cohorts onto the atlas. The first two stages were completed once, in the atlas-construction cohort; only the third stage runs on UK Biobank and ABCD data in the present study.

The BFSC atlas derives from 1,042 Human Connectome Project–Young Adult participants with high-quality dMRI and structural MRI [64]. TractoFlow generates whole-brain tractography for each subject, and streamline endpoints anchor to a 359-region parcellation built from the HCP-MMP cortical atlas and 13 FreeSurfer subcortical regions, with adjacent cortical parcels merged when they belong to the same functional network, correlate above 0.7 in their fiber-count connectivity profiles, and do not reduce test–retest reliability (assessed in 43 pairs of HCP retest scans). The parcels organize into 13 functional networks (12 canonical networks plus a subcortical network) following the Cole–Anticevic Brain-wide Network Partition. After aggregating subject-level tractography into a population-scale tractogram in MNI152 space, functional-network–aware Tract Dictionary Learning clusters the streamlines into 6,090 pathway clusters; each cluster's centroid fiber, defined as the streamline with the smallest median minimum-direct-flip distance to all other members, represents the pathway, and post-clustering quality control (removing small clusters and centroids with FA test–retest reliability below 0.4) retains 5,723 centroids in the released atlas.

We retained all 6,090 fiber clusters in the genetic analyses rather than the 5,723 that survive the reliability filter of [64], because for GWAS low phenotypic reproducibility primarily attenuates genetic effect sizes rather than generating false-positive associations, and the large UKB sample compensates for the weaker signal; the stricter thresholds of [64] reflect the stability requirements of normative growth-chart modeling, where sparse or poorly reproducible pathways yield unstable age-trajectory estimates. Each fiber pathway carries 100 ordered voxels, giving 609,000 voxel-level measurements per subject.

In the mapping stage, CBSS transfers diffusion measures from each new cohort onto the atlas without subject-level whole-brain tractography. Each subject's FA image registers nonlinearly to $1 \times 1 \times 1$ mm$^3$ MNI152 space [66], and a modified tract-based spatial statistics (TBSS) projection assigns values to the atlas skeleton: for every skeleton voxel, the procedure searches perpendicular to the local tract direction for the maximum FA value in the subject's image, records that voxel as the tract center, and extracts all other diffusion statistics (MD, AD) at the same coordinates, with trilinear interpolation at 0.1 mm$^3$ resolution preserving sub-voxel detail. Because corresponding along-tract profiles can still differ by local stretching or compression after volumetric registration, a cross-subject elastic registration step then aligns streamline-wise diffusion profiles using the Fisher–Rao square-root slope function framework: each profile warps toward a cross-subject Karcher mean template through a smooth monotone warping function, regularized toward the identity and scaled by streamline length, yielding reparameterization-invariant correspondence of profiles across subjects [64].

To reduce computational burden and improve statistical efficiency, Ward's minimum-variance clustering groups the fibers into 430 clusters in two stages: fibers first group by functional network pair (as defined within the CBSS framework), and within each pair agglomerative clustering merges fibers whose centroids in MNI152 space lie within 30 mm of one another, preserving both anatomical proximity and functional coherence. The final partition comprises clusters of 1–136 fibers (mean 14.2, median 9, interquartile range 4–18), with the largest clusters concentrated in major projection and callosal pathways.

For downstream interpretation, we assigned each cluster to canonical white-matter regions of interest from the JHU ICBM-DTI-81 atlas [67]: the mean Euclidean distance between a cluster's fiber voxels and the binary mask of each of 26 atlas-defined tract regions (nearest-voxel approach) determines assignment to the closest tract whenever that distance falls below 10 mm. This rule assigned 399 of the 430 clusters to one of 17 major white-matter tracts, leaving the remainder unassigned. We chose the 10 mm threshold to balance anatomical specificity against coverage: thresholds below 8 mm left more than 20% of clusters unassigned, while thresholds above 12 mm produced ambiguous assignments where tracts closely adjoin, as in the corona radiata and internal capsule. Merging bilateral homologs (e.g., left and right corticospinal tract) consolidates the 26 JHU regions into 17 canonical tracts (the genu, body, and splenium of the corpus callosum remain distinct). The 31 unassigned clusters, primarily located at tract intersections or in deep white matter poorly covered by the JHU atlas, remain in all GWAS and heritability analyses but drop out of tract-level aggregation and visualization. All clusters also carry annotations for 66 functional network pairs (corresponding to 13 unique functional networks) as defined within the CBSS framework [68], two fewer than the 68 pairs spanned by the full CBSS atlas. This gap arises because none of the 6,090 fibers of the atlas used here connects the remaining two pairs; no minimum cluster size was imposed, and the 430 clusters contain 1–136 fibers each (§1.2).

#### 1.3 Functional PCA and low-dimensional representations

The CBSS mapping yields 609,000 spatially correlated voxel-level measurements per subject, too many and too redundant for direct genome-wide testing. Within each fiber cluster, functional principal component analysis (fPCA) therefore compresses the voxel-level FA signal into low-dimensional representations (LDRs) [68, 69]. A local linear smoothing approach [70] estimates the functional basis functions from UKB Phase 1–6 data; projecting each subject's cluster-specific FA signal onto these bases yields the LDR scores.

Because fiber clusters vary in size and spatial covariance structure, an adaptive rule selects the number of retained components per cluster, targeting 80–90% of total variance and a reconstruction fidelity (correlation between original and reconstructed signals) of 0.85–0.95; the achieved values were 70–92% and 0.83–0.99 (mean 0.94; Supplementary Fig. 2D; Supplementary Table 1). The resulting eigenvalues summarize the variance each component explains and characterize the intrinsic dimensionality of voxel-level variation within each cluster (Supplementary Table 1).

To account for spatial correlation in downstream inference, the eigenvalue spectrum yields the effective number of independent voxels,

$$M_{\text{eff}} = \left( \sum_{k=1}^{K} \lambda_k \right)^2 \Bigg/ \sum_{k=1}^{K} \lambda_k^2,$$

which quantifies the degree of redundancy among voxel measurements. Summing across clusters gives approximately 5,479 effective independent voxels across the white-matter skeleton. This quantity calibrates multiple-testing correction and significance thresholds in all subsequent voxel-level analyses. Supplementary Table 1 summarizes cluster-level fPCA characteristics.

#### 1.4 Genotype data and quality control

On the genetic side, participants were genotyped on the UK Biobank Axiom and UK BiLEVE arrays, and the UKB team imputed genotypes centrally against the Haplotype Reference Consortium and UK10K + 1000 Genomes reference panels [62]; we obtained the released genotyped and imputed variant data. We applied standard

GWAS quality-control filters [71, 72], implemented in PLINK [73, 74], to every individual with both imaging and genetic data: we excluded individuals with more than 10% missing genotypes and removed variants with minor allele frequency below 0.01, missing genotype rate above 10%, or deviation from Hardy–Weinberg equilibrium at $P < 1 \times 10^{-7}$; for imputed variants we additionally required an imputation INFO score above 0.8, consistent with standard practice in large-scale GWAS [72].

These filters left approximately 7.8 million variants; removing strand-ambiguous variants retained 6.8 million for downstream genome-wide association analyses, alongside approximately 460,000 directly genotyped SNPs from the unimputed dataset. A comparable pipeline processed the ABCD cohort.

#### 1.5 GWAS of low-dimensional representations

With the 8,868 LDR phenotypes (§1.3) and quality-controlled genotypes (§1.4) in hand, the Representation learning-based Voxel-level Genetic Analysis (RVGA) framework, implemented in the Highly Efficient Imaging Genetics (HEIG) tool [68], ran the genome-wide association studies (GWAS). The discovery analysis covered 56,313 individuals of British ancestry from UKB Phases 1–6, and replication used an independent cohort of 15,872 individuals from UKB Phase 7.

For each LDR, linear regression tested the association between each genetic variant and the LDR values [73], adjusting for age, sex, age$\times$sex, age$^2$, age$^2\times$sex, 40 genetic principal components, and assessment center. A two-step ridge regression procedure [75] controls sample relatedness and polygenic background: ridge models fitted within approximately 100 blocks of $\sim$5,000 genotyped variants predict each LDR, and a leave-one-chromosome-out (LOCO) framework [76] combines the block predictions into chromosome-specific polygenic scores that enter each GWAS as offsets.

Five-fold cross-validation selects the ridge shrinkage parameter, reflecting the contribution of polygenic effects to each LDR. This design improves statistical calibration and power while maintaining computational scalability across high-dimensional imaging phenotypes [68].

#### 1.6 Voxel-level reconstruction of GWAS effects

Because the GWAS operates in the reduced LDR space while inference targets anatomical locations, RVGA maps the LDR-level results back to voxel space by integrating LDR summary statistics with the corresponding fPCA basis functions and LDR variance–covariance matrices [68]: for each variant, the fPCA loadings weight the LDR-level effect estimates, and their linear combination yields voxel-level effect sizes and test statistics, enabling spatially resolved inference across the white-matter skeleton.

The reconstruction preserves the covariance structure that dimensionality reduction induces and recovers high-resolution genetic association patterns without performing voxel-wise GWAS directly.

#### 1.7 Spatial post-screening and locus definition

The reconstructed maps of §1.6 contain many nominally significant voxels; a spatial post-screening step therefore prioritizes robust, spatially coherent associations before loci are defined. The screen is based on cluster size, the number of voxels at which a variant is associated at $P < 1 \times 10^{-5}$, under the rationale that a genuine association should not be confined to a handful of voxels given the strong spatial correlation of the maps [68].

We first identified unique significant SNPs in the discovery cohort at a genome-wide threshold of $5 \times 10^{-8}$ [77] divided by the total effective number of independent voxels ($M_{\text{eff}} = 5479.48$), and reconstructed their voxel-level associations at $P < 1 \times 10^{-5}$. An empirical null distribution of cluster size was then obtained by wild bootstrap (Algorithm 1 of 68; `--cluster` in HEIG): in each of $B = 50$ bootstrap replicates, the LDR residuals of every participant were multiplied by an independent standard-normal weight, the LDR GWAS was rerun for approximately 150,000 independent genotyped variants ($r^2 < 0.1$ within 50-variant windows, MAF $> 0.01$), the voxel-level statistics were reconstructed through the same bases and LDR covariance, and the number of voxels passing $P < 1 \times 10^{-5}$ was recorded for every variant, giving approximately 7.5 million null cluster sizes. A SNP survived screening when its cluster size exceeded the $1 - 0.05/(L \times M_{\text{eff}})$ quantile of this null distribution, where $L$ is the number of loci obtained by aggregating all voxel–variant associations before screening. With $L$ in the hundreds and $M_{\text{eff}} = 5{,}479$, this target quantile ($\approx 1 - 10^{-8}$) lies beyond the resolution of the $\sim$7.5 million null draws, so the realised threshold is the maximum null cluster size. Using $P < 1 \times 10^{-5}$ rather than the genome-wide threshold to define cluster size keeps the null distribution away from zero; variants that pass at $1 \times 10^{-5}$ also pass at the stricter threshold, whereas a variant that is extremely significant at only a few voxels is treated as a local perturbation and excluded. This procedure yields variants that are both genetically significant and spatially enriched. A three-stage linkage disequilibrium (LD) clumping procedure [73] then defines independent loci: (i) cluster-level independent significant SNPs at $r^2 > 0.6$ within a 1 Mb window; (ii) cluster-level lead SNPs by further clumping at $r^2 > 0.1$ within 500 kb; and (iii) tract-level lead SNPs by merging signals across clusters at the same LD threshold.

#### 1.8 Replication framework

Every locus defined in §1.7 then faces replication at the tract level in the independent UKB Phase 7 data. For each tract-level lead SNP identified in discovery, the pipeline searches a ±250 kb window in the replication cohort; a locus counts as replicated at the locus level when a corresponding SNP in that window passes the post-screening criteria. The direction of effect is not part of this criterion; it was assessed in aggregate by regressing replication on discovery effect sizes at the lead variants (main text).

Spatial replication compares voxel-level association patterns between discovery and replication: for each locus, the union of voxels across all significant SNPs within the ±250 kb region defines the discovery footprint, and the proportion of those voxels that reach significance in replication quantifies spatial concordance.

In the UKB Phase 7 replication cohort, RVGA reconstructs and evaluates voxel-level associations at $P < 1 \times 10^{-4}$, following the moderately relaxed thresholds common in follow-up analyses of high-dimensional neuroimaging phenotypes [78, 79]. External validation in the ABCD dataset, carried out for FA, uses a more permissive threshold of $P < 1 \times 10^{-2}$ on the discovery candidate variants to accommodate its reduced sample size and cohort differences. In Phase 7, bootstrap-based null distributions determine the cluster-size thresholds [68]; no cluster-size screen was applied in ABCD. Summary metrics count loci exhibiting any spatial overlap ($> 0\%$) and substantial overlap ($> 50\%$ of the discovery footprint, the criterion for spatial replication throughout) between discovery and replication.

#### 1.9 Heritability estimation

The same LDR-to-voxel projection that underlies the association analyses also supports variance-component estimation. The RVGA estimator computes voxel-level SNP heritability efficiently for high-dimensional imaging phenotypes: it estimates genetic covariance in the LDR space, projects it to the voxel level through the corresponding functional basis functions, and derives analytic standard errors following [68]. Quality control removes voxel-level estimates with out-of-bounds values (negative or exceeding 1) or inflated standard errors. A Bonferroni correction over the total effective number of independent voxels determines statistical significance ($M_{\text{eff}} = 5479.48$; $P < 0.05/M_{\text{eff}}$).

To facilitate anatomical interpretation, the voxel-wise estimates aggregate to the ROI level for 17 major white-matter tracts and 66 functional network pairs. For each ROI we report the mean heritability, the proportion of significant voxels, and an ROI-level p-value from the aggregated Cauchy association test (ACAT; 80) (Supplementary Table 3), which provides valid inference under arbitrary correlation among input statistics.

#### 1.10 Genetic correlation analysis

Sections 1.10–1.12 relate the genetic architecture of white matter to external traits through three complementary questions: whether polygenic architecture is shared (genetic correlation), whether there is evidence for directional relationships (Mendelian randomization), and whether individual loci share causal variants (colocalization). First, we estimated voxel-level genetic correlations between white-matter microstructure and 14 external phenotypes using publicly available GWAS summary statistics (Supplementary Table 2): attention-deficit/hyperactivity disorder (ADHD; 81), bipolar disorder [82], major depressive disorder [83], insomnia [84], autism spectrum disorder (ASD; 85), schizophrenia [86], Alzheimer's disease [87], cognitive performance and educational attainment [88], intelligence [89], and neuroticism with its subclusters (depressed affect and worry) plus depression from a large-scale neuroticism GWAS [90].

The two-sample framework implemented in HEIG [68] combines the LDR summary statistics with each external trait's summary statistics to estimate genetic correlations in the LDR space: the pipeline aligns alleles to reference LD panels and constructs LD matrices and their inverses at regularization levels of 90% and 85%, respectively, for numerical stability. The fPCA basis functions then project the LDR-level estimates to voxel space with analytic standard errors, yielding spatial maps of shared genetic architecture. Voxel-level estimates were aggregated to the 17 JHU tracts and the 66 functional network pairs with two complementary tests computed on all voxels of each unit (Supplementary Table 4). The signed test is a precision-weighted generalized-least-squares combination of the voxel-level $r_g$ estimates in which the correlation among voxels within a fiber cluster is reconstructed from the fPCA basis and the LDR covariance (normalized to a correlation matrix and shrunk 10% toward the identity) and fiber clusters are treated as independent; it yields a signed aggregate with a working standard error, $z$ and $P$-value, and asks whether the correlation is consistently of one sign across the unit. The Cauchy (ACAT) combination [80] of the voxel-level $P$-values is robust to unknown dependence and insensitive to sign, and flags units with real but sign-heterogeneous signal. Benjamini–Hochberg FDR (5%) was applied once across all modality $\times$ trait $\times$ unit cells of each aggregation level (714 tract cells; 2,772 network-pair cells), separately for the two tests, and each cell was classified as consistent-direction (signed test significant), mixed-direction (Cauchy test significant only) or not significant. Because dependence between fiber clusters is not modelled, the working standard error of the signed aggregate is anti-conservative; the signed aggregate is therefore used only for classification and is labelled as such in Supplementary Table 4, which reports the plain mean of the voxel-level $r_g$ estimates in each unit as the effect size, together with the Cauchy $q$-value. The GWAS for educational attainment, cognitive performance [88], intelligence [89], neuroticism, depressed affect, worry and depression [90], insomnia [84] and Alzheimer's disease [87] include UK Biobank participants; no correction for sample overlap was applied, and estimates for these traits may be inflated. The schizophrenia, bipolar disorder, major depressive disorder, ADHD and autism GWAS do not include UK Biobank; Supplementary Table 2 records this for every trait.

#### 1.11 Mendelian randomization analysis

Second, two-sample Mendelian randomization (MR) asked whether these shared architectures reflect directional effects, treating white-matter microstructure as the exposure and, as outcomes, the same 14 GWAS used for genetic correlation (§1.10; Supplementary Table 2). Outcome associations were taken from the published summary statistics and matched to the instruments by GRCh37 position and alleles on either strand; odds ratios were converted to log odds ratios and, for GWAS that report only $z$-scores (Alzheimer's disease, depression and neuroticism), effects and standard errors were derived as $\beta = z/\sqrt{2p(1-p)(N+z^2)}$ with $p$ the effect-allele frequency (for Alzheimer's disease, which reports no frequency, the UK Biobank frequency listed in the neuroticism summary statistics), so that those estimates are on a standardized scale. The sign of an LDR score is arbitrary and no sign alignment was applied, so effect directions are not interpreted. Instrument selection retains variants reaching genome-wide significance ($P < 5\times10^{-8}$) in the LDR-level GWAS and prunes them by LD-based clumping ($r^2 < 0.01$, 10 Mb window) against the 1000 Genomes European reference panel to ensure independence. For nine of the outcome GWAS the outcome sample overlaps the exposure sample (§1.10), which biases two-sample MR estimates toward the observational association. The Alzheimer's disease outcome GWAS [87] includes the APOE region; variants in chr19:44.4–46.5 Mb (GRCh37) were therefore removed from the outcome statistics before matching, so that no instrument in the

APOE region enters the Alzheimer's disease analysis. Reverse-direction MR (trait to white matter) was not performed.

The TwoSampleMR package [91] harmonizes exposure and outcome datasets, standardizing allele alignment across traits. Twelve estimators probe robustness to pleiotropy and model assumptions: inverse-variance weighted (IVW) regression under a fixed-effect model and under the TwoSampleMR default random-effects model (in which the standard error is not allowed to fall below its fixed-effect value), radial IVW [92], MR-Egger, simple median, weighted median, penalised weighted median [93], simple mode, simple mode under the no-measurement-error (NOME) assumption, weighted mode, weighted mode (NOME) [94], and MR-GRIP [95].

MR runs at the LDR level and aggregates to the tract level to improve interpretability and statistical power. For each tract–outcome–estimator combination, the aggregated Cauchy association test (ACAT; 80) combines the $P$-values across the LDRs of the clusters assigned to the tract (fits with fewer than five instruments excluded), and the Benjamini–Hochberg procedure controls the false discovery rate at 5% across the full tract $\times$ outcome grid, separately for each estimator (the same flat correction applied to the genetic-correlation cells; the grid comprises 17 tracts $\times$ 14 outcomes). A directional-consistency filter requires the contributing LDR effects to agree in sign with the fixed-effect IVW estimate. We report tract–outcome pairs that (i) pass FDR $\leq 0.05$ under the fixed-effect IVW estimator and (ii) are supported by at least three estimators in total, fixed-effect IVW included, that are FDR-significant and directionally concordant; 68 pairs meet these criteria (Supplementary Table 9, which gives all tract $\times$ outcome $\times$ estimator ACAT $P$- and $q$-values, the number of contributing clusters and the number of instruments per pair). The fixed-effect and random-effects IVW estimators share one point estimate and radial IVW is a reweighted form of the same regression, so the three IVW estimators are close to concordant by construction: in all 68 pairs they are the three concordant estimators. At least one non-IVW estimator is FDR-significant in 55 of the 68 pairs, but none passes the directional-consistency filter in any cell of the grid; across the 68 pairs a median of 5.5 of the 12 estimators are FDR-significant (range 3–10). The reported pairs therefore rest on IVW, and Supplementary Table 9 lists, for each pair, the number of estimators of any kind that are FDR-significant. Radial IVW could not be computed for 2,226 LDR–outcome fits (1.8% of the radial IVW fits); the other eleven estimators of those fits were retained. Heterogeneity statistics, MR-Egger intercepts and Steiger directionality tests were not retained from the LDR-level fits, and reverse-direction MR was not performed.

#### 1.12 Colocalization analysis

Third, moving from genome-wide summaries to individual loci, Bayesian colocalization implemented in the *coloc* R package [96] tested whether white-matter microstructure and external traits share common causal variants at each discovery locus. For each locus, the voxel with the strongest association (minimum P-value) with the lead SNP serves as the representative voxel; RVGA reconstructs genome-wide association statistics at that voxel, and these merge with GWAS summary statistics for a panel of 19 external traits (Supplementary Table 2): Alzheimer's disease, ADHD, autism spectrum disorder, bipolar disorder, cognitive performance, educational attainment, intelligence, insomnia, major depressive disorder, schizophrenia, Parkinson's disease, reaction time, drinks per week, number of sexual partners, coronary atherosclerosis, stroke, type 2 diabetes, glomerular filtration rate and liver disease after alignment of SNP-wise effect estimates, standard errors, minor allele frequencies, and sample sizes. Summary statistics for ADHD, autism spectrum disorder, bipolar disorder, cognitive performance, educational attainment, intelligence, insomnia, major depressive disorder and schizophrenia were those used for genetic correlation (§1.10); the others were Alzheimer's disease [97], Parkinson's disease [98], reaction time [99], drinks per week and number of sexual partners [100], coronary atherosclerosis and liver disease [101], stroke [102], type 2 diabetes [103] and glomerular filtration rate [104] (Supplementary Table 2).

The approximate Bayes factor method (*coloc.abf*) evaluates five competing hypotheses: no association (H0), association with only the imaging trait (H1), association with only the external trait (H2), two distinct causal variants (H3), or a shared causal variant (H4). The model treats the imaging phenotype (voxel-level FA) as a quantitative trait and specifies each external trait as quantitative or case-control according to its study design, incorporating case prevalence for binary traits. For each locus–trait pair, the posterior probability of the shared causal variant model (PP.H4) quantifies evidence for colocalization (Supplementary Table 8).

To prioritize robust signals, we restricted analyses to the 140 loci exhibiting spatial replication between discovery and replication cohorts ($>50\%$ overlap of associated voxels), and we define strong colocalization as PP.H4 $\geq 0.8$ with PP.H4 $>$ PP.H3, ensuring support for a shared rather than distinct causal signal. Default *coloc* priors were used ($p_1 = p_2 = 10^{-4}$, $p_{12} = 10^{-5}$). Four of the 2,660 locus–trait pairs could not be evaluated (at the 17q21.31 locus, too few shared variants for cognitive performance and educational attainment and a failed fit for reaction time; missing sample sizes for one locus with bipolar disorder) and were treated as not colocalized; they are listed in the notes to Supplementary Table 8. The Alzheimer's disease summary statistics used (GCST90027158; 97) exclude the APOE region, so colocalization with Alzheimer's disease could not be assessed at 19q13.32; the Alzheimer's disease GWAS used for genetic correlation and MR [87] is a different study and includes the APOE region (§1.10–1.11). Summaries rank locus–trait pairs by PP.H4 and spatial overlap, and the visualizations include distributions of PP.H4, heatmaps of locus–trait colocalization, trait-level counts of colocalized loci, and locus-specific regional association plots illustrating concordance between imaging and external traits.

#### 1.13 Multi-omics association analysis

Genetics is only one molecular layer; the next three sections add the measured layers. A framework exactly parallel to the GWAS identifies proteomic, metabolomic, and clinical-biomarker correlates of white-matter microstructure at the voxel level, so that molecular and genetic association maps are directly comparable. Three UK Biobank modalities supply the data: plasma proteomics from the UKB Pharma

Proteomics Project (UKB-PPP; $n = 5{,}614$; 105), nuclear magnetic resonance (NMR)–based metabolomics from Nightingale Health ($n = 26{,}412$), and clinical laboratory measurements from UKB assessment center blood panels ($n = 18{,}689$).

The same fPCA bases and FA LDRs derived from unrelated UKB Phase 1–6 participants (see Functional PCA) carry over. For each modality, linear models regress each LDR on each molecular feature within the subset of subjects with available omics data, adjusting for the same covariates as the GWAS (age, sex, their interactions and squares, 40 genetic principal components and assessment center); body-mass index, lipid-lowering medication, smoking, fasting time, the interval between blood sampling and imaging and, for proteomics, blood-cell counts were not included. The shared fPCA basis functions then reconstruct voxel-level association statistics, making these maps directly comparable with the GWAS-derived atlases.

The multiplicity correction multiplies two effective numbers: the effective number of omics features and the effective number of voxels from the GWAS calibration (5,479.48). The effective number of features was taken as $(\sum \lambda)^2 / \sum \lambda^2$ from the eigenvalue spectrum of the feature–feature correlation matrix (25 of 2,923 proteins, 5 of 502 metabolomic measures and 13 of 57 biomarkers). This estimator is dominated by the leading eigenvalue and can under-count independent tests when features are highly collinear (for the 502 Nightingale measures it implies a per-feature threshold of $0.05/(5 \times 5{,}479) \approx 1.8 \times 10^{-6}$), so the multi-omics thresholds are more permissive than a per-feature Bonferroni correction; this is a limitation of the present analysis. To further control false positives arising from spatial correlation, a spatial-extent screen mirrors the GWAS procedure: 50 wild-bootstrap replicates ($B = 50$), generated by resampling LDR residuals within subjects, build empirical null distributions of cluster sizes (voxels exceeding $P < 1 \times 10^{-5}$), and only voxel clusters exceeding these thresholds count as statistically and spatially significant.

The output is a modality-specific multi-omics association atlas mapping proteomic, metabolomic, and clinical biomarker effects onto white-matter microstructure at voxel-level resolution (Supplementary Table 10).

#### 1.14 Cross-omics spatial concordance

With genetic and molecular association maps expressed on the same 609,000 locations, two similarity measures quantify their spatial correspondence across features and omics layers, computed from the voxel-wise Z-score maps reconstructed in §1.13. Pearson correlation coefficients ($r$) across the 609,000 voxels of the white-matter skeleton quantify pairwise spatial similarity for every feature pair, both within and across modalities; mean absolute correlations ($|r|$) summarize concordance independent of effect direction.

Dice coefficients complement the correlations: after thresholding the voxel-wise maps at a false discovery rate (FDR) of 0.05 using the joint effective-number correction described above, binary significance masks for each feature yield

$$\text{Dice} = \frac{2|A \cap B|}{|A| + |B|},$$

where $A$ and $B$ denote the sets of significant voxels for the two features. The Dice coefficient measures spatial co-localization while accounting for differences in signal extent. Because all maps are reconstructed from the same fPCA bases and inherit the same spatial covariance, the expected value of both metrics for unrelated features is not zero, and they are reported as descriptive measures of co-location. Together, correlation- and Dice-based metrics characterize how molecular association patterns converge or diverge across omics layers.

#### 1.15 Biological enrichment and network integration

Finally, integrative enrichment and network analyses join the gene, protein, metabolite, and disease layers, placing the location-level associations of §§1.5–1.14 in a common biological framework. For GWAS-derived signals, the representative voxel (minimum P-value with the tract-level lead SNP) summarizes each locus, and FUMA (v1.5.7; 106) annotates the genome-wide summary statistics at these voxels.

Cis-eQTL relationships from GTEx v8 (GRCh37/hg19), retained at an FDR threshold of 0.05 and drawn primarily from brain tissues, map genome-wide significant SNPs to one or more candidate genes. Mapping proteomic features from the UKB Pharma Proteomics Project to their encoding genes unifies GWAS and proteomic results at the gene level.

g:Profiler (v0.2.2; 107) tests gene-set enrichment across Gene Ontology (Biological Process, Molecular Function, Cellular Component), KEGG, and Reactome, separately for gene sets derived from GWAS and proteomic associations, which we then integrate for interpretation (Supplementary Table 11); the g:SCS method controls multiple testing, and adjusted $P < 0.05$ defines significance.

STRING (v12.0; 108) builds protein–protein interaction (PPI) networks from experimentally supported interactions with confidence score $> 0.4$; its network enrichment statistic evaluates whether the observed connectivity exceeds random expectation, and Gene Ontology terms and UniProt keywords annotate clusters enriched for extracellular signaling, synaptic processes, and metabolic pathways (Supplementary Table 11).

The Network Analysis module in MetaboAnalyst (v5.0; 109) joins the molecular layers into gene–metabolite–disease networks: the Human Metabolome Database (HMDB) supplies gene–metabolite relationships, MetaboAnalyst's internal ontology framework assigns disease annotations, and no pathway pre-filtering restricts the included entities. Analysis of the resulting networks identifies connected components and highly interconnected subnetworks representing shared biological mechanisms across omics layers, including neuronal maintenance, oxidative stress, and lipid metabolism (Supplementary Table 11).

#### 1.16 Druggability annotation and spatial closure

Genes at the 898 FA, MD and AD loci were taken from the FUMA mapping of §1.15 (657 unique genes) and annotated with Open Targets Platform tractability evidence (small-molecule, antibody and PROTAC buckets; release 26.06, queried through the GraphQL API on 2026-09-22) [110], with compounds and their highest clinical phase, mechanism and indication from the ChEMBL-derived drug records in the same release [111], and with drug–gene interactions from DGIdb (export of 2026-09-22) [112]. Genes were placed in four tiers following Open Targets: approved-drug target, clinical-stage target, tractable without a reported compound, and not currently tractable (Supplementary Table 12); DGIdb interactions are reported separately, because DGIdb lists compounds for some genes that Open Targets does not tier as tractable (for example PDE8B and TNIK). For the 35 genes in the first two tiers, the tracts in which the locus footprint is enriched were taken from the tract-level locus assignments. Gene assignment is positional and no adjustment was made for locus size or gene density; the annotation is a prioritization.

Spatial closure asks, for each colocalized locus, whether its footprint coincides with the region in which the disease's genetic correlation with white matter concentrates. For each of the 47 colocalized FA loci (§1.12) and each diffusion measure, the footprint was the set of voxels at which the lead variant passes the atlas threshold ($P < 9.12 \times 10^{-12}$) and each disease map the set of voxels at which the genetic correlation with that trait passes $P < 9.12 \times 10^{-6}$ ($0.05/M_{\text{eff}}$); voxels with a reconstructed $P$ of exactly zero (1,833 voxels on 19 fibers, identical across variants in the MD and AD maps and attributable to the aggregation step) were set to missing on both sides. The fold enrichment is the observed overlap divided by $n_{\text{footprint}} \times n_{\text{map}}/n_{\text{valid}}$, and is bounded above by $n_{\text{valid}}/n_{\text{footprint}}$. Its null distribution was obtained from 10,000 draws in which every fiber's footprint pattern was circularly shifted along the fiber's 100 points by an independent random offset and fibers were then permuted within their JHU tract (clusters without a tract assignment were permuted among themselves), which preserves the number of footprint voxels, their along-fiber runs and their distribution across tracts; $P = (1 + \#\{\text{null} \geq \text{observed}\})/(1 + 10{,}000)$. A pair was testable when both sets were non-empty in that measure (828 pairs). Benjamini–Hochberg correction was applied across all testable pairs and, separately, across the 56 pairs in which the disease is the trait the locus colocalizes with ("own-trait" tests; Alzheimer's disease, schizophrenia, bipolar disorder, educational attainment, cognitive performance, intelligence, ADHD, insomnia, major depressive disorder and autism have maps). Footprints defined by Benjamini–Hochberg FDR over voxels, and threshold-free versions using the top 500, 2,000 and 10,000 voxels by $-\log_{10} P$ against the same null, are reported as sensitivity columns in Supplementary Table 13. Results are given in §2.8, Supplementary Table 13 and Supplementary Fig. 47.

#### 1.17 Cross-modality comparison framework

The preceding sections describe fractional anisotropy (FA), the primary phenotype. The full pipeline (preprocessing, dimensionality reduction, GWAS, and voxel-level reconstruction) runs identically on mean diffusivity (MD) and axial diffusivity (AD), making all three diffusion modalities directly comparable.

Three complementary questions structure the comparison. First, do discovery loci preferentially localize to similar tract systems across modalities? Mapping each modality's tract-level lead SNPs to their associated white-matter regions and comparing patterns of tract-level enrichment identifies loci shared across tract systems and summarizes the distribution of shared and modality-specific tract associations. Second, are spatial association patterns conserved across modalities? For each locus detected in multiple modalities, the reconstructed voxel-wise Z-score maps yield Pearson correlations and overlap metrics (e.g., Dice coefficients) within the locus-specific regions. Third, do shared locus signals arise from the same underlying causal variant? The same Bayesian colocalization framework compares summary statistics within each locus and estimates posterior probabilities of shared versus distinct causal variants. Together, these analyses characterize the extent to which genetic influences on white-matter microstructure are conserved or modality-specific across diffusion measures.

##### 1.18 Sensitivity analyses

Sensitivity analyses probe the robustness of key findings to analytic choices in the reconstruction, replication, and downstream prioritization steps. Because voxel-level replication depends on the threshold used to reconstruct association maps in the validation cohort, we repeated the replication analyses at alternative significance thresholds of $P < 1 \times 10^{-3}$ and $P < 1 \times 10^{-2}$ in addition to the primary threshold of $P < 1 \times 10^{-4}$, re-computing genetic replication counts, spatial overlap summaries, and the proportion of loci meeting the predefined replication criteria.

Alternative definitions of spatial overlap between discovery and replication signals, including any nonzero overlap and more stringent thresholds beyond the primary criterion, test the robustness of the spatial replication framework, and varying the posterior probability cutoff that defines strong evidence of a shared causal variant tests the stability of the prioritized locus–trait pairs. Our primary conclusions rest on patterns that remain qualitatively consistent under reasonable variations in thresholding and prioritization criteria.

#### 2 Supplementary Results

These results follow the order of the Methods: properties and statistical calibration of the low-dimensional representations (§§2.1–2.2), heritability and spatial GWAS atlases across the three diffusion modalities (§§2.3–2.4), replication of the identified loci (§2.5), shared polygenic architecture with external traits via genetic correlation (§2.6), and the measured molecular layers (§2.7). Supplementary results for the Mendelian randomization (§1.11) are given in Supplementary Table 9, for the biological enrichment and network analyses (§1.15) in Supplementary Table 11 and Supplementary Figs. 44–46, and for the druggability annotation and spatial closure (§1.16), the latter reported as a negative result, in Supplementary Tables 12–13 and Supplementary Fig. 47 (§2.8).

##### 2.1 Properties of low-dimensional representations

Across the 430 fiber clusters, the adaptive selection rule targeted 80–90% of total variance (achieved range 70–92%), and 8,868 LDRs in total were carried into the FA GWAS (Supplementary Table 1). The LDRs preserve voxel-level information: correlations between the original voxel-level signals and their fPCA-based reconstructions range from 0.85 to 0.99, with a mean of 0.94. The number of retained components varies across clusters, tracking heterogeneity in the spatial complexity and covariance structure of white-matter regions.

The eigenvalue spectra of the fPCA decompositions decay rapidly across clusters, so a relatively small number of components captures most of the variability in voxel-level signals. These eigenvalues also yield each cluster's effective number of independent voxels; summing across clusters gives approximately 5,479 effective independent voxels across the white-matter skeleton, a substantial dimensionality reduction from the original 609,000 voxel measurements.

#### 2.2 Quality control and calibration

Quantile–quantile (QQ) plots for each LDR compare the observed distribution of test statistics against expectations under the null hypothesis. The genomic inflation factor ($\lambda_{\mathrm{GC}}$) centered on 1 (mean = 1.046, range = 0.983–1.185): the association statistics show no evidence of systematic inflation beyond that expected from polygenic signal.

The two-step ridge regression procedure with leave-one-chromosome-out (LOCO) adjustment removes polygenic background effects prior to association testing, which improves calibration and controls type I error across the high-dimensional imaging phenotypes.

Voxel-level association statistics reconstructed from the LDR-level results likewise match null expectations, with no evidence of artifactual inflation introduced by the reconstruction procedure. Both the LDR-level and voxel-level analyses are therefore well calibrated and support downstream genetic and spatial inference.

#### 2.3 Voxel-wise heritability patterns

Extending the voxel-level heritability analyses to mean diffusivity (MD) and axial diffusivity (AD) characterizes the shared and modality-specific genetic architecture of white-matter microstructure. Using the same UK Biobank discovery cohort (Phases 1–6; $n$ = 56,313), we performed GWAS on the LDRs for each modality and reconstructed voxel-level SNP heritability estimates.

Genetic influence pervades all three modalities: 94.2% of voxels pass correction for the effective number of voxel-level tests for MD and 93.4% for AD ($P < 0.05/M_{\mathrm{eff}}$, with modality-specific $M_{\mathrm{eff}}$ = 2,946 and 3,895), comparable to the 90.7% observed for FA (proportions of significant voxels use each measure's own $M_{\mathrm{eff}}$ over its non-missing voxels), with mean voxel-level heritability of 15.9% for MD and 14.6% for AD, relative to 16.4% for FA (Supplementary Fig. 4). Genetic influences on white-matter microstructure are thus pervasive and detectable across complementary diffusion measures.

Anatomically, the MD and AD heritability patterns recapitulate many spatial features of FA, with strong signals concentrated in major projection and callosal tracts (Supplementary Table 3). Tract means quoted here and in the main text are means over the significant voxels of each tract (the convention of main Fig. 2a); Supplementary Table 3 reports means over all voxels of each tract (25.1% for FA in the GCC) and Supplementary Fig. 6 medians. The genu of the corpus callosum (GCC) shows among the highest mean heritability in all three modalities (FA: 26.1%, MD: 26.2%,

AD: 22.8%), reflecting strong genetic influences on commissural fiber microstructure. By contrast, several other robustly heritable tracts vary markedly within tract: the superior corona radiata (SCR; FA: 21.8%, MD: 17.8%, AD: 18.2%) and external capsule (EC; FA: 20.3%, MD: 16.0%, AD: 15.0%) show the largest within-tract variability across modalities (voxel-level coefficient of variation $> 0.39$), consistent with their complex crossing-fiber and mixed-fiber architecture. The posterior limb of the internal capsule (PLIC; FA: 25.6%, MD: 12.5%, AD: 14.3%) diverges most across modalities, with FA-based heritability among the highest observed but substantially reduced MD and AD estimates; genetic regulation of directional coherence in this densely packed projection pathway is evidently not fully captured by isotropic diffusion measures.

This anatomical pattern extends across the full set of 17 JHU tracts: tract-level heritability is highest across the corpus callosum (genu, body, and splenium), the fornix, and the anterior limb of the internal capsule in all three modalities, and lowest in association tracts including the anterior corona radiata, sagittal stratum, inferior fronto-occipital fasciculus, and posterior thalamic radiation (Supplementary Table 3, Supplementary Fig. 6). Among all 17 tracts, PLIC and the cerebral peduncle (CP) show the two largest FA-versus-diffusivity heritability gaps, consistent with the FA-specific elevation described above for PLIC. Beyond overall magnitude, however, the spatial pattern of heritability within a tract is not always concordant across modalities: correlating voxel-level heritability estimates within each tract between modality pairs, median within-tract correlation is lowest between FA and MD ($r = 0.53$) and progressively higher between FA and AD ($r = 0.64$) and between MD and AD ($r = 0.73$), indicating that the two diffusivity-based measures share a more similar within-tract spatial pattern of heritability than either shares with FA (Supplementary Fig. 5). This spatial concordance is not simply predicted by magnitude concordance: GCC has among the highest heritability in all three modalities, yet its within-tract spatial pattern correlates poorly between FA and the two diffusivity measures ($r = 0.16$ for FA vs. MD; $r = 0.10$ for FA vs. AD), while remaining highly correlated between MD and AD ($r = 0.73$). PLIC shows a similar dissociation, with near-zero FA–MD spatial correlation ($r = -0.09$) despite both modalities showing substantial mean heritability in this tract. These results indicate that even where two modalities agree on a tract's overall heritability, they can implicate largely distinct voxels within it, consistent with FA capturing aspects of directional coherence not fully recovered by isotropic diffusivity measures.

The heritability estimates reproduce across independent cohorts: comparing discovery (Phases 1–6) and validation (Phase 7; $n$ = 15,872), voxel-wise correlations of heritability maps reach r = 0.75 for MD and r = 0.68 for AD, comparable to the FA correlation of 0.72, and cluster-level correlations exceed 0.9 in all modalities (Supplementary Fig. 4).

Together, these results show that the genetic architecture of white-matter microstructure is largely shared across diffusion modalities, with consistent anatomical organization and high reproducibility, while modality-specific patterns likely reflect the distinct microstructural sensitivities of FA, MD, and AD.

#### 2.4 Spatial GWAS atlases

The MD and AD analyses identified 238 and 345 genome-wide significant loci, respectively (Supplementary Table 6, Supplementary Table 7), providing complementary spatial resolution to the 315 FA loci reported in the main text (Supplementary Table 5). Projecting the low-dimensional GWAS summary statistics back into image space builds a voxel-level GWAS atlas for each modality and visualizes each locus's footprint across the white-matter skeleton.

Across both MD and AD, genome-wide significant loci span a similarly broad range as FA, from single-tract, highly localized effects to distributed patterns spanning many white-matter systems: 33.0% of FA loci, 38.7% of MD loci, and 39.1% of AD loci involve only a single JHU tract, while 22.2%, 24.4%, and 22.0%, respectively, span five or more tracts (maximum 17, 16, and 17 tracts at a single locus for FA, MD, and AD). All three atlases enrich the same major projection and callosal pathways: the superior longitudinal fasciculus (SLF) is the most frequently implicated tract in FA (42.9% of loci) and MD (39.9%), and the second most frequent in AD (38.8%, behind the superior corona radiata at 40.3%); the corona radiata (anterior, superior, and posterior subdivisions combined) is present at 55.0–59.7% of loci across all three modalities, and the corpus callosum (genu, body, and splenium combined) at 43.5% of FA loci and 52.5–55.9% of MD/AD loci. The internal capsule, including its posterior limb (PLIC), is present at 11.0–14.3% of loci overall (PLIC alone in 5.2–7.1%).

Association tracts (SLF, sagittal stratum, external capsule, cingulum, inferior fronto-occipital fasciculus) are present at 62.9% of FA loci, 55.5% of MD loci, and 52.2% of AD loci; callosal tracts are present at 43.5% of FA loci, 55.9% of MD loci, and 52.5% of AD loci. Mean tracts per locus are similar across modalities (3.42 for FA, 3.41 for MD, 3.19 for AD). MD loci cover a larger average discovery voxel footprint than FA or AD (mean 4,824 discovery voxels/locus, median 155, versus FA's mean 2,261/median 118 and AD's mean 1,984/median 94), reflecting a subset of very large MD loci (maximum 246,709 voxels, versus 185,624 for FA and 136,026 for AD); AD's typical per-locus footprint is the smallest of the three modalities.

To assess spatial concordance directly, we reconstructed voxel-level (609,000-location) statistics for FA and, at the 47 FA-discovered, colocalization-prioritized loci, for MD and AD, and tested each FA lead SNP's reconstructed signal against the voxel-effective threshold $P5 \times 10^{-8}/M_{\mathrm{eff}} \approx 9.13 \times 10^{-12}$ ($M_{\mathrm{eff}} = 5479.48$). MD reached this threshold at 19 of 47 loci (40.4%) and AD at 23 of 47 (48.9%); 16 of 47 (34.0%) were significant in all three modalities simultaneously. Among these 16 tri-modality loci, three-way Dice overlap (restricted to the 592,467 voxels with valid reconstructed statistics in all three modalities) was nonzero for 4 of 16 (dice = 0.02–0.03), and pairwise overlap varied across loci, with no single modality pair consistently dominating.

At the locus level – treating each modality's genome-wide significant loci as independently discovered, rather than testing a single FA lead SNP's reconstructed signal in the other modalities – cross-modality concordance among the 47 colocalization-prioritized FA loci was higher. Matching each FA lead SNP to the nearest independently significant MD or AD lead SNP within ±250 kb identified a corresponding locus in at least one other modality for 34 of 47 loci (72.3%), and in both MD and AD for 21 of 47 loci (44.7%); 2 loci had a matching MD locus only, 11 had a matching AD locus only, and 13 (27.7%) had no independently significant locus in either modality within this window.

Representative examples illustrate the range of these patterns. At rs9262135 (6p21) (Supplementary Fig. 35), FA associates across 11 tracts (3,145 discovery voxels); MD reaches genome-wide significance 110 kb away at rs3130117 across 9 tracts (13,957 voxels), and AD reaches significance 25 kb away at rs9262141 across 12 tracts (18,426 voxels) – a large, pleiotropic locus with independent, substantial signal in all three modalities, consistent with the coincident, multi-tract pattern seen for the largest loci in each individual atlas. At rs6062264 (20q13.12) (Supplementary Fig. 36), FA signal spans 13 tracts (12,589 voxels) and an independently significant MD locus lies 10 kb away (rs12479469; 2 tracts, CP/SCR; 105 voxels), with no AD locus within 250 kb; this pairs with a near-zero FA–MD voxel overlap in the reconstruction analysis above (Dice = 0.0009), indicating that the two modalities' significant voxels occupy largely distinct spatial territory at this locus despite both reaching independent significance. At rs1747223 (13q14.11) (Supplementary Fig. 37), FA spans 3 tracts (GCC/PCR/PTR; 51 voxels) and AD reaches significance at the same lead SNP position with 1 tract (PCR; 117 voxels), with no MD locus nearby, matching the FA+AD-only detection pattern from the voxel-level reconstruction for this locus.

A small number of loci show significant association in MD and/or AD with no corresponding FA signal. At 18p11.21 (rs2306811, chr18:21,705,624) (Supplementary Fig. 38), MD ($P = 1.97 \times 10^{-19}$; tracts BCC, SCR; 2,339 discovery voxels) and AD ($P = 4.46 \times 10^{-21}$; tracts BCC, GCC, SCR; 2,500 discovery voxels) are both independently genome-wide significant, with no FA locus within 250 kb of this position, consistent with the main text's description of 18p11.21 as an MD/AD-coherent locus with no corresponding FA signal. More broadly, the independent MD and AD locus lists contain loci with no nearby counterpart in either of the other two modalities even outside this colocalization-prioritized set; for example, rs78903072 (17q21.31) (Supplementary Fig. 39) is MD-significant across 15 tracts (140,058 discovery voxels, one of the largest single loci in any of the three atlases) with no FA or AD locus within 250 kb. These loci have not yet been evaluated for disease-trait colocalization and are reported here only to illustrate that modality-specific discovery extends beyond the FA-anchored, colocalization-prioritized set profiled elsewhere in this section.

Together, independent locus-level concordance across FA, MD, and AD is more common (72.3% of colocalization-prioritized loci show a match in at least one other modality) than literal spatial (voxel-level) coincidence at those same loci (34.0% detected in all three modalities' reconstructed maps; three-way Dice nonzero in only 4 of 16 such loci). This gap suggests that FA, MD, and AD frequently share genetic architecture at the locus level while marking distinct, only partially overlapping, subsets of the white-matter voxel space – consistent with the three measures indexing complementary rather than redundant aspects of microstructure even where they are driven by shared underlying variants.

#### 2.5 Replication of white-matter loci

We replicated the identified loci in an independent held-out cohort (UK Biobank Phase 7; $n = 15{,}872$; see §1.1) against two complementary criteria: locus-level replication (a variant within ±250 kb of the discovery lead passing the Phase 7 screen; §1.8), and spatial concordance of voxel-level signal patterns.

Locus-level replication is nearly universal. Of the 315 FA loci identified in the discovery GWAS, 311 (98.7%) replicate in at least one tract in the validation cohort; rates are similarly high for MD (233 of 238 loci, 97.9%) and AD (339 of 345 loci, 98.3%).

Spatial footprints replicate less completely, as the smaller replication sample predicts. Among the locus-level replicated loci, 224 of 311 FA loci (72.0%), 158 of 233 MD loci (67.8%), and 218 of 339 AD loci (64.3%) show nonzero spatial overlap between discovery and replication association maps; defining substantial spatial replication as at least 10% overlap, 207 of 311 FA loci (66.6%), 140 of 233 MD loci (60.1%), and 202 of 339 AD loci (59.6%) meet the threshold. FA loci overlap modestly more on average (mean 0.43) than MD (mean 0.39) or AD (mean 0.36), consistent with the generally higher heritability of FA relative to diffusivity measures. Across modalities, replicated loci span a mean of approximately 3.2–3.4 tracts, and the majority of multi-tract loci reproduce their discovery footprint across multiple tracts in the replication sample.

The illustrative loci of §2.4 behave as their discovery footprints predict. The 12q23.3 locus (rs12146713) replicates with high voxel overlap across 12 FA tracts (overlap 0.72), 10 MD tracts (0.71), and 13 AD tracts (0.63), confirming its role as a broadly distributed cross-modality locus. The 5q14.1 locus (rs115877304) replicates across all five of its discovery tracts in FA and AD (overlap 0.21 and 0.02, respectively) and its single MD tract (overlap 0.20), consistent with its modality-shared signal. At 7p22.3, the FA footprint replicates across eight tracts (overlap 0.64), while the distinct MD and AD lead variants replicate across nine and eleven tracts, respectively (overlap 0.39 and 0.41), supporting modality-specific fine-mapping at a shared genomic region. The 18p11.21 locus (rs2306811), absent in FA, replicates in both MD and AD across the body of the corpus callosum and adjacent tracts with modest spatial overlap (MD 0.05, AD 0.003), an attenuation of replication voxel distributions that the reduced sample size explains.

Against prior work, the atlas substantially expands the known architecture. Comparing with [113], the largest published GWAS of white-matter tract microstructure to date, the majority of loci identified here are new: 258 of 315 FA loci (81.9%), 186 of 238 MD loci (78.2%), and 298 of 345 AD loci (86.4%) show no overlap with Zhao et al. within a ±500 kb window on the same chromosome, with even higher novelty rates when requiring overlap within the same tract (87.0%, 85.3%, and 92.5%, respectively). At the same time, our discovery sample recovers the substantial majority of the distinct loci previously identified by Zhao et al.: 57 of their 70 global FA loci (81.4%), 52 of 61 MD loci (85.2%), and 46 of 55 AD loci (83.6%). This high recovery rate, combined with the large number of novel loci, reflects the improved sensitivity of the fPCA-based framework: representing each tract's diffusion signal through multiple low-dimensional components rather than a single summary statistic enables reconstruction of voxel-level GWAS signals and detection of spatially focal effects that tract averaging would attenuate. A modestly larger discovery cohort (n = 56,313 versus n = 34,024 in the Zhao et al. British-ancestry discovery GWAS) adds power.

Collectively, the high locus-level replication rates (>97% across all three modalities) and the spatial concordance at the majority of loci indicate that the identified genetic associations are reproducible within the UK Biobank; because the Phase 7 cohort shares scanners, protocol, ancestry and processing with the discovery cohort, this replication guards against sampling error rather than against cohort-specific confounding. The lower spatial replication rates relative to locus-level rates are consistent with the attenuation of spatial footprints expected in the smaller replication sample (Supplementary Fig. 25).

#### 2.6 Genetic correlation results

We quantified shared polygenic architecture between white-matter microstructure and a panel of 14 neuropsychiatric, neurodegenerative, and cognitive traits (Supplementary Table 2) using the two-sample genetic-correlation framework implemented in HEIG (see §1.10). Voxel-level correlations were aggregated to 17 JHU tracts (Supplementary Fig. 7) and to 66 pairwise combinations of resting-state functional networks (Supplementary Fig. 8; Supplementary Table 4), in addition to the per-trait voxel-level atlases presented individually in Supplementary Figs. 9–22.

At the network-pair level (Supplementary Fig. 8), FA shows FDR-significant genetic correlation (Benjamini–Hochberg, $\alpha = 0.05$) with at least one of the 14 traits in 395 of 924 network-pair×trait combinations (42.7%), of which 75.9% are directionally consistent across their constituent fiber clusters (Supplementary Table 4). Trait involvement ranges widely in spatial breadth: schizophrenia is significant in 60 of 66 network pairs, the broadest of any trait, with the strongest signal concentrated in the frontoparietal control network and its cross-network connections (Frontoparietal–Frontoparietal, $z = -4.99$; Default–Default, $z = -5.45$; Default–Frontoparietal, $z = -4.52$; Supplementary Fig. 15). Educational attainment is significant in 32 of 66 pairs, concentrated instead in subcortical and frontoparietal connections (Sub–Sub, $z = 8.21$; Frontoparietal–Frontoparietal, $z = 7.97$; Language–Sub, $z = 7.50$; Supplementary Fig. 19). Alzheimer's disease (49 pairs; Supplementary Fig. 16) and depression (41 pairs; Supplementary Fig. 12) show similarly broad network-pair involvement, whereas ADHD (3 pairs; Supplementary Fig. 9) and autism spectrum disorder (1 pair; Supplementary Fig. 10) are the most spatially restricted of the 14 traits.

At the voxel level (Supplementary Figs. 9–22), significant genetic correlations remain generally sparse, with most traits showing associations in only a small fraction of locations after multiple-testing correction. The surviving signals organize spatially rather than scattering at random, aligning with the same network- and tract-level structure summarized here: traits with broad network-pair involvement (schizophrenia, Alzheimer's disease, depression) show voxel-level signal distributed across commissural, projection, and association pathways, whereas the more spatially restricted traits (educational attainment, ADHD, autism spectrum disorder) show voxel-level signal concentrated within the specific tracts and network pairs identified above.

Aggregate correlation estimates vary in sign as well as magnitude across traits. The signed aggregate of §1.10 is a precision-weighted combination that summarizes direction and consistency rather than the magnitude of $r_g$, and its working $z$-statistics are large because fiber clusters are treated as independent; magnitudes are therefore quoted below as the mean of the voxel-level $r_g$ estimates in each unit (Supplementary Table 4). Educational attainment shows uniformly positive tract- and network-pair-level correlations with FA (signed aggregate +0.016 in the posterior limb of the internal capsule, PLIC), while schizophrenia and bipolar disorder show uniformly negative correlations (signed aggregate −0.0015 at the superior longitudinal fasciculus for both traits), consistent with the patterns reported in the main text for Alzheimer's disease and depression.

At the tract level (Supplementary Table 4, Supplementary Fig. 7), aggregation of voxel-wise signals identifies 145 of 238 possible tract-trait pairs as FDR-significant in FA (60.9%), of which 77.2% (112) are directionally consistent within tracts (signed test), indicating coherent genetic effects along anatomically defined pathways, and 70 of the 145 are also significant under the dependence-robust Cauchy test (173 of the 395 significant network-pair cells). Beyond the major associations described in the main text, schizophrenia is significant in all 17 of 17 JHU tracts, the broadest tract-level involvement of any trait, most strongly at the superior longitudinal fasciculus (SLF; working $z = -8.07$), anterior corona radiata (ACR; $z = -6.29$), and sagittal stratum (SS; $z = -5.82$); bipolar disorder is significant in 13 of 17 tracts, likewise strongest at the SLF ($z = -6.44$), confirming that schizophrenia and bipolar disorder associate most strongly with the superior longitudinal fasciculus, with additional significant correlations in the genu, body, and splenium of the corpus callosum. Educational attainment is significant in 13 of 17 tracts; its largest-magnitude single-tract correlation is in the posterior limb of the internal capsule (PLIC; signed aggregate 0.016, $q = 0.004$), while its strongest statistical evidence lies in association and projection tracts (superior corona radiata, $z = 11.01$; ACR, $z = 10.99$; SLF, $z = 9.47$; body of the corpus callosum, $z = 9.37$). At the opposite extreme, ADHD (4 of 17 tracts) and autism spectrum disorder (0 of 17 tracts) show minimal tract-level FA signal.

The same overall pattern recurs, at a somewhat lower yield, in mean diffusivity (MD: 133 of 238 tract pairs, 55.9%; 392 of 924 network pairs, 42.4%) and axial diffusivity (AD: 112 of 238 tract pairs, 47.1%; 271 of 924 network pairs, 29.3%; Supplementary Table 4, Supplementary Figs. 7, 8).

#### 2.7 Multi-omics associations and cross-omics spatial concordance

Extending the multi-omics analyses of the main text to the full discovery-validation screen (Supplementary Figs. 41–43) confirms the modality ordering. Pooling discovery and validation voxel counts across FA, MD, and AD, 131 of 195 metabolomic features (67.2%) clear the 10% discovery-validation overlap gate, compared with 10 of 68 proteomic features (14.7%) and 2 of 21 biomarkers (9.5%); metabolomic features also account for 6,170,747 of 6,290,629 pooled discovery-significant voxels across the three omics layers (98.1%) (Supplementary Table 10). Considered separately by modality, this metabolomics-dominant pattern holds throughout, though the metabolomic replication rate itself declines from FA to AD: 108 of 137 metabolomic features (78.8%) clear the overlap gate in FA, versus 124 of 179 (69.3%) in MD and 112 of 186 (60.2%) in AD, while proteomic (FA 1/7; MD 9/52; AD 5/32) and biomarker (FA 2/8; MD 2/19; AD 2/16) replication remain comparably low across all three modalities; metabolomic features likewise dominate discovery-significant voxels within each modality individually (FA 96.7%, MD 98.1%, AD 98.6% of that modality's voxels). Clinical biomarkers thus show the least extensive and least reproducible spatially structured association with white-matter microstructure of the three molecular layers.

Beyond the features highlighted in the main text, individual discovery-phase locus atlases (Supplementary Figs. 48–62) illustrate the range of additional significant associations: neuronal and synaptic signaling proteins (ADGRG1/GPR56, BAIAP2/IRSp53, NPY, SYT1) and the molybdopterin-synthesis enzyme MOCS2 among proteomics; lipid-related metabolites spanning fatty-acid composition ratios (e.g., the saturated, omega-6, and polyunsaturated fatty acid ratios) and HDL lipoprotein subfraction ratios among metabolomics; and hematological and endocrine markers (albumin-adjusted calcium, iron, free thyroxine, lymphocyte count, red blood cell count) among biomarkers (Supplementary Table 10). Footprints range from localized signals in individual pathways to distributed patterns spanning multiple association and projection tracts (Supplementary Figs. 41–43). Molecular influences on white-matter microstructure thus extend beyond the most prominent signals emphasized in the main text and involve a broad set of biological pathways.

Pairwise Pearson correlations of the voxel-level Z-score maps across the 271 significantly associated proteomic, metabolomic, and biomarker features (161, 100, and 10 features, respectively) quantify cross-omics spatial structure (Supplementary Fig. 40B), alongside the corresponding phenotypic correlations measured in the same individuals (Supplementary Fig. 40A). Consistent with the main text, spatial association maps cluster primarily by omics type: metabolomic features cohere most strongly within type (mean $|r| = 0.41$), proteomic features next ($|r| = 0.19$), and biomarkers least ($|r| = 0.09$, close to the cross-type average of $|r| = 0.14$).

Spatial concordance and phenotypic correlation dissociate in both directions across the 36,414 feature pairs with an evaluable phenotypic correlation, which correlate positively overall ($r = 0.78$) but with a non-trivial minority of exceptions. 122 pairs (0.3%) combine low phenotypic correlation ($|r| < 0.1$) with high spatial concordance ($|r| > 0.5$); for example, the omega-6 and polyunsaturated fatty-acid ratios each correlate with glutamine at $|r| < 0.06$ phenotypically but $|r| > 0.7$ in their voxel-wise effect maps. Conversely, 231 pairs (0.6%) combine high phenotypic correlation ($|r| > 0.5$) with weak spatial concordance ($|r| < 0.1$): several members of the Nightingale lipoprotein subfraction panel (e.g., concentration of small LDL particles versus total cholesterol minus HDL-C, cholesteryl esters in LDL, or cholesterol in small and medium VLDL) correlate at $r > 0.9$ phenotypically yet show near-uncorrelated ($r < 0.1$) spatial association patterns with white matter. Marginal feature correlations alone thus do not capture shared spatial genetic architecture.

Dice coefficients computed on the same feature panel after FDR correction (§1.14) identify substantial spatial overlap (Dice $\geq$ 0.1) for 12,663 of all 36,585 feature pairs (34.6%), of which 3,147 (24.8%) cross omics-type boundaries – most commonly metabolomic-proteomic pairs (2,972), with fewer biomarker-metabolomic (128) and biomarker-proteomic (47) pairs. The strongest cross-omics overlaps include triglycerides in HDL with the inflammatory chemokines CCL7 (Dice = 0.55) and CCL3 (Dice = 0.46), and glucose with the lipid-metabolism enzyme LCAT (Dice = 0.51) and with ADGRG1 (Dice = 0.49), indicating convergence of metabolic, inflammatory, and neuronal-signaling influences on shared white-matter pathways.

Together, these findings show that multi-omics associations with white-matter microstructure are both modality-dependent – with metabolomic features showing the broadest extent and most reproducible replication – and spatially structured, with convergent effects across distinct molecular layers, including recurrent metabolite-protein overlap, on shared anatomical systems.

#### 2.8 Pharmacological annotation and spatial closure

Of the 657 genes at the 898 FA, MD and AD loci, 24 are targets of approved drugs and 11 of clinical-stage compounds under the Open Targets tiers (Supplementary Table 12); the 35 compound-bearing genes, their locus measures, enriched tracts and colocalized traits are given in main Fig. 5d.

The spatial-closure test (§1.16) asks, for each colocalized locus, whether its footprint coincides with the region in which the disease's genetic correlation with white matter concentrates. Because footprints and disease maps are both organized by tract, a locus with a large footprint overlaps every map by chance, and a null that places voxels at random is too permissive; the rotation null therefore preserves the footprint's size, its along-fiber clustering and its share in each tract while breaking only where along the fiber it sits. The test was applied to all 47 colocalized FA loci in the three diffusion measures. Pairs were testable only when the footprint and the map each contained at least one voxel in that measure, which gave 828 locus–map pairs (423 in FA, 152 in MD and 253 in AD); in FA the Alzheimer's disease map contains no voxel at threshold ($0.05/M_{\text{eff}}$), so Alzheimer's pairings could be tested only in MD and AD, where the map has 105 voxels; the other disease maps are likewise sparse at this threshold (Supplementary Table 13).

The result is largely negative. Sixteen of the 828 pairs, at 13 of the 47 loci, overlapped more than the rotation null predicts (Benjamini–Hochberg $q < 0.05$); the other 34 loci exceeded the null on no map. Twelve of the 16 pairs reached the resolution floor of the null ($P = 10^{-4}$ at 10,000 draws), so their ordering by $P$ is tied; the same 13 loci were obtained with 1,000 draws, with one exchange (DUSP8 out, MOBP in). The APOE/APOC1 locus (19q13.32, rs7256200) served as a positive control: its MD footprint (36,555 voxels, 6% of the skeleton) contained all 105 voxels of the Alzheimer's disease map (16-fold, the maximum attainable for a footprint of this size; $P = 10^{-4}$), but the Alzheimer's disease GWAS behind this map [87] includes the APOE region, so the locus contributes to the map it is tested against and part of the coincidence is expected by construction: the result shows that the test detects a strong known signal, not that it validates the map independently. The remaining closures concentrated in the genu of the corpus callosum and the corona radiata, where the bipolar disorder, schizophrenia and educational attainment maps overlap one another, and we read them as one hotspot rather than as independent results: the 19q13.33 locus at RASIP1 (rs8112983; colocalized with stroke and schizophrenia) held 66 of its 127 FA footprint voxels on the bipolar map in the genu (174-fold; tied at the null floor with 11 other pairs), the 2q35 locus at SMARCAL1 (rs7590302) 41- and 14-fold on the bipolar and schizophrenia maps, the 20q13.33 locus (rs6062264) 12- and 8-fold, and MOBP (3p22.1, rs28810495; colocalized with glomerular filtration rate) 21-fold on the schizophrenia map in the genu, while two loci colocalized with kidney function and cardiometabolic traits (KIAA1614, rs3795503; ICA1L/NBEAL1, rs186399184) overlapped the educational attainment map in the superior corona radiata 16- to 17-fold.

Only one locus coincided with the map of the trait it colocalizes with. Of the 56 locus–trait pairs in which the colocalized trait has a genetic-correlation map ("own-trait" tests), one passed: the 6p21.33 locus in the major histocompatibility complex (rs9262135; colocalized with bipolar disorder, PP.H4 = 0.92; footprint in 11 tracts, 65% spatial replication) held 73 of its 895 FA footprint voxels on the bipolar disorder map against 2.7 expected (27-fold; $P = 10^{-4}$; $q = 0.006$ across own-trait tests and 0.007 across all pairs), with the overlap in the genu of the corpus callosum. This pair carries two caveats. Single-causal-variant colocalization in the extended MHC is unreliable because of the region's long-range linkage disequilibrium (§1.12), so the colocalization that makes this an own-trait test is itself uncertain; and the extended LD means that our positional mapping (C6orf136, with no reported compound) does not resolve the effector gene. The MHC has been linked to tract-averaged FA and MD before through complement component 4 [114]; our lead variant lies ∼1.3 Mb from C4 and its independence from that signal was not tested. We therefore do not count this pair as a positive own-trait closure. The compound-bearing colocalized loci did not close on their own trait: DPP4 (educational attainment), SPI1 (Alzheimer's disease, whose FA map is empty at threshold) and two loci whose genes carry DGIdb-listed but not Open Targets-tiered compounds, PDE8B (type 2 diabetes, which has no genetic-correlation map in our panel) and TNIK (reaction time), overlapped no map, and EPHA3 (colocalized with cognitive performance) overlapped the schizophrenia map in MD (5.8-fold, $q = 0.013$) but not the cognitive-performance map.

Two properties of the test limit what a negative result can show. First, the disease maps are empty or sparse at $0.05/M_{\text{eff}}$ (Alzheimer's disease: 0 voxels in FA, 105 in MD), so the thresholded test has little power for several traits; a threshold-free statistic, in which the footprint voxels are ranked against the continuous $r_g$ map, would suit these traits better and was not computed here. Second, the resolution floor of 10,000 draws ties the strongest pairs. Within these limits, the test detects the APOE–Alzheimer's relationship (with the circularity noted above) and otherwise finds that single-variant footprints, which are focal, rarely coincide with polygenic genetic-correlation maps, which are tract-wide; for each candidate it says which link of the gene–tract–disease chain is missing (the disease map for PDE8B and DPP4, the gene for the MHC locus, the compound for APOE/APOC1) rather than nominating a target. All pairs, with fold, null $P$, both $q$-values, the tracts in which the overlap lies and the sensitivity columns, are listed in Supplementary Table 13 and summarized in Supplementary Fig. 47.

### Supplementary Figures

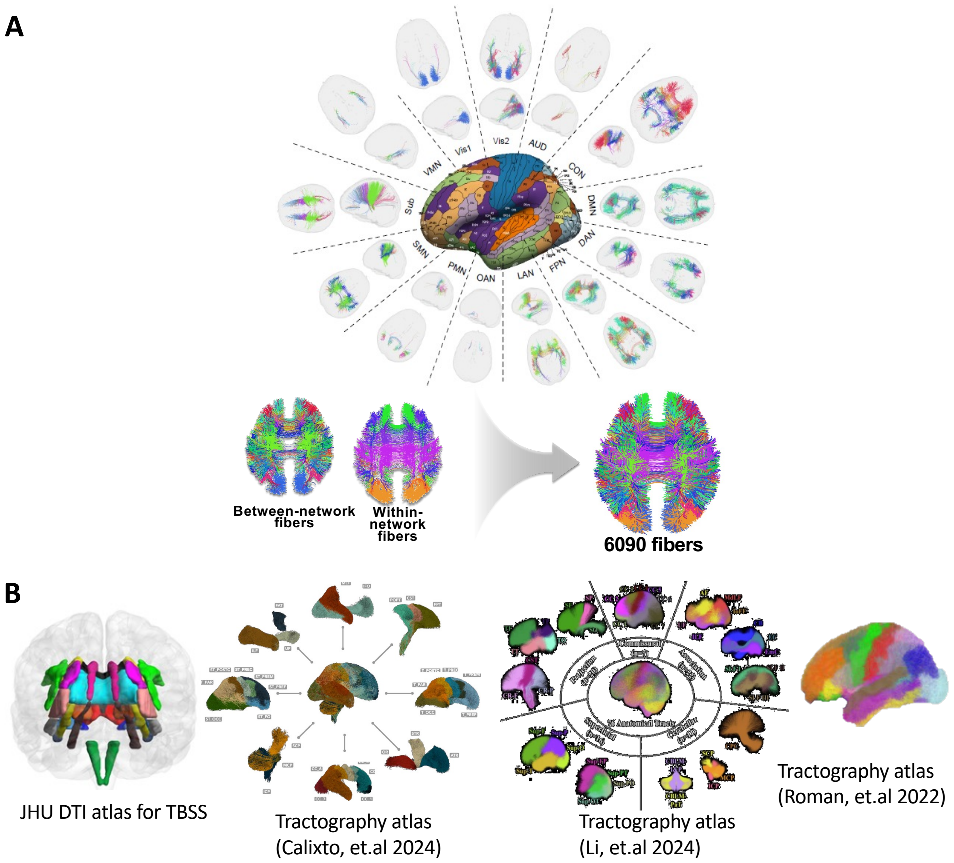

**Supplementary Fig. 1**: **The CBSS atlas encodes the white-matter skeleton as 6,090 anatomically coherent fiber pathways, providing the common anatomical frame for all voxel-level analyses in this study.** **(A)** Atlas construction: connectome-based spatial statistics (CBSS) [64] projects diffusion tensor imaging (DTI) measures onto a white-matter skeleton, and spatially contiguous fiber segments, identified and aggregated across subjects, partition the skeleton into 6,090 anatomically coherent fiber pathways that integrate information from existing tractography atlases. **(B)** Reference tractography and diffusion MRI atlases used during preprocessing and anatomical alignment, supporting consistent mapping of white-matter structures across subjects and studies.

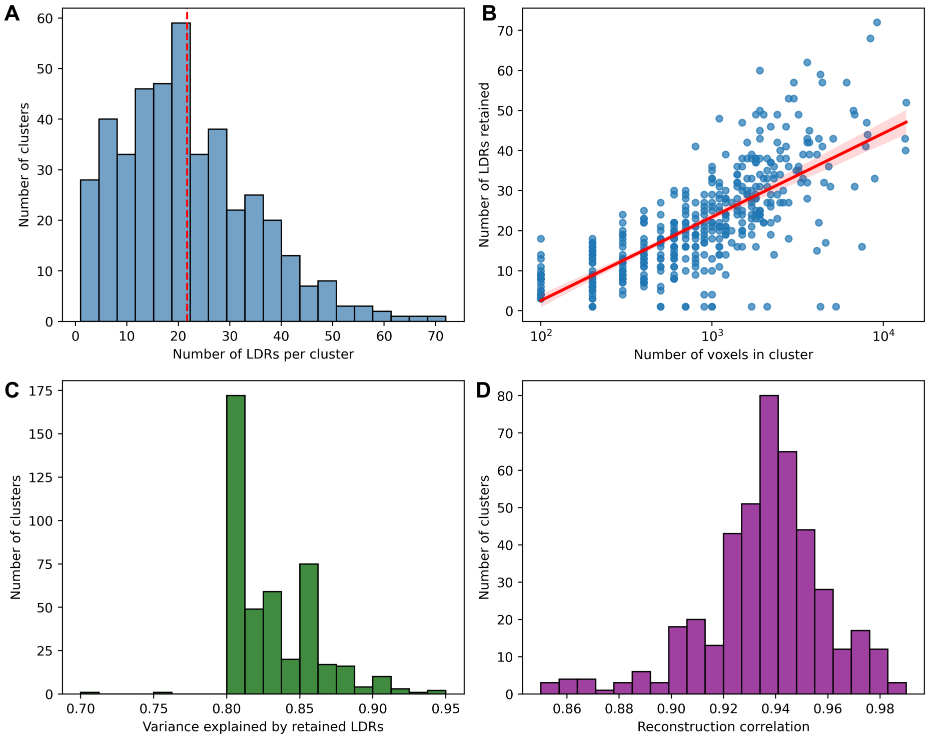

**Supplementary Fig. 2**: **A small number of functional principal components per fiber cluster captures 80–90% of voxel-level diffusion variance with reconstruction fidelity of 0.85–0.99.** **(A)** Number of LDRs retained per fiber cluster; the dashed red line marks the median across the 430 clusters. **(B)** Number of retained LDRs versus cluster size (voxels); each point is a fiber cluster and the red line shows the fitted linear trend; larger clusters require more components to capture their spatial variation. **(C)** Proportion of variance explained by the retained LDRs across clusters; most clusters retain components explaining approximately 80–90% of total variance. **(D)** Correlations between original voxel-level signals and their LDR-based reconstructions across clusters (range 0.85–0.99, mean 0.94).

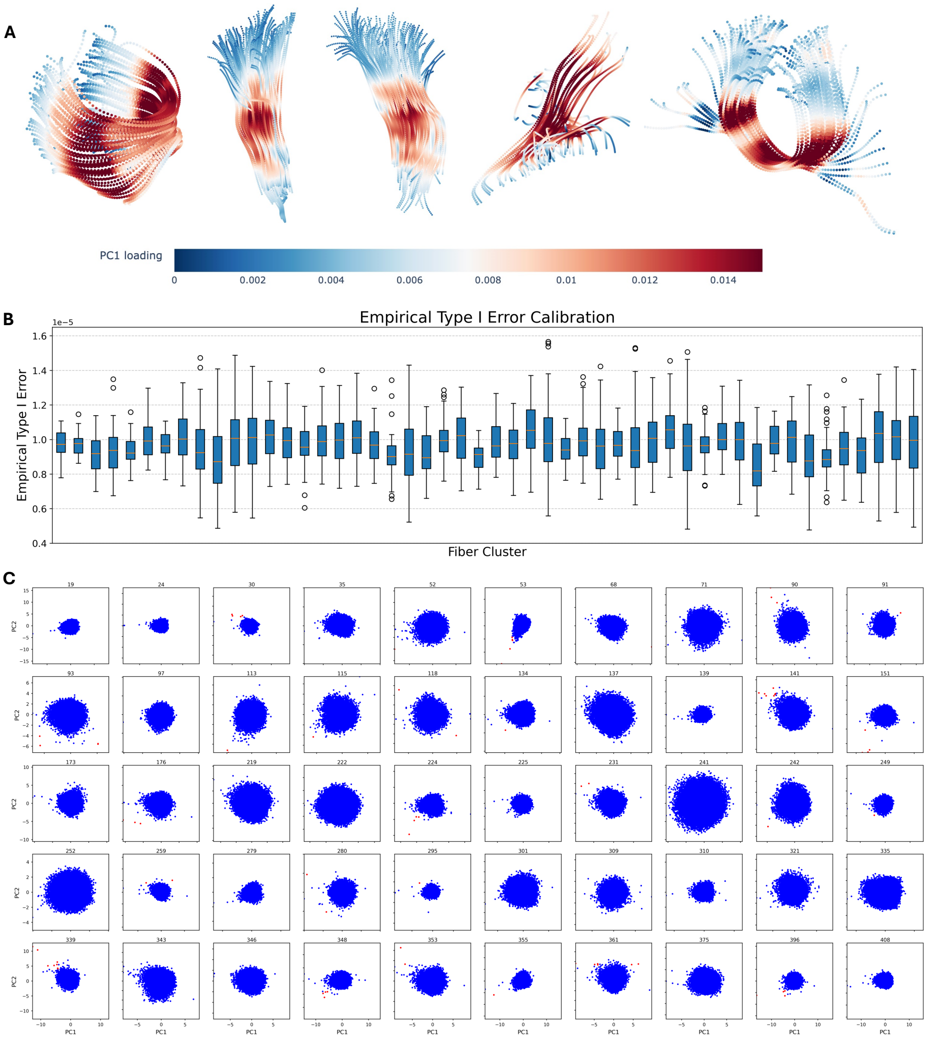

**Supplementary Fig. 3**: **Voxel-level association statistics reconstructed from LDRs are statistically well calibrated, with empirical type-I error at the nominal rate across fiber clusters.** **(A)** Spatial maps of first principal component (PC) loadings for five representative fiber clusters, showing the smooth along-tract patterns the low-dimensional representation captures. **(B)** Empirical type-I error across 50 randomly selected fiber clusters; each boxplot summarizes null-simulation estimates normalized by the expected error rate, centering on 1. **(C)** Scatterplots of PC1 versus PC2 for the same 50 clusters, showing the distribution of LDR scores and potential outliers.

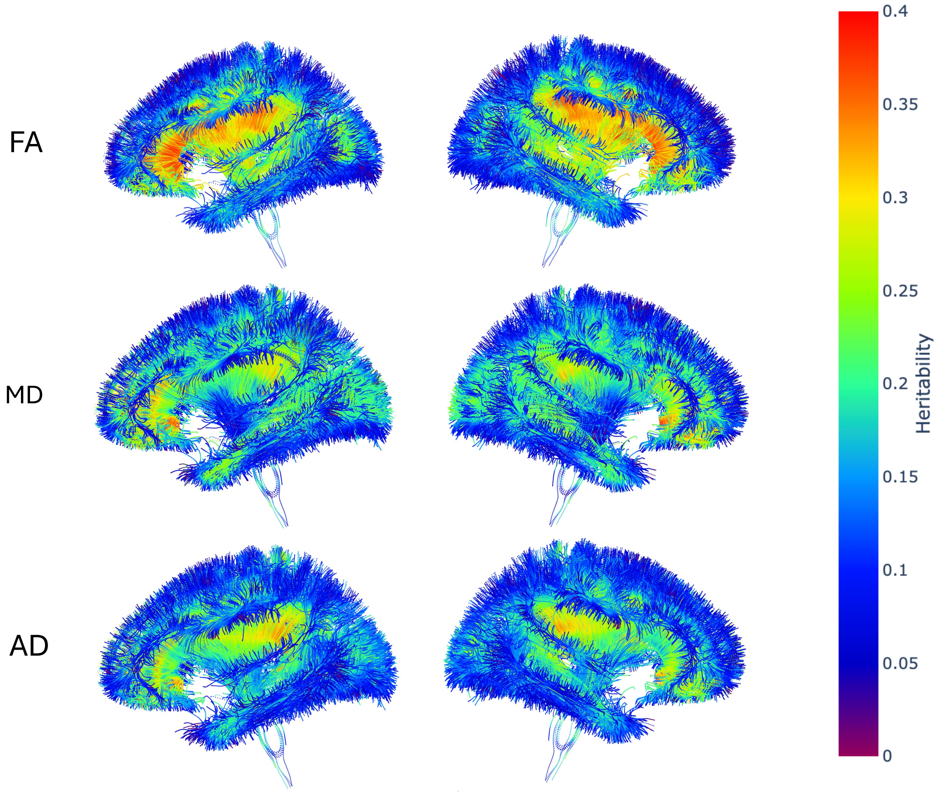

**Supplementary Fig. 4**: **SNP heritability pervades the white-matter skeleton in all three diffusion measures (>90% of voxels significant), with shared anatomical organization and modality-specific divergence.** Voxel-level SNP heritability estimates across the white-matter skeleton for fractional anisotropy (FA; 90.7% of voxels significant, mean $h_v^2 = 16.4\%$), mean diffusivity (MD; 94.2%, 15.9%), and axial diffusivity (AD; 93.4%, 14.6%). Heritability concentrates in projection and callosal tracts in every modality, while the posterior limb of the internal capsule diverges across modalities (mean $h^2$ over significant voxels: FA 25.6% versus MD 12.5% and AD 14.3%).

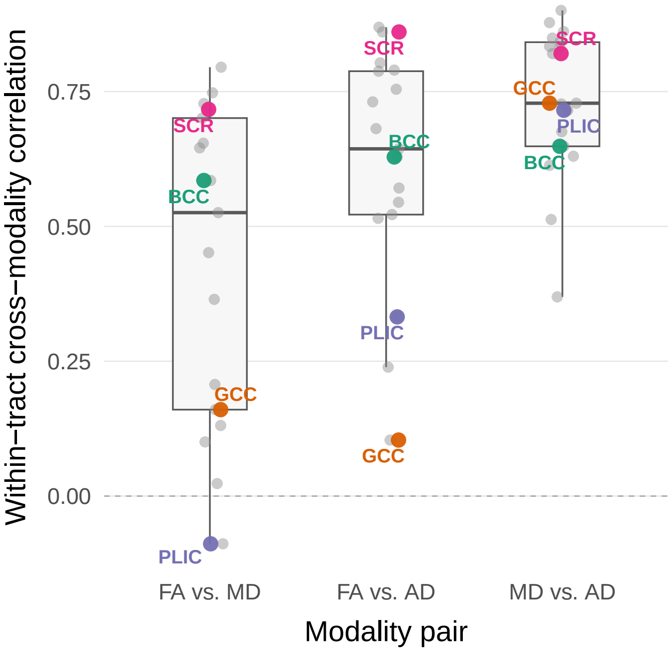

**Supplementary Fig. 5**: **Within-tract SNP heritability correlates most weakly between FA and MD, and diverges most sharply at PLIC and GCC.** Pearson correlation of voxel-level SNP heritability estimates within each of 17 JHU tracts, computed between each pair of diffusion modalities. Correlation is lowest for FA vs. MD (median $r = 0.53$ across the 17 tracts) and progressively higher for FA vs. AD (median $r = 0.64$) and MD vs. AD (median $r = 0.73$), indicating that MD and AD share a more similar within-tract spatial pattern of heritability than either shares with FA. Four tracts are highlighted to illustrate the range of concordance: the superior corona radiata (SCR) and body of the corpus callosum (BCC) correlate strongly and consistently across all three modality pairs (SCR: $r = 0.72$, 0.86, 0.82; BCC: $r = 0.59$, 0.63, 0.65; for FA–MD, FA–AD, and MD–AD respectively); the posterior limb of the internal capsule (PLIC) and genu of the corpus callosum (GCC) diverge sharply depending on which pair is compared, with near-zero or negative FA–MD correlation (PLIC: $r = -0.09$; GCC: $r = 0.16$) and, for GCC, low FA–AD correlation as well ($r = 0.10$), despite both tracts remaining well-correlated between MD and AD (PLIC: $r = 0.72$; GCC: $r = 0.73$); PLIC's FA–AD correlation is intermediate ($r = 0.33$).

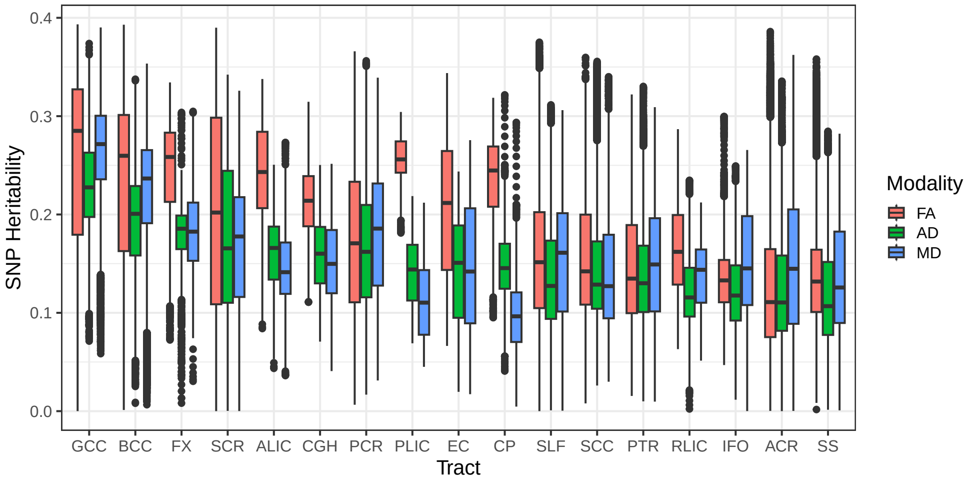

**Supplementary Fig. 6**: **Tract-level SNP heritability is highest in callosal and periventricular projection tracts and lowest in association tracts, with FA exceeding MD and AD by the largest margin in the two tracts showing the sharpest cross-modality divergence.** Median voxel-level SNP heritability within each of 17 JHU tracts, by modality (FA, MD, AD). Heritability is highest in the genu (GCC; median $h^2 = 0.285$ FA, 0.272 MD, 0.228 AD), body (BCC; 0.260/0.237/0.201) of the corpus callosum, the fornix (FX; 0.258/0.183/0.186), and the anterior limb of the internal capsule (ALIC; 0.243/0.141/0.166) across all three modalities. FA heritability exceeds MD and AD by the largest margin in the posterior limb of the internal capsule (PLIC; 0.256 FA vs. 0.110 MD and 0.144 AD) and the cerebral peduncle (CP; 0.245 FA vs. 0.096 MD and 0.145 AD) – the two tracts with the largest FA-vs.-diffusivity heritability gap of all 17 tracts, and the same two tracts identified as the sharpest cross-modality outliers in Supplementary Fig. 5. Association tracts – anterior corona radiata (ACR; 0.111/0.145/0.111), sagittal stratum (SS; 0.132/0.126/0.107), inferior fronto-occipital fasciculus (IFO; 0.133/0.145/0.118), and posterior thalamic radiation (PTR; 0.135/0.149/0.130) – show the lowest heritability across all three modalities; notably, FA does not consistently exceed the other two modalities in this group (e.g., ACR: MD 0.145 > FA 0.111).

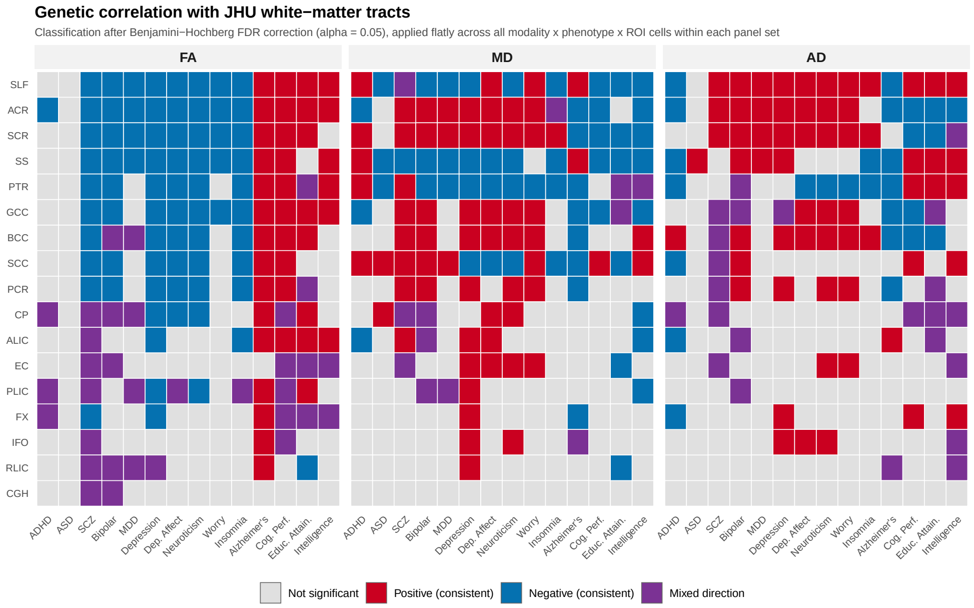

**Supplementary Fig. 7**: **Genetic correlation with the 17 JHU white-matter tracts.** Genetic correlation between tract-aggregated microstructure (location-level estimates aggregated over the fiber clusters assigned to each JHU tract; §1.2, §1.10) and the same 14 phenotypes, across the three diffusion modalities (FA, MD, AD; column panels), for each of 17 white matter tracts (rows). Cells are classified as consistently positive or consistently negative (signed test significant), mixed-direction (Cauchy test significant only) or not significant after Benjamini–Hochberg FDR correction ($\alpha = 0.05$) applied across all modality $\times$ phenotype $\times$ tract cells (§1.10).

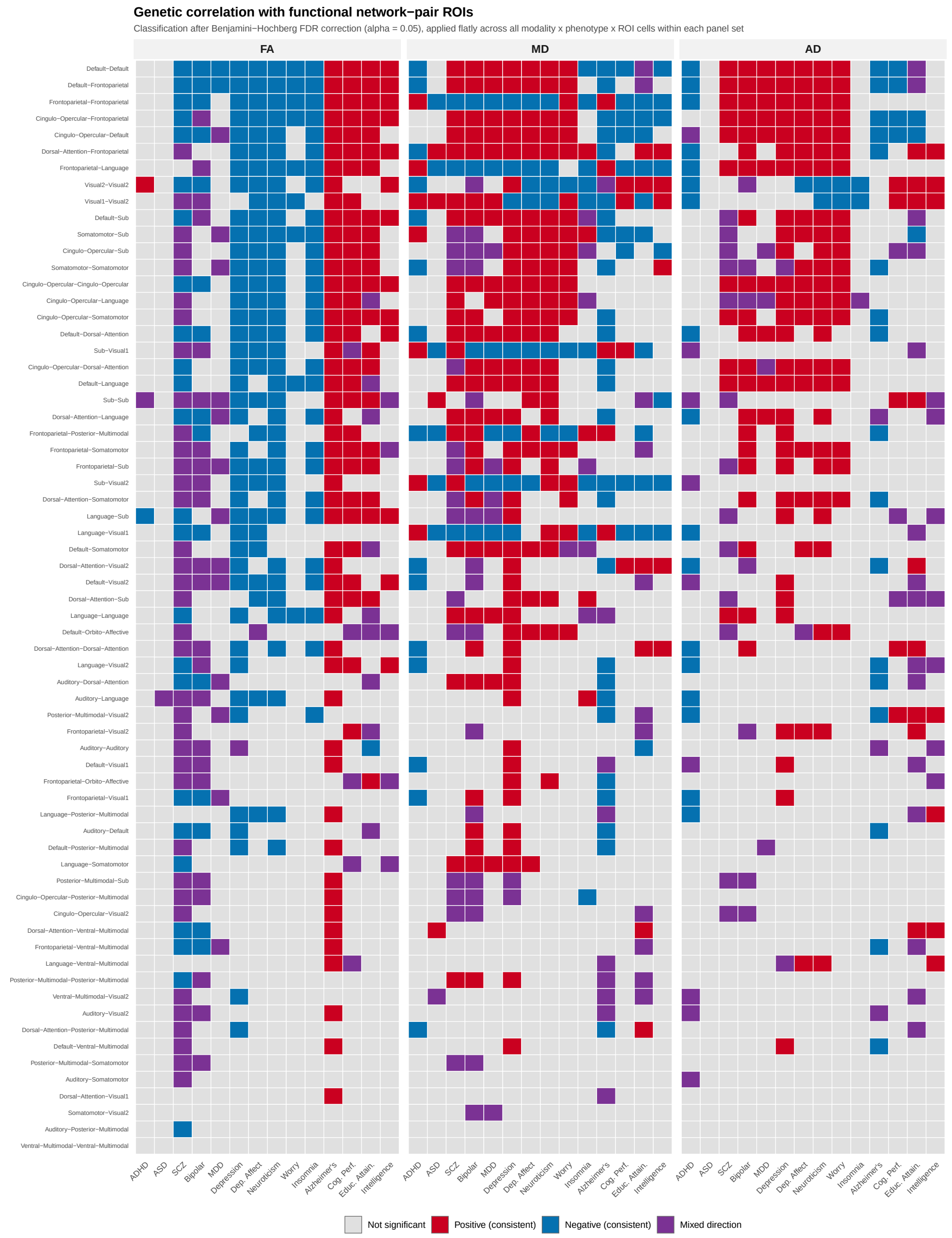

**Supplementary Fig. 8**: **Genetic correlation with functional network-pair ROIs.** Genetic correlation between white-matter microstructure, aggregated over the fiber clusters connecting each pair of functional networks (rows; §1.2), across 14 neuropsychiatric and cognitive phenotypes and three diffusion modalities (FA, MD, AD; column panels). Same significance classification and FDR correction scheme as above, applied across all modality × phenotype × network-pair cells.

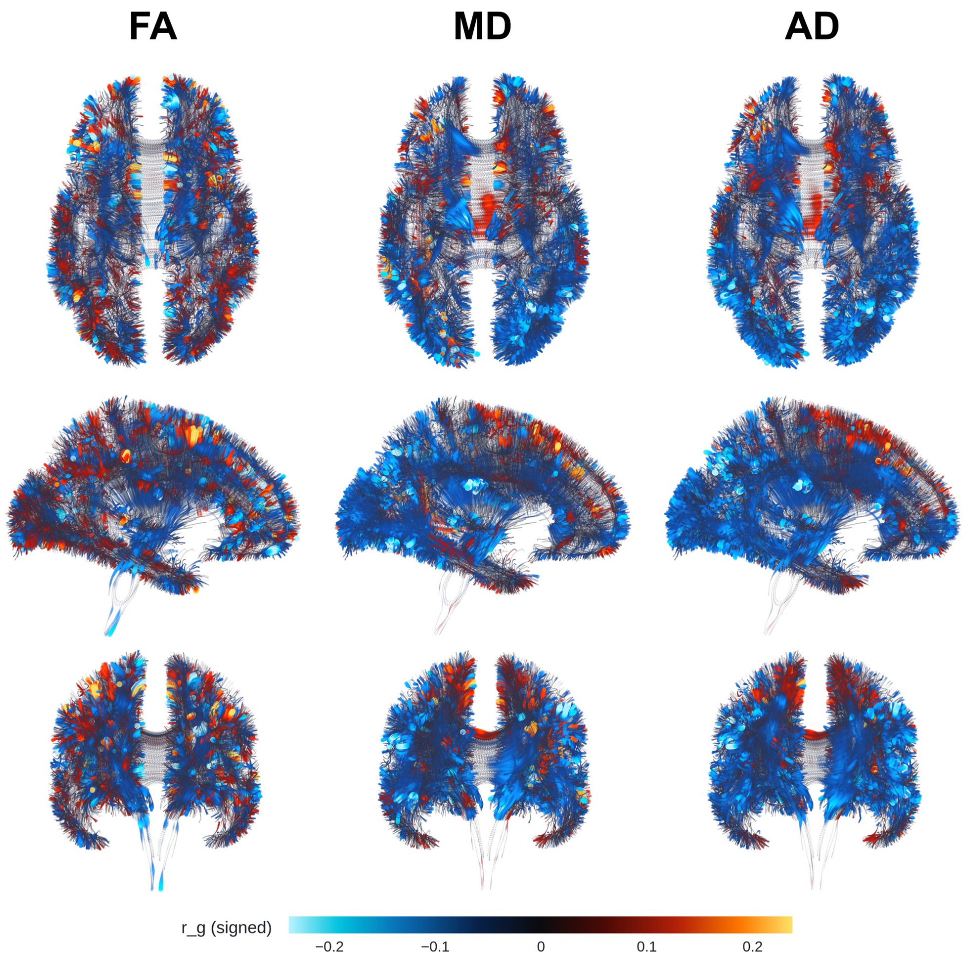

**Supplementary Fig. 9**: **Genetic correlation between white matter microstructure and ADHD.** Per-streamline genetic correlations ($r_g$) between fractional anisotropy (FA), mean diffusivity (MD), and axial diffusivity (AD) (columns, left to right) and attention-deficit/hyperactivity disorder (ADHD) risk are shown on the ENIGMA FA template, in axial, sagittal, and coronal view (rows, top to bottom). Color indicates signed $r_g$ (colorbar, bottom); the color scale is independent per figure. GWAS summary statistics were drawn from [81] ($n$ = 225,534; 38,691 cases and 186,843 controls, from cohorts that do not include UK Biobank). Voxel-level correlations are shown without additional thresholding; tract-level significance (Benjamini–Hochberg FDR, $q < 0.05$, applied jointly across all tract × trait × modality cells; §1.10) is summarized in Supplementary Table 4 and Supplementary Fig. 7. ADHD shows the weakest tract-level FA signal of the panel (4 of 17 tracts), strongest at the anterior corona radiata (ACR; $z = -4.04$); the signal is far more extensive in MD (8 of 17 tracts) and AD (9 of 17), both strongest along the superior longitudinal fasciculus (SLF) and sagittal stratum (SS; MD $z = 38.93$ and 29.13; AD $z = -9.99$ and $-6.67$), with the largest single association at the posterior thalamic radiation in AD (PTR; $z = -12.40$).

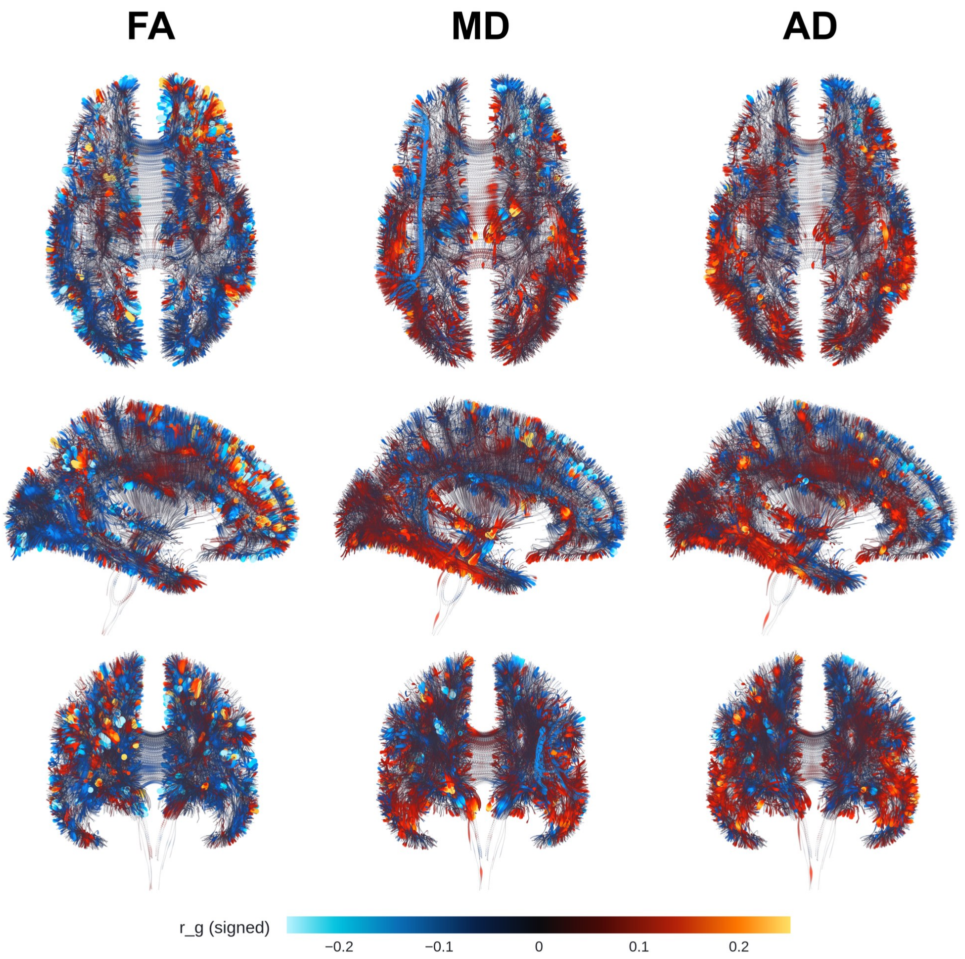

**Supplementary Fig. 10**: **Genetic correlation between white matter microstructure and ASD.** Per-streamline genetic correlations ($r_g$) between FA, MD, and AD (columns, left to right) and autism spectrum disorder (ASD) risk, axial/sagittal/ coronal views (rows, top to bottom), ENIGMA FA template. Color indicates signed $r_g$ (colorbar, bottom); scale independent per figure. GWAS summary statistics were drawn from [85] ($n$ = 46,350; 18,381 cases and 27,969 controls, ascertained from a Danish population with no UK Biobank overlap). Voxel-level correlations are shown without additional thresholding; tract-level significance (Benjamini–Hochberg FDR, $q < 0.05$, applied jointly across all tract × trait × modality cells; §1.10) is summarized in Supplementary Table 4 and Supplementary Fig. 7. ASD shows no FDR-significant tract-level correlation in FA, the most spatially restricted signal of any trait in this panel; MD shows a broader, if still modest, pattern (5 of 17 tracts), strongest along the superior longitudinal fasciculus and splenium of the corpus callosum (SLF, $z = -52.46$; SCC, $z = 43.62$; sagittal stratum, SS, $z = -31.43$), while AD reaches significance only at the sagittal stratum (SS; $z = 2.33$).

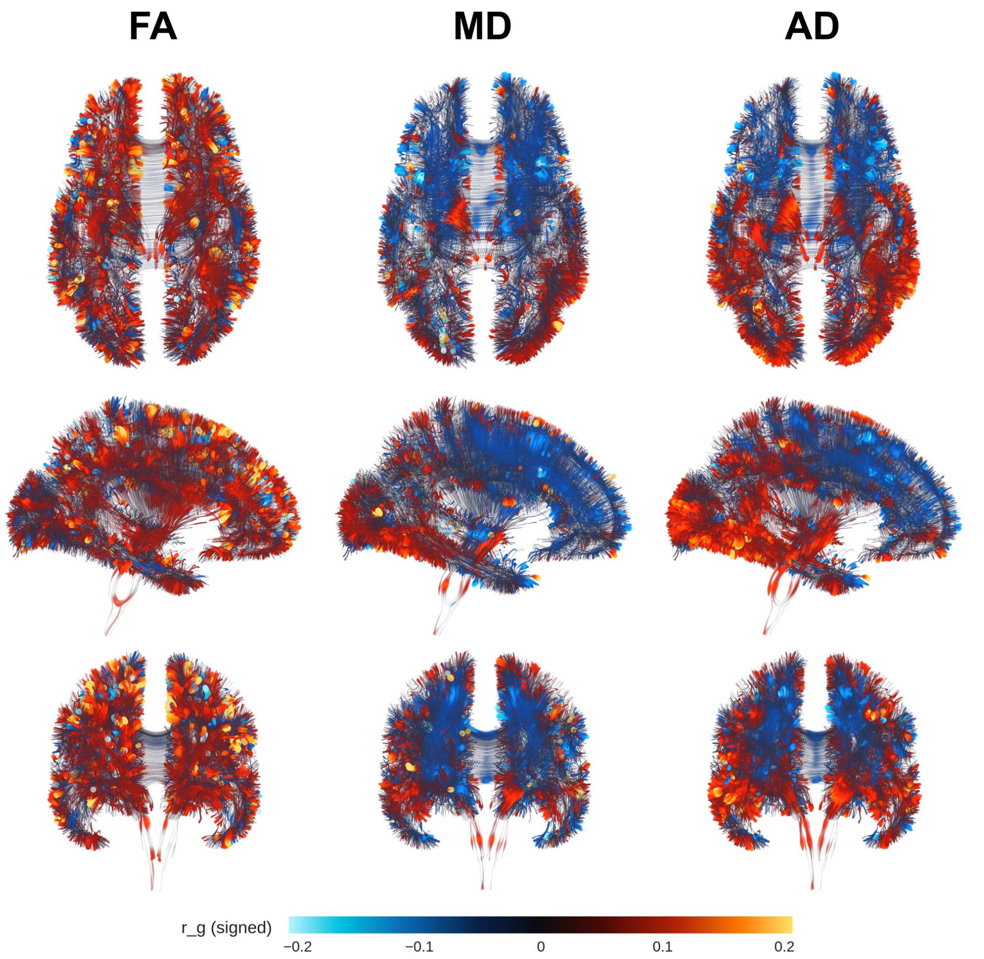

**Supplementary Fig. 11**: **Genetic correlation between white matter microstructure and cognitive performance (CP).** Per-streamline genetic correlations ($r_g$) between FA, MD, and AD (columns, left to right) and cognitive performance (CP), axial/sagittal/coronal views (rows, top to bottom), ENIGMA FA template. Color indicates signed $r_g$ (colorbar, bottom); scale independent per figure. GWAS summary statistics were drawn from [88] ($n$ = 257,828); this GWAS includes UK Biobank participants, so shared sample overlap with the present imaging cohort may modestly inflate the estimated genetic correlations. Voxel-level correlations are shown without additional thresholding; tract-level significance (Benjamini–Hochberg FDR, $q < 0.05$, applied jointly across all tract × trait × modality cells; §1.10) is summarized in Supplementary Table 4 and Supplementary Fig. 7. Cognitive performance shows broad, FDR-significant FA correlation across 15 of 17 tracts, strongest at the superior longitudinal fasciculus (SLF; $z = 16.84$), anterior corona radiata (ACR; $z = 14.41$), and genu of the corpus callosum (GCC; $z = 11.51$); MD (6 of 17 tracts) and AD (10 of 17) are strongest at the SLF/splenium (MD $z = -28.45/22.44$) and posterior thalamic radiation (AD, PTR; $z = 8.30$), respectively. Cognitive performance derives from the same discovery GWAS as educational attainment (Supplementary Fig. 19) and is genetically correlated with intelligence (Supplementary Fig. 20); the concordant regional pattern across all three traits likely reflects this shared genetic architecture rather than independent evidence.

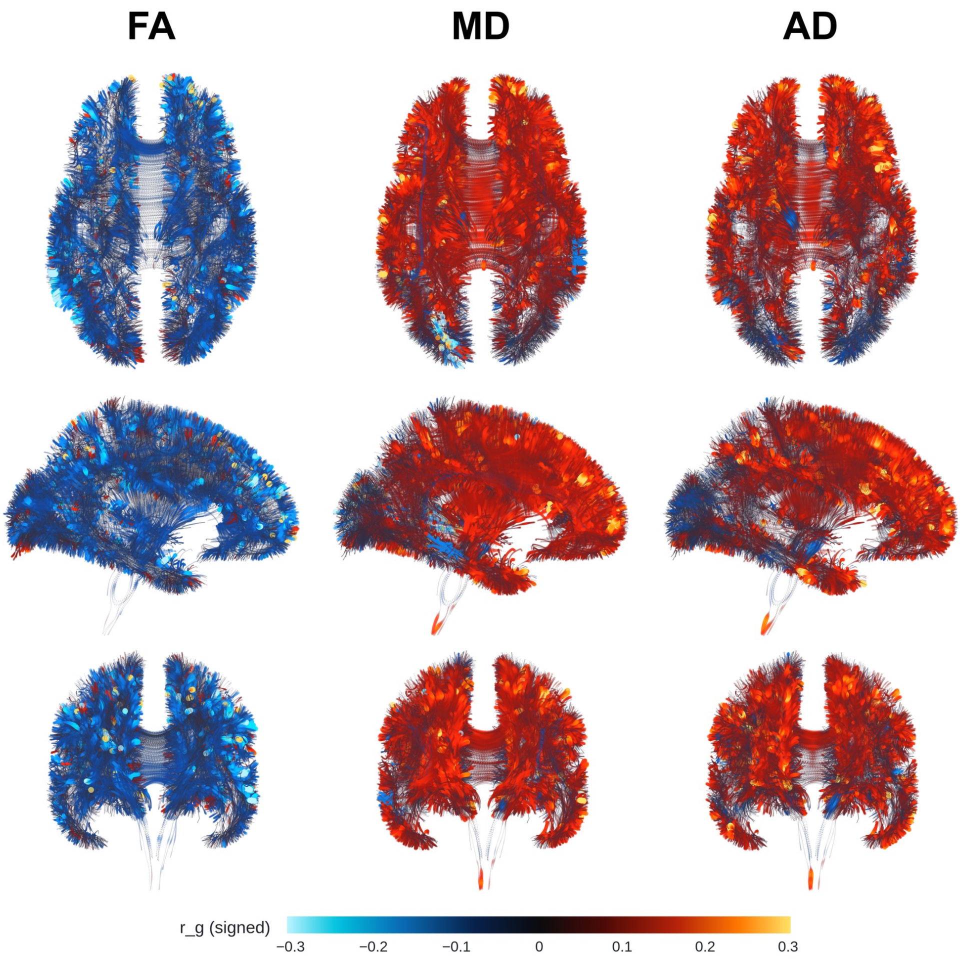

**Supplementary Fig. 12**: **Genetic correlation between white matter microstructure and depression.** Per-streamline genetic correlations ($r_g$) between FA, MD, and AD (columns, left to right) and depression, axial/sagittal/coronal views (rows, top to bottom), ENIGMA FA template. Color indicates signed $r_g$ (colorbar, bottom); scale independent per figure. GWAS summary statistics for a continuous, item-derived depression phenotype were drawn from [90] ($n$ = 381,455); this phenotype is distinct from the clinically ascertained major depressive disorder (MDD) case-control phenotype analyzed separately (Supplementary Fig. 14), and its direct UK Biobank sampling means genetic-correlation estimates here are more susceptible to sample-overlap inflation than the MDD estimates. Voxel-level correlations are shown without additional thresholding; tract-level significance (Benjamini–Hochberg FDR, $q < 0.05$, applied jointly across all tract × trait × modality cells; §1.10) is summarized in Supplementary Table 4 and Supplementary Fig. 7. Depression shows FDR-significant FA correlation in 14 of 17 tracts, strongest at the superior longitudinal fasciculus (SLF; $z = -22.28$), genu (GCC; $z = -15.57$), and anterior corona radiata (ACR; $z = -15.48$); MD shows the broadest involvement (15 of 17 tracts), strongest at the splenium and sagittal stratum (SCC, $z = -47.99$; SS, $z = -29.58$), while AD (9 of 17) is strongest at the ACR ($z = 21.06$).

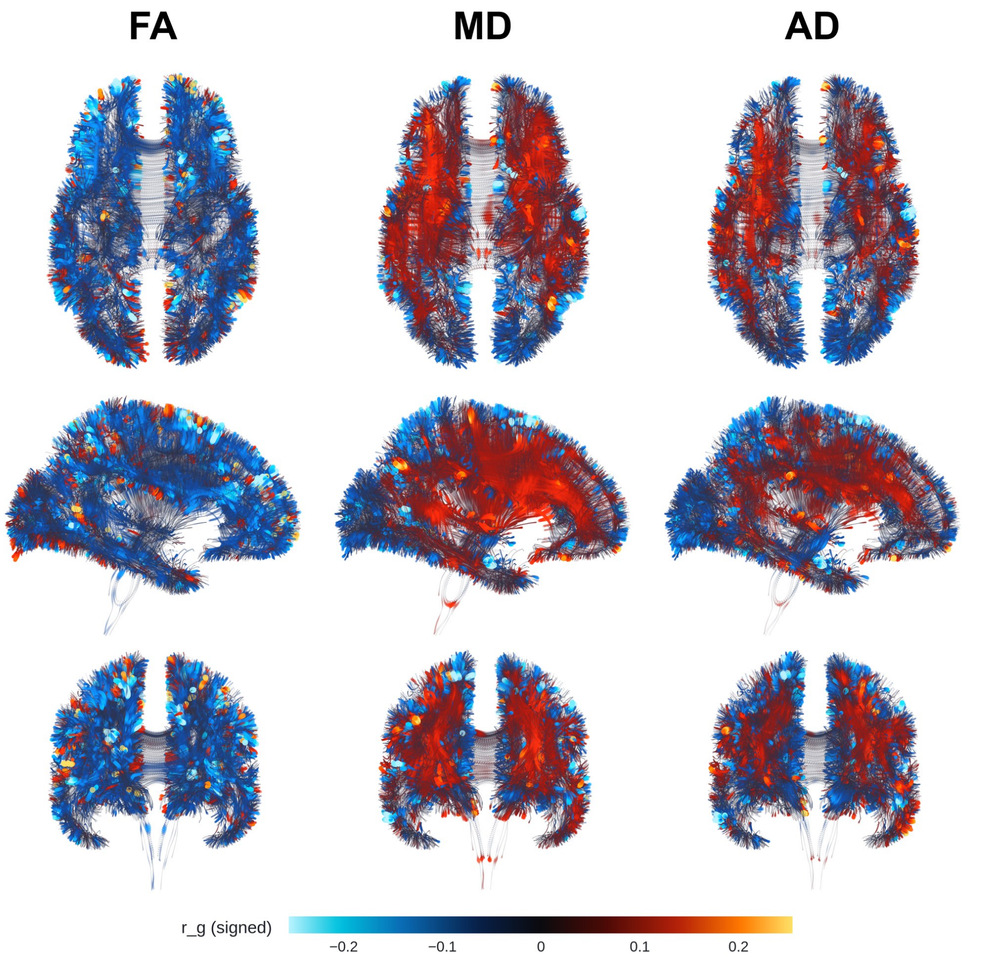

**Supplementary Fig. 13**: **Genetic correlation between white matter microstructure and insomnia.** Per-streamline genetic correlations ($r_g$) between FA, MD, and AD (columns, left to right) and insomnia, axial/sagittal/coronal views (rows, top to bottom), ENIGMA FA template. Color indicates signed $r_g$ (colorbar, bottom); scale independent per figure. GWAS summary statistics were drawn from [84] ($n = 386{,}988$; 109,548 cases and 277,440 controls); this UK Biobank-based cohort overlaps with the present imaging sample, which may modestly inflate the estimated genetic correlations. Voxel-level correlations are shown without additional thresholding; tract-level significance (Benjamini–Hochberg FDR, $q < 0.05$, applied jointly across all tract × trait × modality cells; §1.10) is summarized in Supplementary Table 4 and Supplementary Fig. 7. Insomnia shows FDR-significant FA correlation in 11 of 17 tracts, strongest at the anterior corona radiata (ACR; $z = -17.28$), superior longitudinal fasciculus (SLF; $z = -13.28$), and genu of the corpus callosum (GCC; $z = -10.12$); MD (6 of 17 tracts) is strongest at the splenium (SCC; $z = -41.02$), and AD (5 of 17) at the SLF ($z = 6.35$).

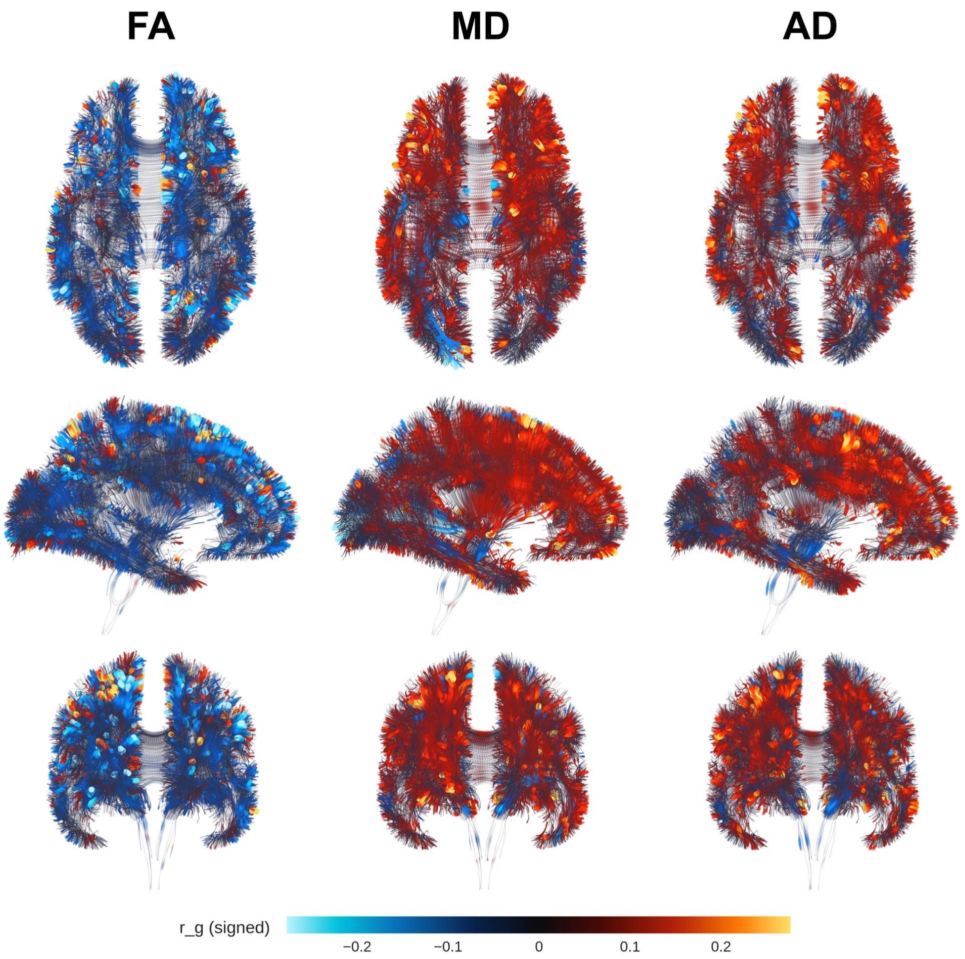

**Supplementary Fig. 14**: **Genetic correlation between white matter microstructure and major depressive disorder (MDD).** Per-streamline genetic correlations ($r_g$) between FA, MD, and AD (columns, left to right) and major depressive disorder (MDD), axial/sagittal/coronal views (rows, top to bottom), ENIGMA FA template. Color indicates signed $r_g$ (colorbar, bottom); scale independent per figure. GWAS summary statistics were drawn from [83] ($n$ = 480,359; 135,458 cases and 344,901 controls, drawn from PGC cohorts that exclude UK Biobank). Voxel-level correlations are shown without additional thresholding; tract-level significance (Benjamini–Hochberg FDR, $q < 0.05$, applied jointly across all tract × trait × modality cells; §1.10) is summarized in Supplementary Table 4 and Supplementary Fig. 7. MDD shows FDR-significant FA correlation in 8 of 17 tracts, strongest at the superior longitudinal fasciculus (SLF; $z = -4.50$) and anterior corona radiata (ACR; $z = -4.28$); MD (7 of 17 tracts) is strongest at the splenium (SCC; $z = 68.72$) and posterior thalamic radiation (PTR; $z = -45.04$), and AD (4 of 17) is strongest at the SLF and ACR ($z = 8.28$ and 8.12). Although the MDD and depression (Supplementary Fig. 12) discovery cohorts do not overlap in participants, the two phenotypes are highly genetically correlated, non-independent measures of depressive illness, so the largely concordant regional pattern between them reflects shared genetic liability rather than independent replication.

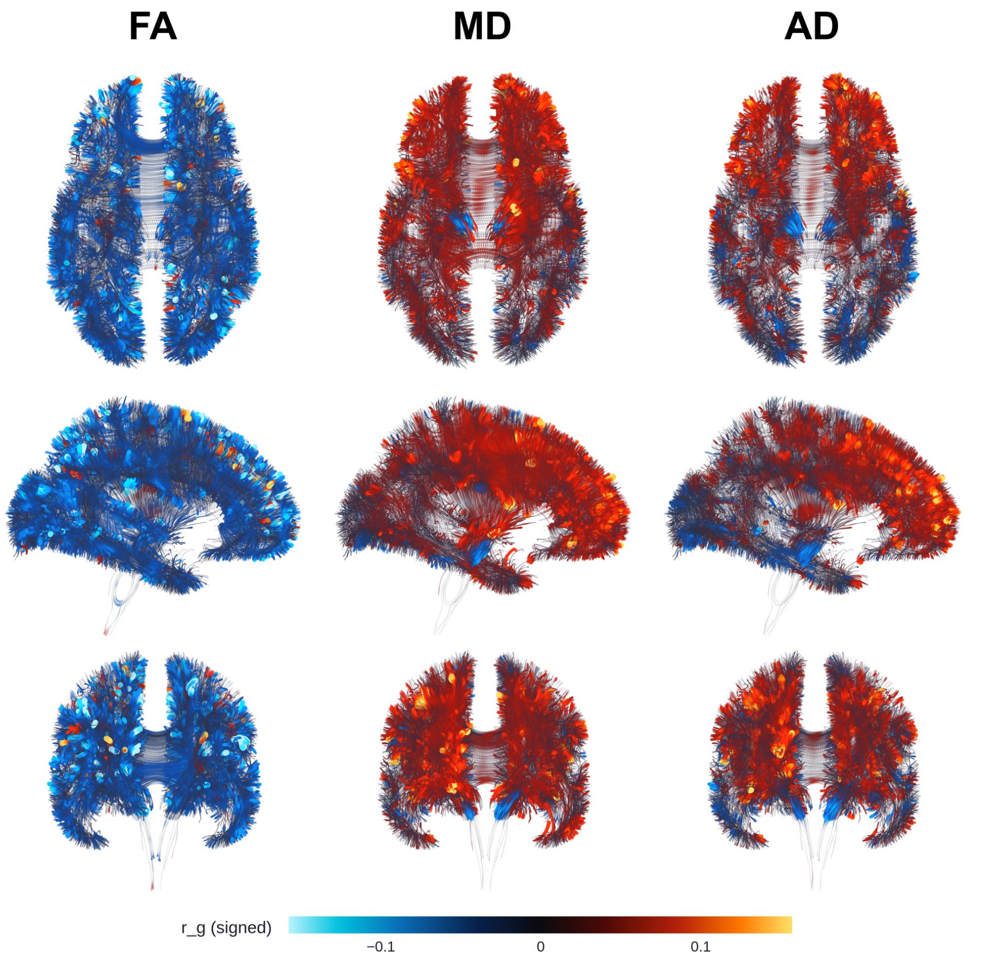

**Supplementary Fig. 15**: **Genetic correlation between white matter microstructure and schizophrenia (SCZ).** Per-streamline genetic correlations ($r_g$) between FA, MD, and AD (columns, left to right) and schizophrenia (SCZ) risk, axial/sagittal/coronal views (rows, top to bottom), ENIGMA FA template. Color indicates signed $r_g$ (colorbar, bottom); scale independent per figure. GWAS summary statistics were drawn from [86] ($n$ = 161,405; 67,390 cases and 94,015 controls, from an independent PGC cohort with no UK Biobank overlap). Voxel-level correlations are shown without additional thresholding; tract-level significance (Benjamini–Hochberg FDR, $q < 0.05$, applied jointly across all tract × trait × modality cells; §1.10) is summarized in Supplementary Table 4 and Supplementary Fig. 7. Schizophrenia shows the broadest tract-level FA involvement of any trait in the panel, reaching FDR significance in all 17 of 17 tracts, strongest at the superior longitudinal fasciculus (SLF; $z = -8.07$), anterior corona radiata (ACR; $z = -6.29$), and sagittal stratum (SS; $z = -5.82$); MD (12 of 17 tracts) is strongest at the splenium (SCC; $z = 22.01$) and posterior thalamic radiation (PTR; $z = 15.64$), and AD (9 of 17) at the ACR ($z = 10.57$).

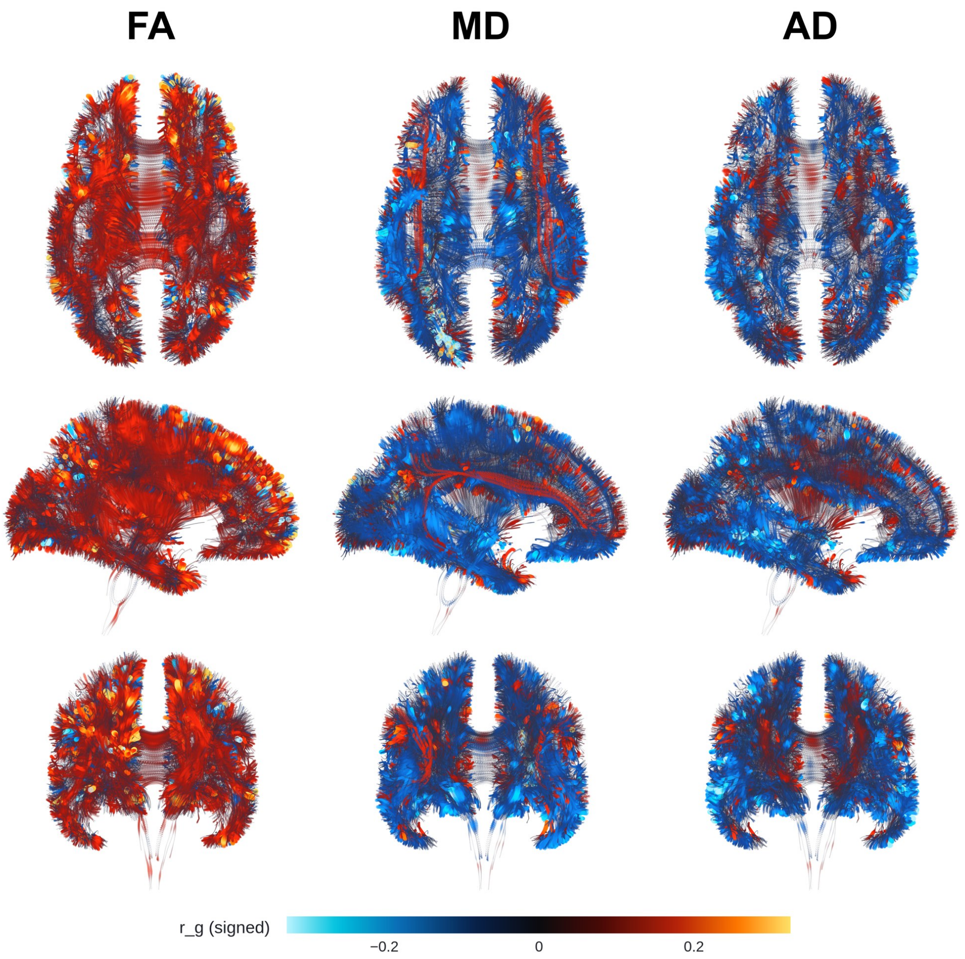

**Supplementary Fig. 16**: **Genetic correlation between white matter microstructure and Alzheimer's disease risk.** Per-streamline genetic correlations ($r_g$) between FA, MD, and AD (columns, left to right) and Alzheimer's disease risk (PGC-ALZ2, excluding 23andMe), axial/sagittal/coronal views (rows, top to bottom), ENIGMA FA template. Color indicates signed $r_g$ (colorbar, bottom); scale independent per figure. GWAS summary statistics were drawn from [87] ($n$ = 1,126,563; 90,338 cases and 1,036,225 controls); this meta-analysis includes UK Biobank participants, so estimated genetic correlations may be modestly inflated by sample overlap. Voxel-level correlations are shown without additional thresholding; tract-level significance (Benjamini–Hochberg FDR, $q < 0.05$, applied jointly across all tract $\times$ trait $\times$ modality cells; §1.10) is summarized in Supplementary Table 4 and Supplementary Fig. 7. Alzheimer's disease risk shows FDR-significant FA correlation in 15 of 17 tracts, strongest at the superior corona radiata (SCR; $z = 35.49$), superior longitudinal fasciculus (SLF; $z = 26.77$), and anterior corona radiata (ACR; $z = 20.18$); MD (11 of 17 tracts) is strongest at the SLF ($z = 62.73$), and AD (9 of 17) at the sagittal stratum (SS; $z = -8.84$).

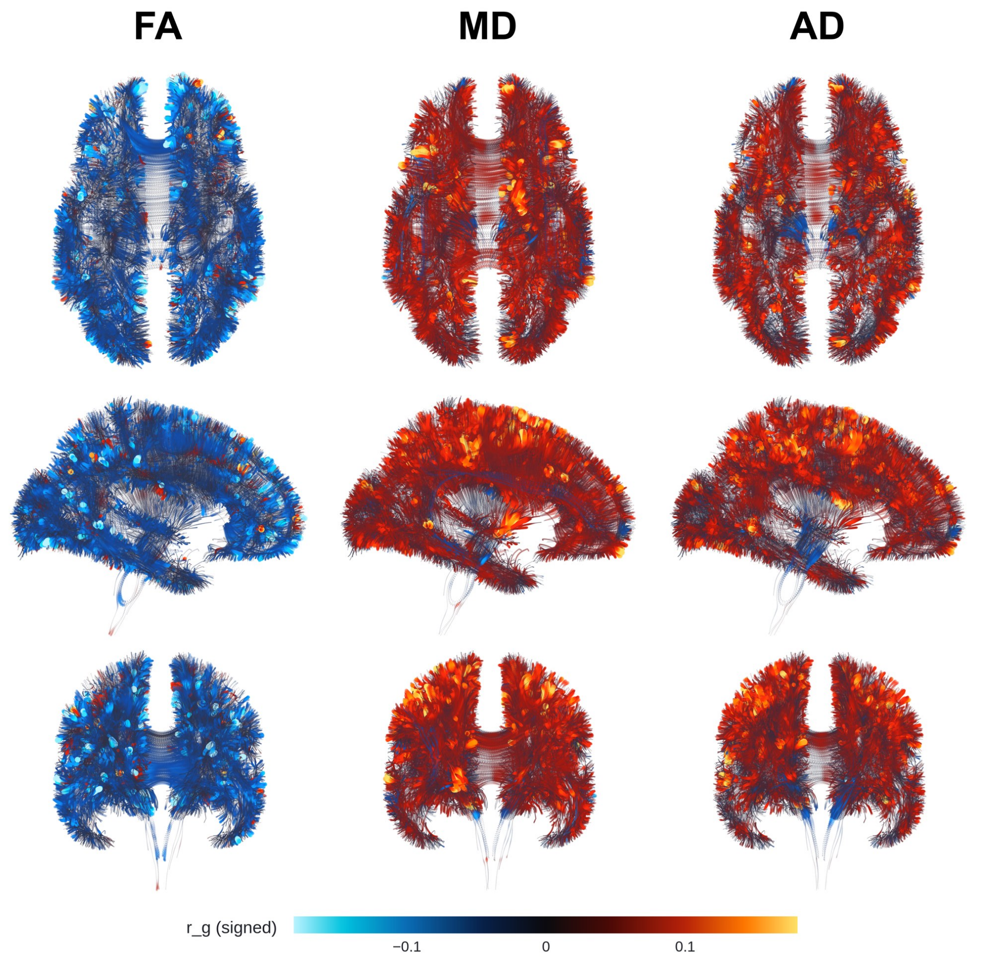

**Supplementary Fig. 17**: **Genetic correlation between white matter microstructure and bipolar disorder.** Per-streamline genetic correlations ($r_g$) between FA, MD, and AD (columns, left to right) and bipolar disorder risk, axial/sagittal/coronal views (rows, top to bottom), ENIGMA FA template. Color indicates signed $r_g$ (colorbar, bottom); scale independent per figure. GWAS summary statistics were drawn from [82] ($n$ = 413,466; 41,917 cases and 371,549 controls, from PGC cohorts that exclude UK Biobank). Voxel-level correlations are shown without additional thresholding; tract-level significance (Benjamini–Hochberg FDR, $q < 0.05$, applied jointly across all tract × trait × modality cells; §1.10) is summarized in Supplementary Table 4 and Supplementary Fig. 7. Bipolar disorder shows FDR-significant FA correlation in 13 of 17 tracts, strongest at the superior longitudinal fasciculus (SLF; $z = -6.44$), anterior corona radiata (ACR; $z = -5.24$), and sagittal stratum (SS; $z = -5.00$), the same tract most strongly implicated in schizophrenia (Supplementary Fig. 15); MD (12 of 17 tracts) is strongest at the SLF ($z = -47.34$), and AD (11 of 17) likewise at the SLF ($z = 11.90$).

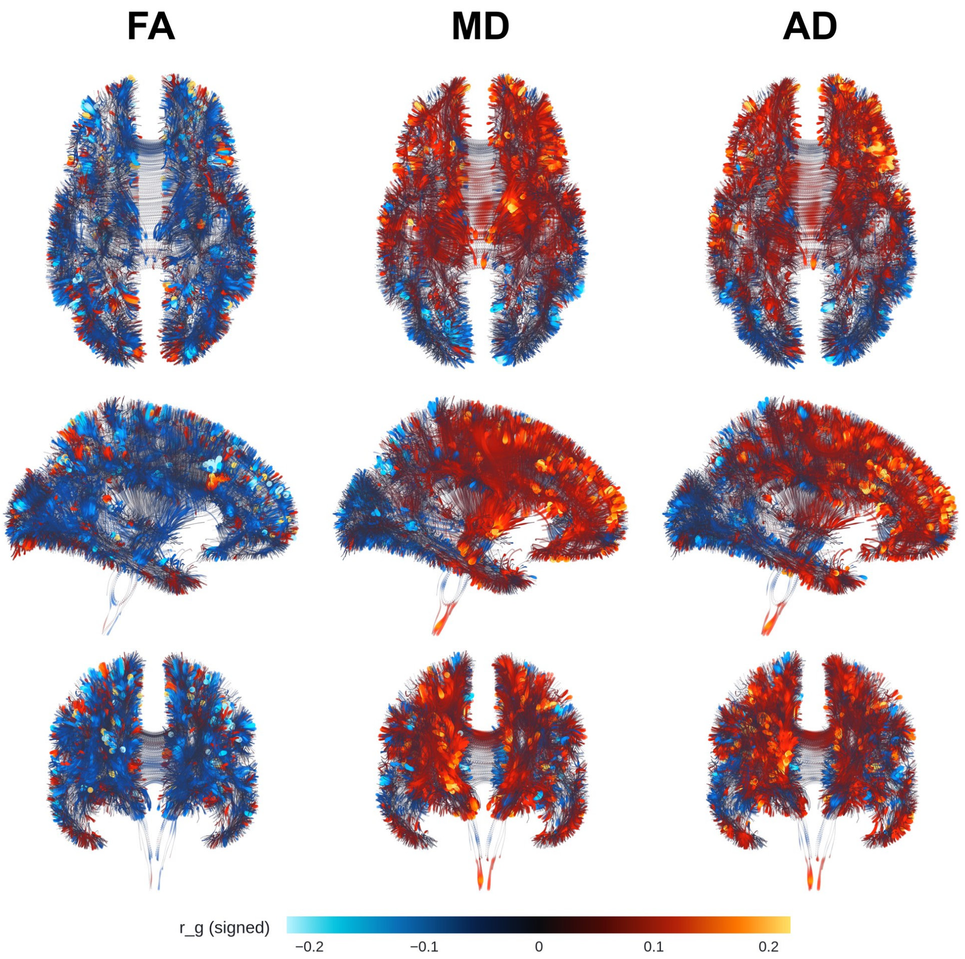

**Supplementary Fig. 18**: **Genetic correlation between white matter microstructure and depressed affect.** Per-streamline genetic correlations ($r_g$) between FA, MD, and AD (columns, left to right) and depressed affect, axial/sagittal/-coronal views (rows, top to bottom), ENIGMA FA template. Color indicates signed $r_g$ (colorbar, bottom); scale independent per figure. GWAS summary statistics for this depressed-affect subcluster phenotype, derived from the same large-scale item-level GWAS as neuroticism and its other subclusters, were drawn from [90] ($n = 357{,}957$); this UK Biobank-based sample overlaps with the present imaging cohort, which may modestly inflate the estimated genetic correlations. Voxel-level correlations are shown without additional thresholding; tract-level significance (Benjamini–Hochberg FDR, $q < 0.05$, applied jointly across all tract × trait × modality cells; §1.10) is summarized in Supplementary Table 4 and Supplementary Fig. 7. Depressed affect shows FDR-significant FA correlation in 11 of 17 tracts, strongest at the superior longitudinal fasciculus (SLF; $z = -13.63$) and superior corona radiata (SCR; $z = -12.55$); MD (11 of 17 tracts) shows a markedly larger effect at the splenium of the corpus callosum than any other tract-trait pair in the panel (SCC; $z = -87.19$), and AD (7 of 17) is strongest at the anterior corona radiata (ACR; $z = 14.27$). The regional pattern broadly overlaps with depression (Supplementary Fig. 12) and neuroticism (Supplementary Fig. 21), consistent with depressed affect being drawn from the same broader depression/neuroticism item battery.

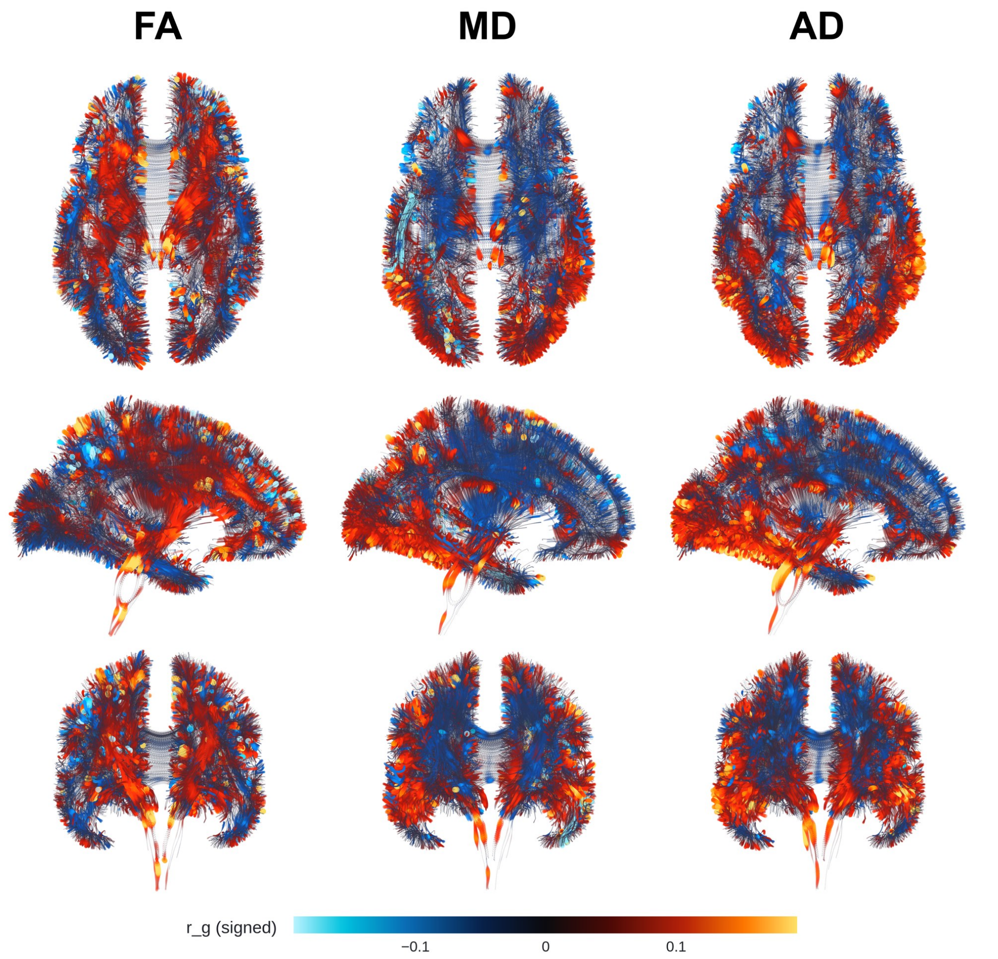

**Supplementary Fig. 19**: **Genetic correlation between white matter microstructure and educational attainment (EA).** Per-streamline genetic correlations ($r_g$) between FA, MD, and AD (columns, left to right) and educational attainment (EA), axial/sagittal/coronal views (rows, top to bottom), ENIGMA FA template. Color indicates signed $r_g$ (colorbar, bottom); scale independent per figure. GWAS summary statistics were drawn from [88] ($n$ = 766,345); this GWAS includes UK Biobank participants, so shared sample overlap with the present imaging cohort may modestly inflate the estimated genetic correlations. Voxel-level correlations are shown without additional thresholding; tract-level significance (Benjamini–Hochberg FDR, $q < 0.05$, applied jointly across all tract × trait × modality cells; §1.10) is summarized in Supplementary Table 4 and Supplementary Fig. 7. Educational attainment shows FDR-significant FA correlation in 13 of 17 tracts, strongest at the superior corona radiata (SCR; $z$ = 11.01), anterior corona radiata (ACR; $z$ = 10.99), and superior longitudinal fasciculus (SLF; $z$ = 9.47); MD (8 of 17 tracts) is strongest at the SLF ($z$ = −51.74), and AD (10 of 17) at the posterior thalamic radiation (PTR; $z$ = 14.38). Educational attainment derives from the same discovery GWAS as cognitive performance (Supplementary Fig. 11) and is genetically correlated with intelligence (Supplementary Fig. 20); the similar regional pattern across all three traits reflects this shared cognitive genetic architecture.

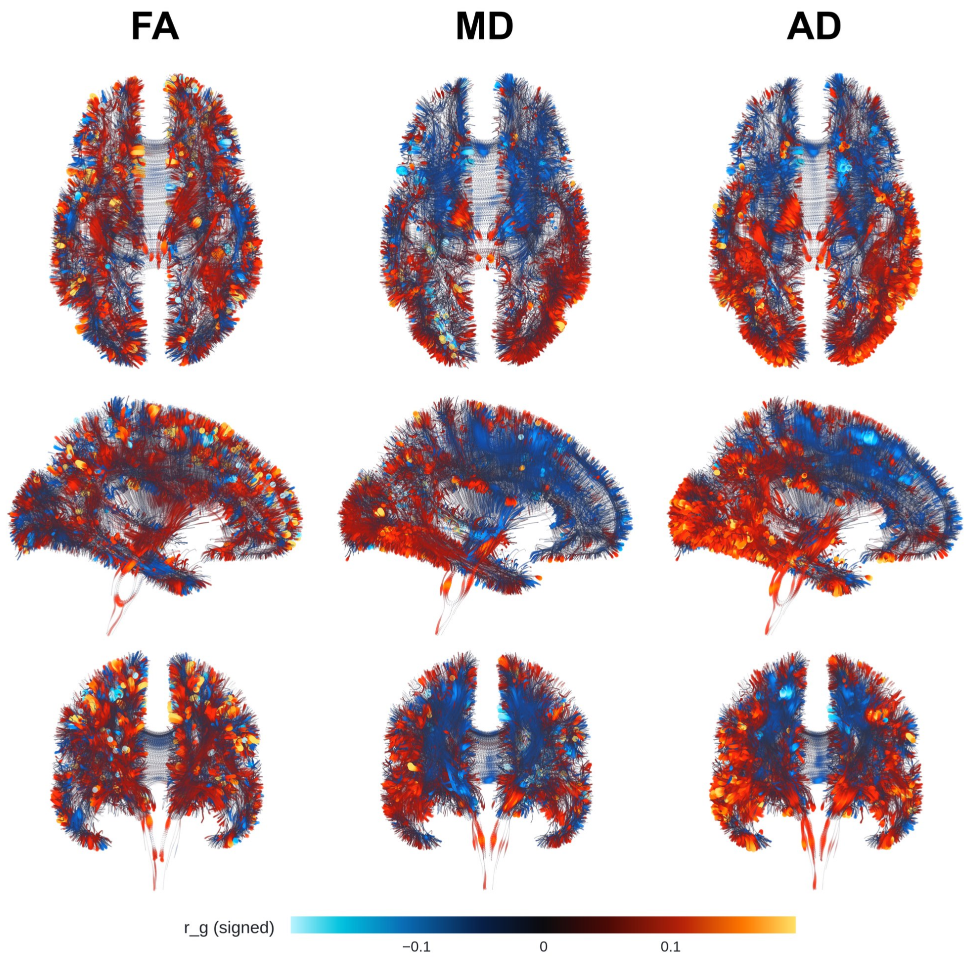

**Supplementary Fig. 20**: **Genetic correlation between white matter microstructure and intelligence.** Per-streamline genetic correlations ($r_g$) between FA, MD, and AD (columns, left to right) and intelligence, axial/sagittal/coronal views (rows, top to bottom), ENIGMA FA template. Color indicates signed $r_g$ (colorbar, bottom); scale independent per figure. GWAS summary statistics were drawn from [89] ($n$ = 269,867); this GWAS includes UK Biobank participants, so shared sample overlap with the present imaging cohort may modestly inflate the estimated genetic correlations. Voxel-level correlations are shown without additional thresholding; tract-level significance (Benjamini–Hochberg FDR, $q < 0.05$, applied jointly across all tract $\times$ trait $\times$ modality cells; §1.10) is summarized in Supplementary Table 4 and Supplementary Fig. 7. Intelligence shows FDR-significant FA correlation in 8 of 17 tracts, strongest at the superior longitudinal fasciculus (SLF; $z = 9.69$), anterior corona radiata (ACR; $z = 8.57$), and genu of the corpus callosum (GCC; $z = 6.23$); MD and AD show broader involvement (11 of 17 tracts each), strongest at the SLF in MD ($z = -35.31$) and the posterior thalamic radiation in AD (PTR; $z = 12.10$). The regional pattern closely parallels cognitive performance (Supplementary Fig. 11) and educational attainment (Supplementary Fig. 19), consistent with the well-established genetic correlation among these cognitive traits.

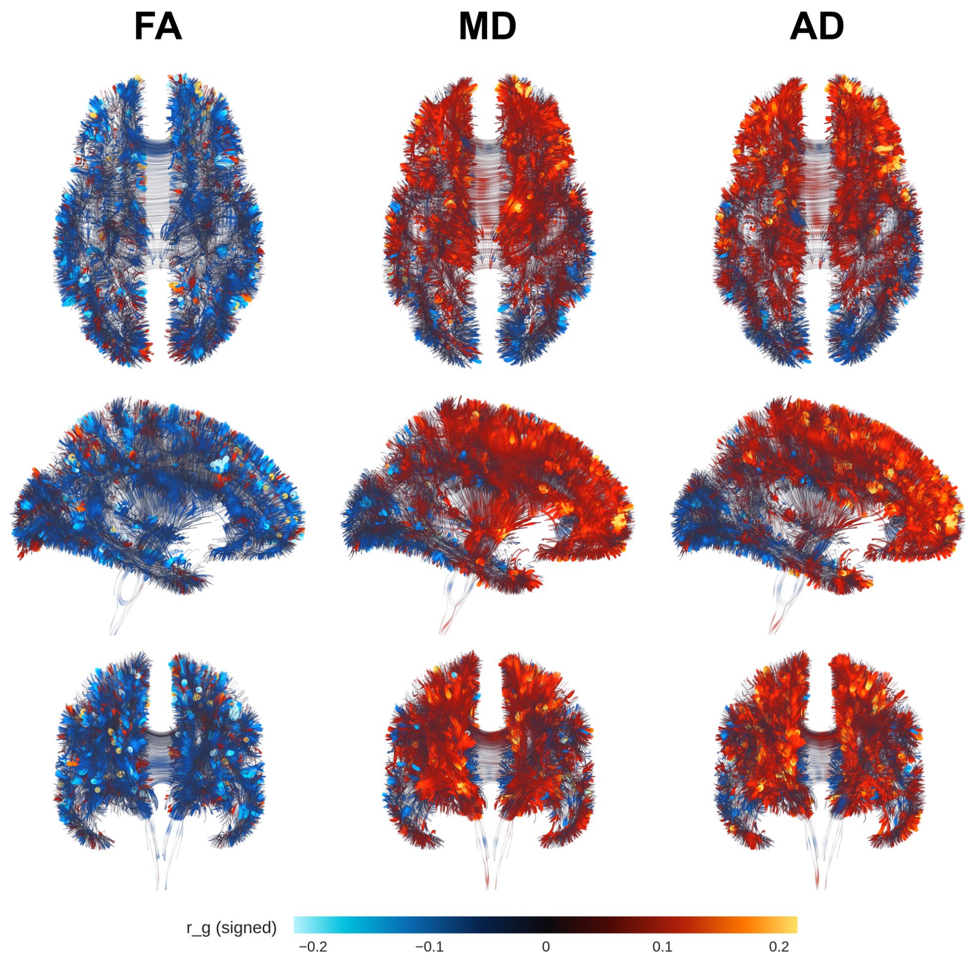

**Supplementary Fig. 21**: **Genetic correlation between white matter microstructure and neuroticism.** Per-streamline genetic correlations ($r_g$) between FA, MD, and AD (columns, left to right) and neuroticism, axial/sagittal/coronal views (rows, top to bottom), ENIGMA FA template. Color indicates signed $r_g$ (colorbar, bottom); scale independent per figure. GWAS summary statistics were drawn from [90] ($n$ = 390,278); this UK Biobank-based sample overlaps with the present imaging cohort, which may modestly inflate the estimated genetic correlations. Voxel-level correlations are shown without additional thresholding; tract-level significance (Benjamini–Hochberg FDR, $q < 0.05$, applied jointly across all tract × trait × modality cells; §1.10) is summarized in Supplementary Table 4 and Supplementary Fig. 7. Neuroticism shows FDR-significant FA correlation in 11 of 17 tracts, strongest at the superior longitudinal fasciculus (SLF; $z = -17.02$), anterior corona radiata (ACR; $z = -14.73$), and superior corona radiata (SCR; $z = -13.81$); MD (12 of 17 tracts) is strongest at the splenium (SCC; $z = -26.94$), and AD (9 of 17) at the ACR ($z = 20.43$). Neuroticism, depressed affect (Supplementary Fig. 18), and worry (Supplementary Fig. 22) are drawn from overlapping item batteries in the same source GWAS, consistent with their similar regional patterns here.

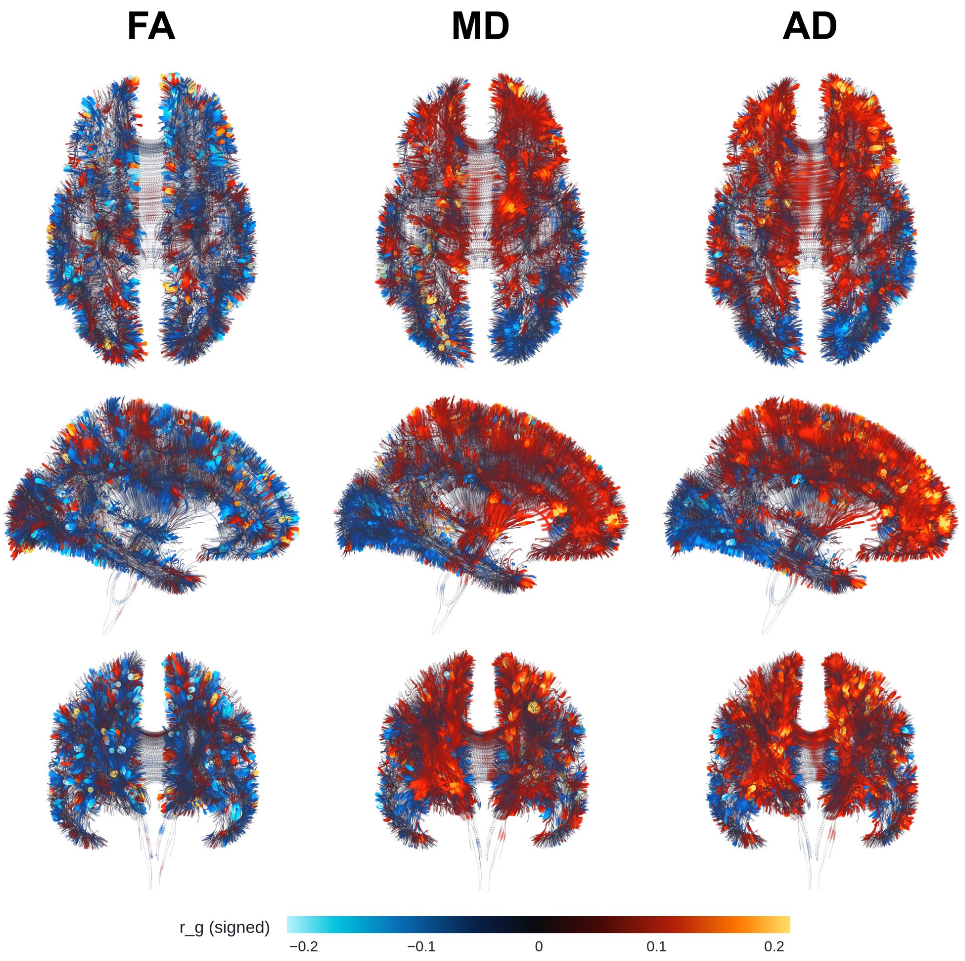

**Supplementary Fig. 22**: **Genetic correlation between white matter microstructure and worry.** Per-streamline genetic correlations ($r_g$) between FA, MD, and AD (columns, left to right) and worry, axial/sagittal/coronal views (rows, top to bottom), ENIGMA FA template. Color indicates signed $r_g$ (colorbar, bottom); scale independent per figure. GWAS summary statistics were drawn from [90] ($n$ = 348,219); this UK Biobank-based sample overlaps with the present imaging cohort, which may modestly inflate the estimated genetic correlations. Voxel-level correlations are shown without additional thresholding; tract-level significance (Benjamini–Hochberg FDR, $q < 0.05$, applied jointly across all tract × trait × modality cells; §1.10) is summarized in Supplementary Table 4 and Supplementary Fig. 7. Worry shows FDR-significant FA correlation in 5 of 17 tracts, the most restricted FA signal among the neuroticism-family phenotypes, strongest at the anterior corona radiata (ACR; $z = -5.93$) and genu of the corpus callosum (GCC; $z = -5.43$); MD (9 of 17 tracts) is strongest at the splenium (SCC; $z = 33.36$), and AD (8 of 17) at the ACR ($z = 17.68$). As with depressed affect, the regional pattern overlaps substantially with neuroticism (Supplementary Fig. 21), consistent with a shared source GWAS and item battery.

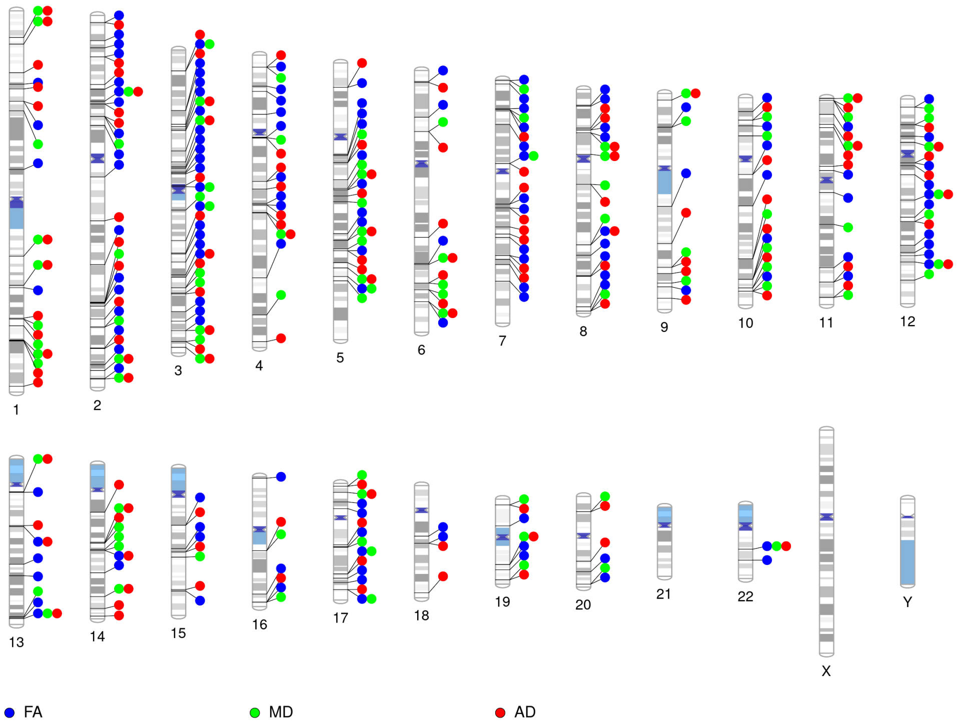

**Supplementary Fig. 23**: **White-matter loci distribute across every autosome, reflecting a polygenic architecture shared by all three diffusion measures.** Chromosomal ideograms of lead variants from the voxel-wise GWAS of fractional anisotropy (FA; 315 loci), mean diffusivity (MD; 238 loci), and axial diffusivity (AD; 345 loci). No single chromosome dominates any modality, consistent with a broadly polygenic architecture underlying variation in white-matter microstructure.

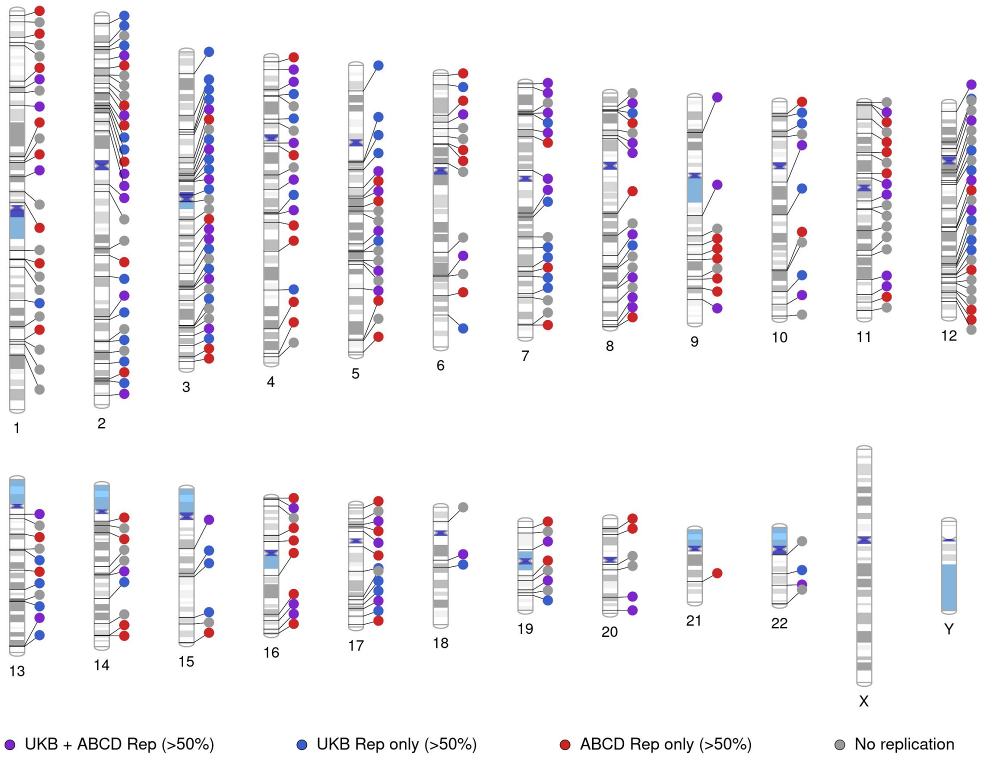

**Supplementary Fig. 24**: **FA loci discovered in UK Biobank adults are nominally reproduced across the genome in ABCD children.** Chromosomal ideogram comparing the genomic locations of FA lead variants identified in the UK Biobank discovery cohort with loci whose discovery footprint was reproduced in the ABCD cohort at a nominal threshold ($P < 10^{-2}$, models adjusted for acquisition site, $n = 7{,}403$; 149 of 315 loci with >50% spatial overlap, 72 of them also in UK Biobank Phase 7). Loci are colored according to replication status: red indicates replication in ABCD only, blue indicates replication in UK Biobank Phase 7 only, purple indicates replication in both cohorts, and gray indicates no replication in either cohort.

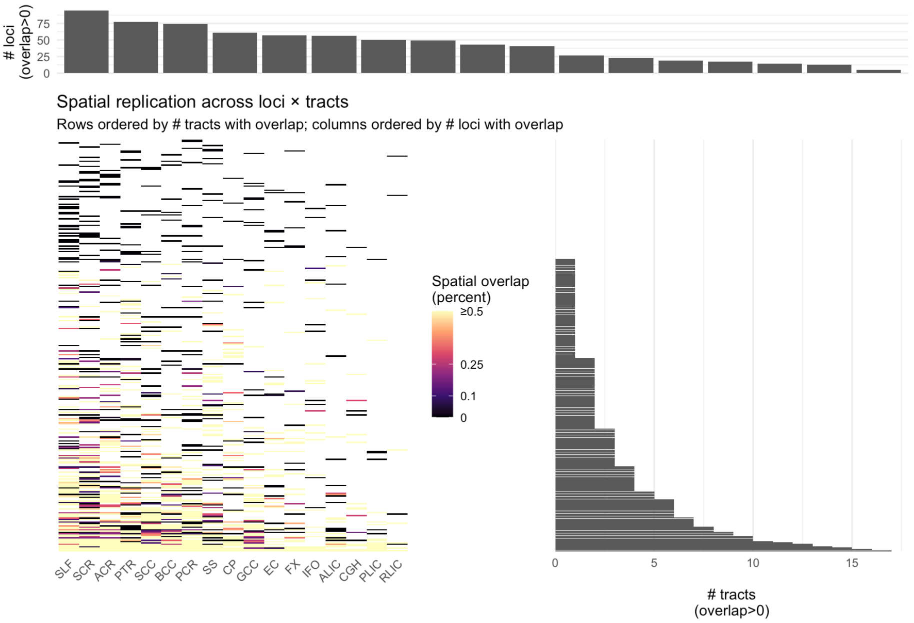

**Supplementary Fig. 25**: **Spatial footprints of discovered loci reproduce in the held-out cohort: 71% of the 315 FA loci overlap replication signals and 44% exceed 50% overlap.** Heatmap of spatial overlap between discovery loci and replication signals across major white-matter tracts. Each row is a locus and each column a tract; colors give the proportion of overlapping voxels between discovery and replication. Rows and columns are ordered by the number of tracts and loci showing overlap, with marginal bar plots summarizing these counts.

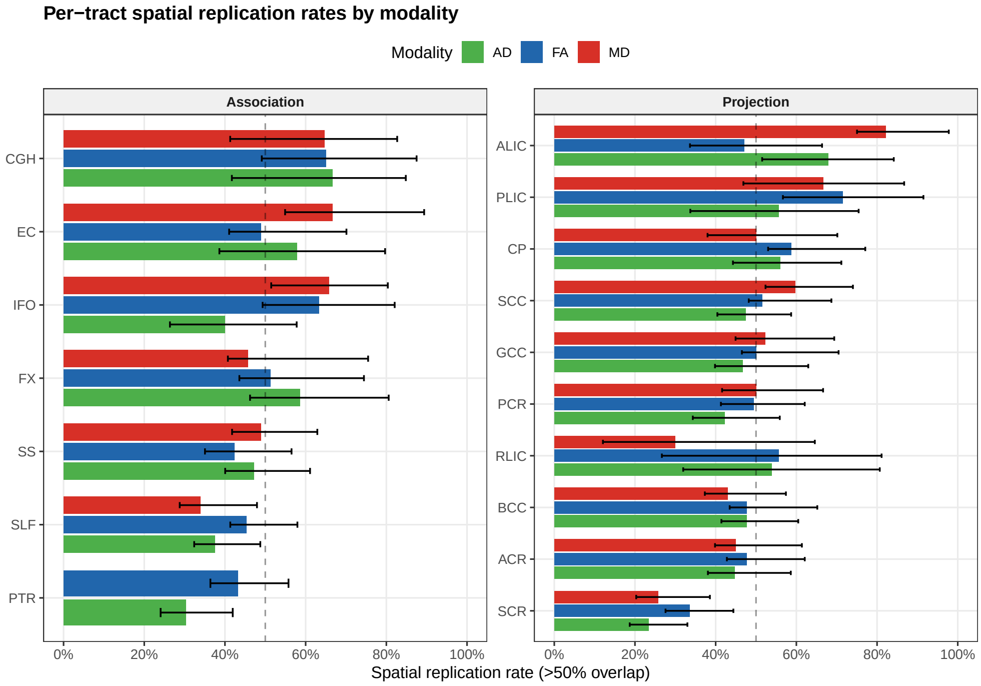

**Supplementary Fig. 26**: **Per-tract spatial replication rates by modality.** Fraction of discovery loci replicating with $> 50\%$ voxel overlap, shown per tract (mean $\pm$ 95% CI) and split into association tracts (left panel) and projection tracts (right panel), colored by modality (AD, FA, MD). Dashed line marks 50% replication for reference.

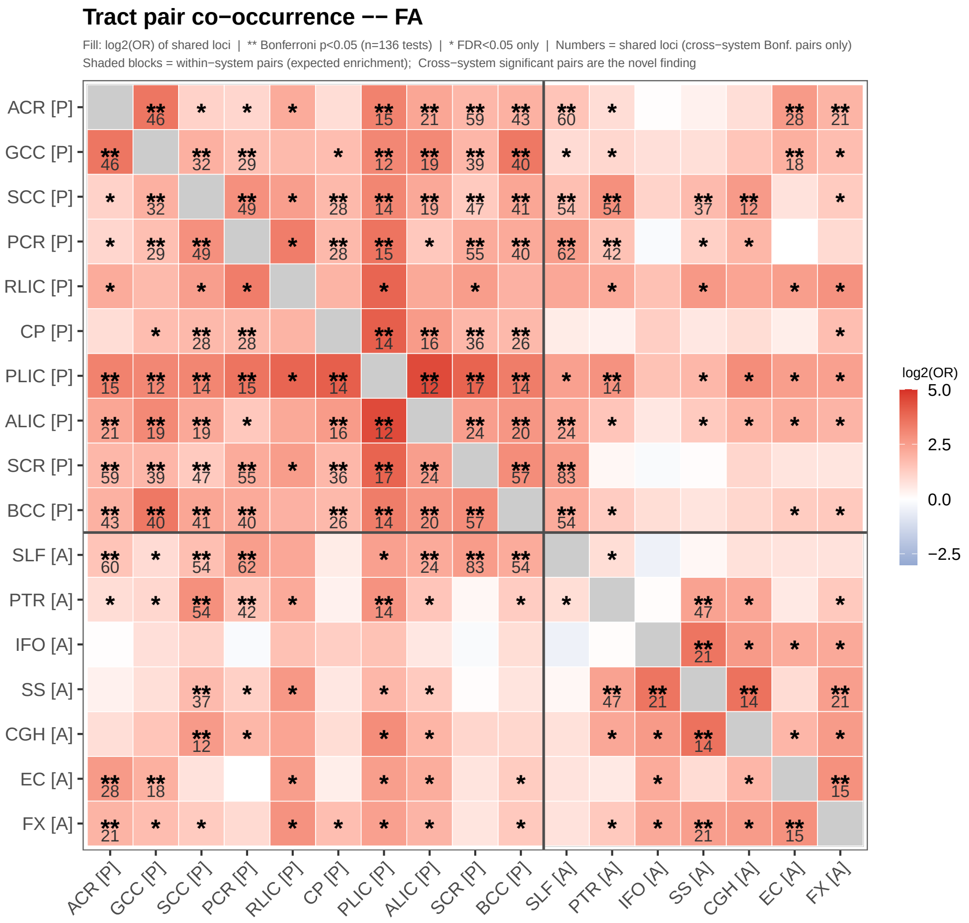

**Supplementary Fig. 27**: **Tract pair co-occurrence, FA.** Pairwise co-occurrence of discovery loci across 17 white-matter tract ROIs for fractional anisotropy (FA). Fill color shows log2(OR) of shared loci between each tract pair; ** marks Bonferroni-significant pairs ($p < 0.05$, n=136 tests), * marks FDR-significant-only pairs, and numbers give the shared-locus count for cross-system Bonferroni-significant pairs. Shaded blocks mark within-system (projection [P] vs. association [A]) pairs, where enrichment is expected; the significant cross-system pairs outside these blocks are the novel finding.

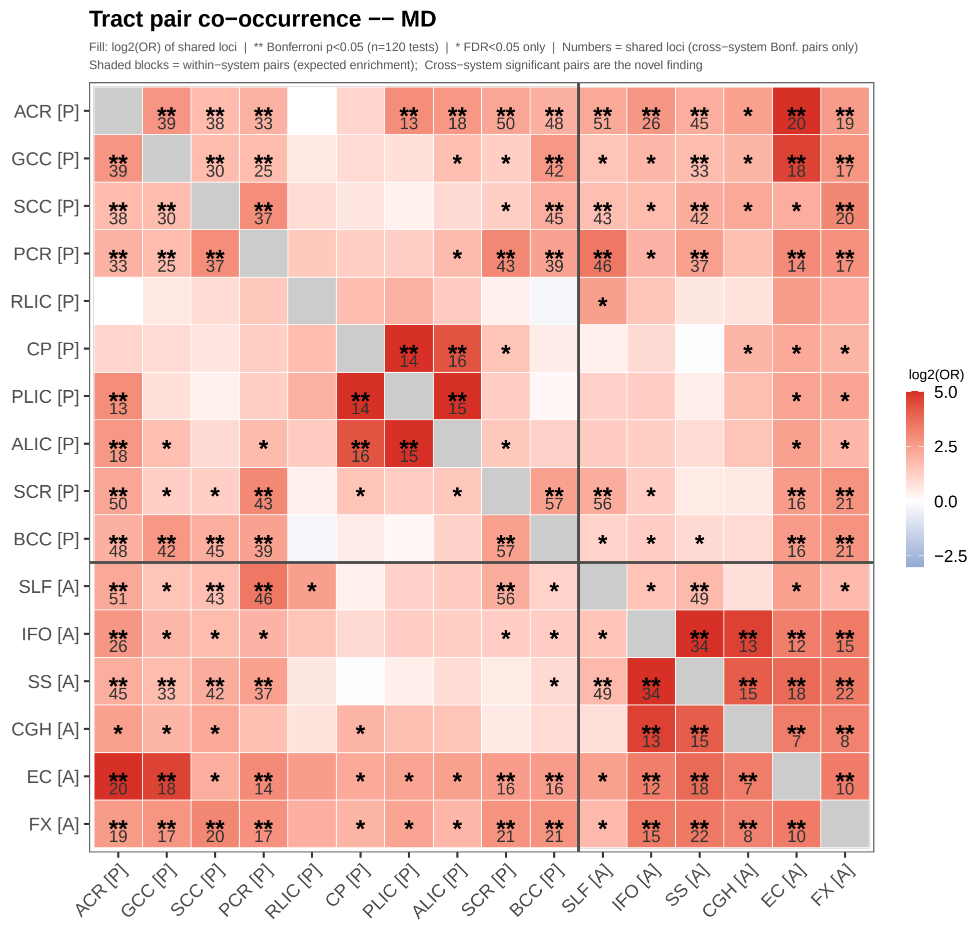

**Supplementary Fig. 28**: **Tract pair co-occurrence, MD.** Same as above, for mean diffusivity (MD). One fewer tract than the FA/AD panels (PTR is absent), giving 16 tracts and n=120 tests.

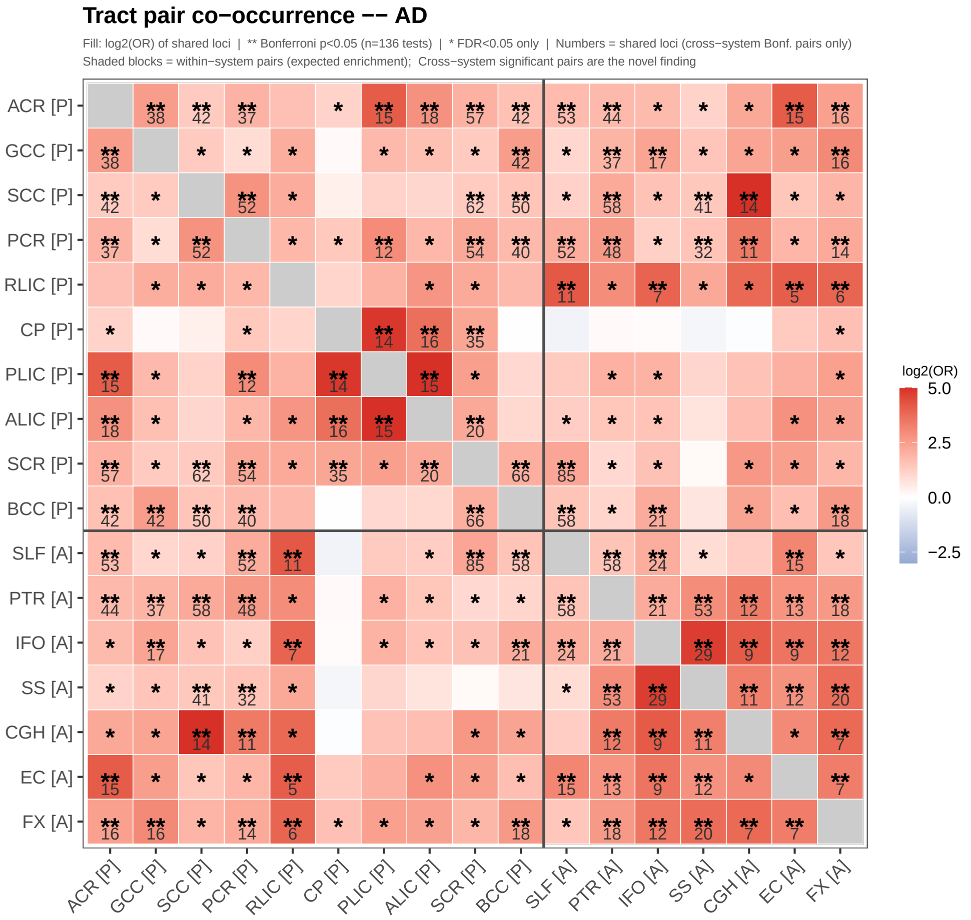

**Supplementary Fig. 29**: **Tract pair co-occurrence, AD.** Same as above, for axial diffusivity (AD); also 17 tracts, n=136 tests.

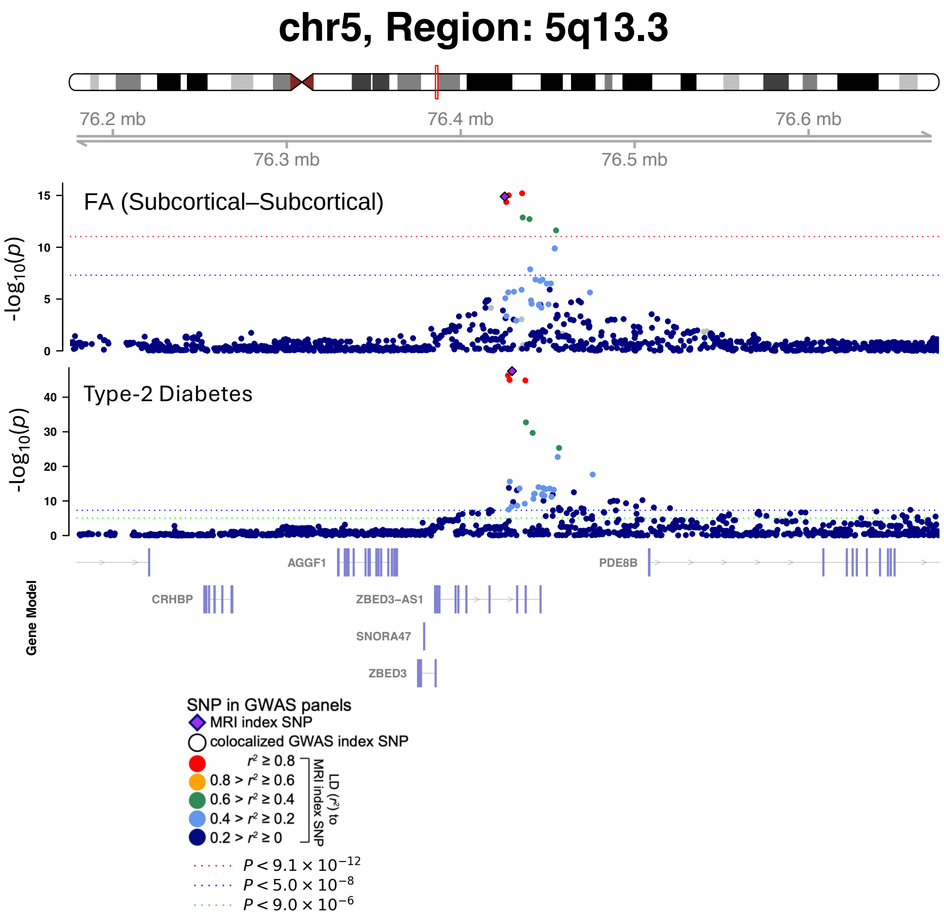

**Supplementary Fig. 30**: **The 5q13.3 white-matter locus shares a causal variant with type 2 diabetes (PP.H4 = 0.997).** Regional association plots for white-matter microstructure (FA at the representative location of the locus, which lies in fiber cluster 428 of the subcortical–subcortical functional network pair, a cluster not assigned to a JHU tract; the locus footprint spans the cerebral peduncle, posterior and superior corona radiata and posterior limb of the internal capsule; index variant rs4457053) and type 2 diabetes (index variant rs6878122) at 5q13.3. Bayesian colocalization supports a single shared causal variant (PP.H4.abf = 0.997).

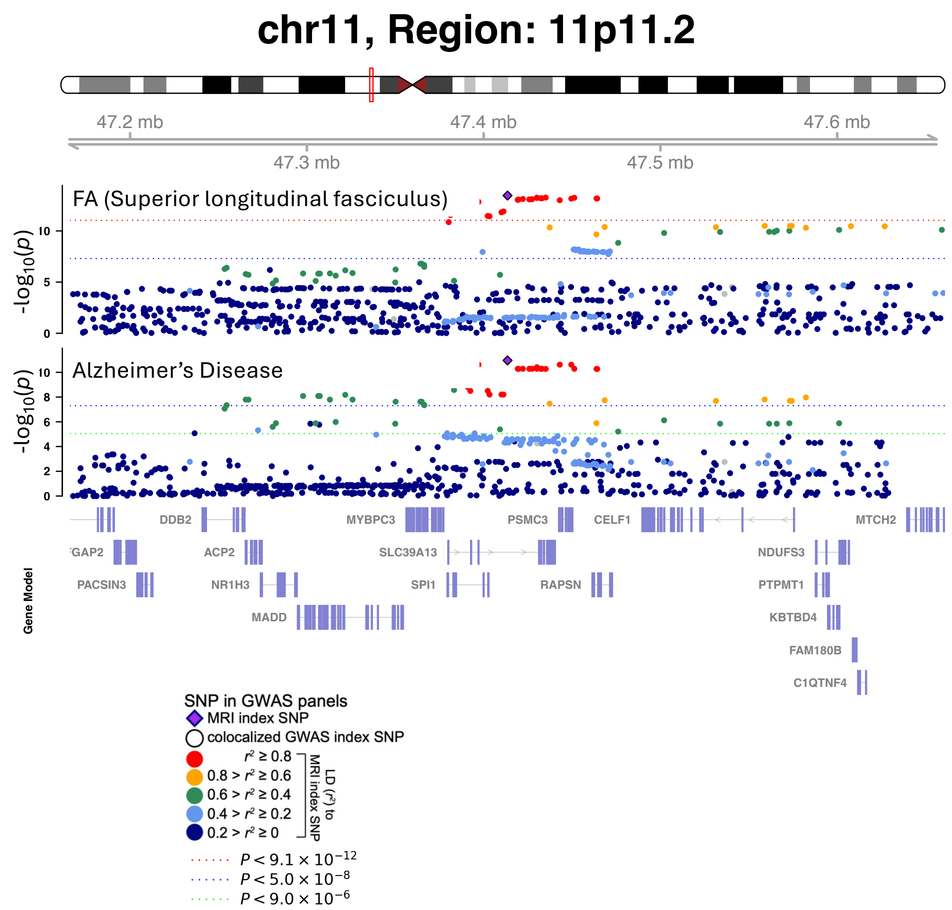

**Supplementary Fig. 31**: **The 11p11.2 white-matter locus shares a causal variant with Alzheimer's disease (PP.H4 = 0.982).** Regional association plots for white-matter microstructure (FA in the superior longitudinal fasciculus) and Alzheimer's disease at 11p11.2, with shared index variant rs10838702. Bayesian colocalization supports a single shared causal variant (PP.H4.abf = 0.982).

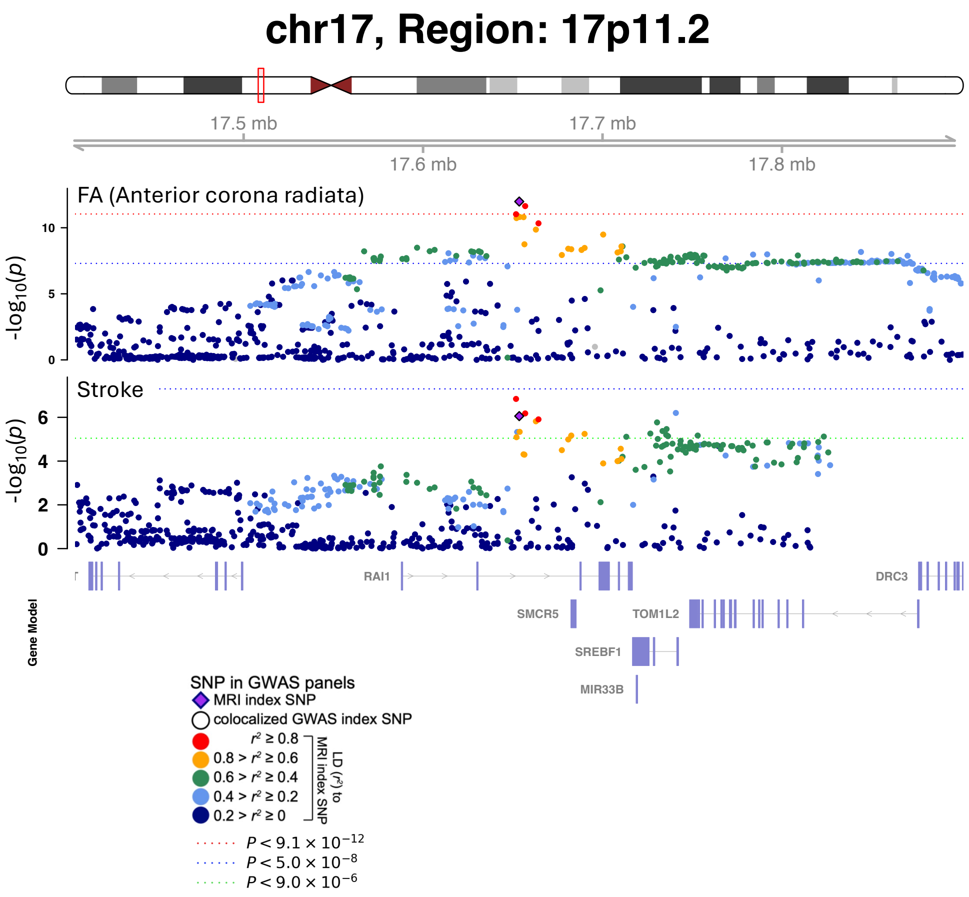

**Supplementary Fig. 32**: **The 17p11.2 white-matter locus shares a causal variant with stroke (PP.H4 = 0.986).** Regional association plots for white-matter microstructure (FA in the anterior corona radiata) and stroke at 17p11.2, with shared index variant rs9900803. Bayesian colocalization supports a single shared causal variant (PP.H4.abf = 0.986).

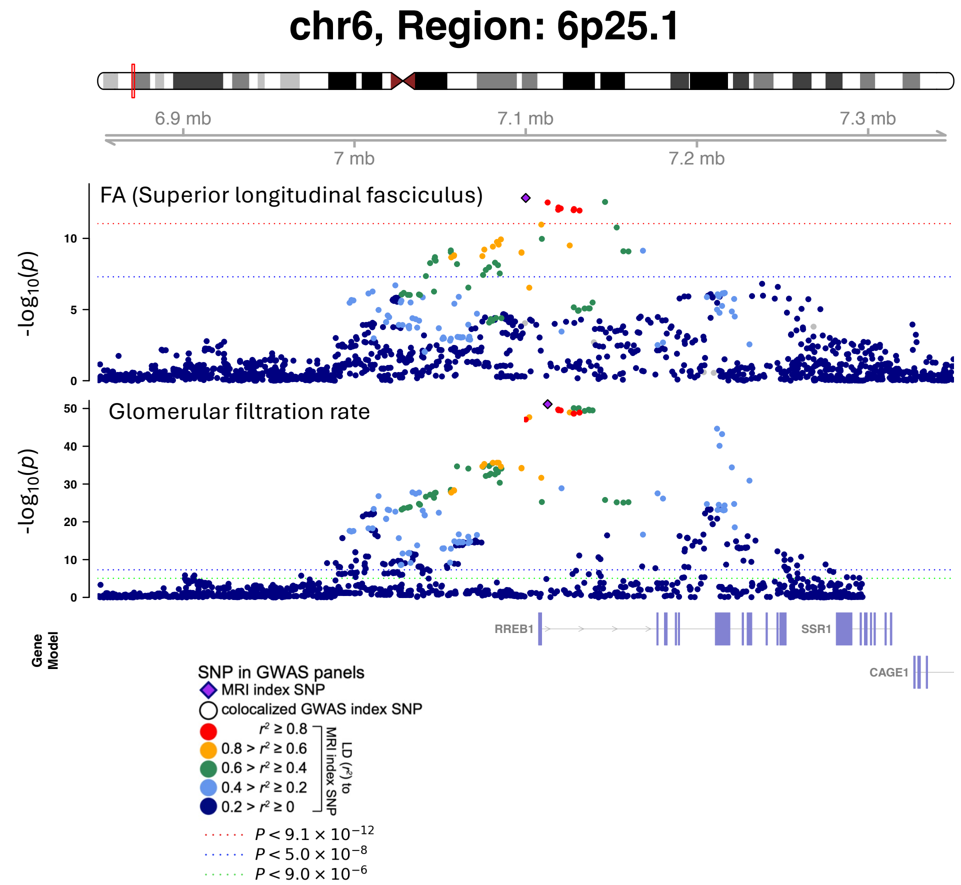

**Supplementary Fig. 33**: **The 6p25.1 white-matter locus shares a causal variant with glomerular filtration rate (PP.H4 = 0.992).** Regional association plots for white-matter microstructure (FA in the superior longitudinal fasciculus, index variant rs12211604) and glomerular filtration rate (index variant rs6925389) at 6p25.1. Bayesian colocalization supports a single shared causal variant (PP.H4.abf = 0.992).

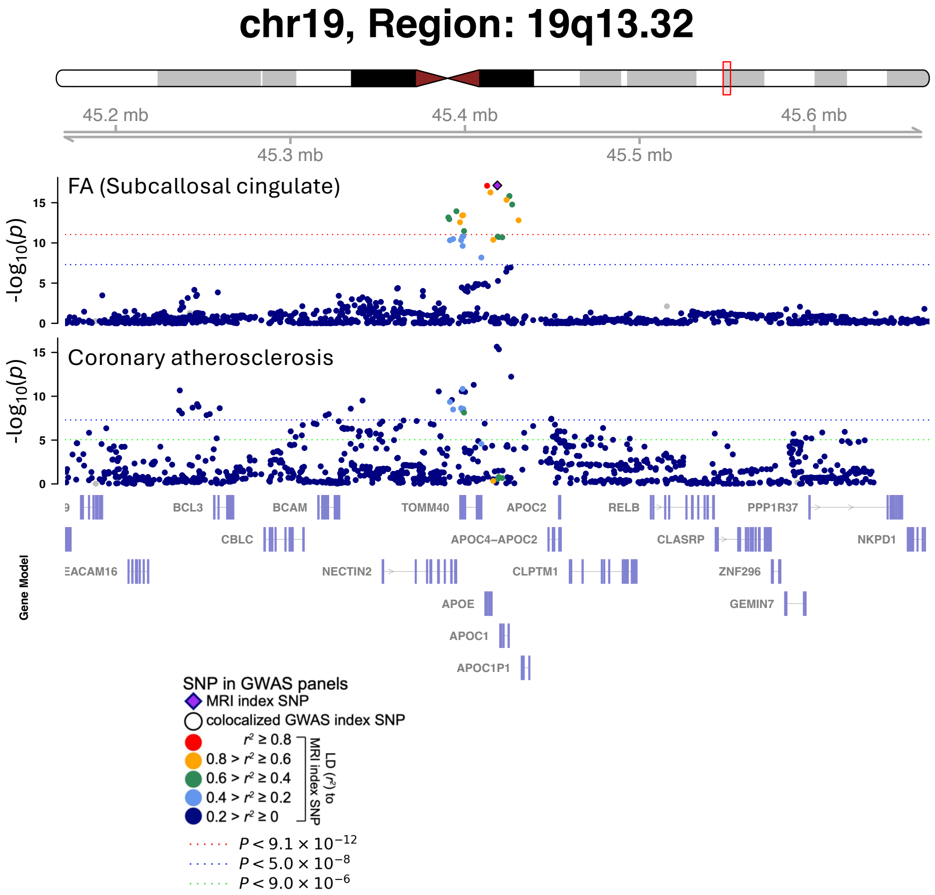

**Supplementary Fig. 34: The 19q13.32 (*APOE/APOC1*) white-matter locus shares a causal variant with coronary atherosclerosis (PP.H4 = 0.995).** Regional association plots for white-matter microstructure (FA in the splenium of the corpus callosum, index variant rs7256200) and coronary atherosclerosis (index variant rs12972970) at 19q13.32. Bayesian colocalization supports a single shared causal variant (PP.H4.abf = 0.995).

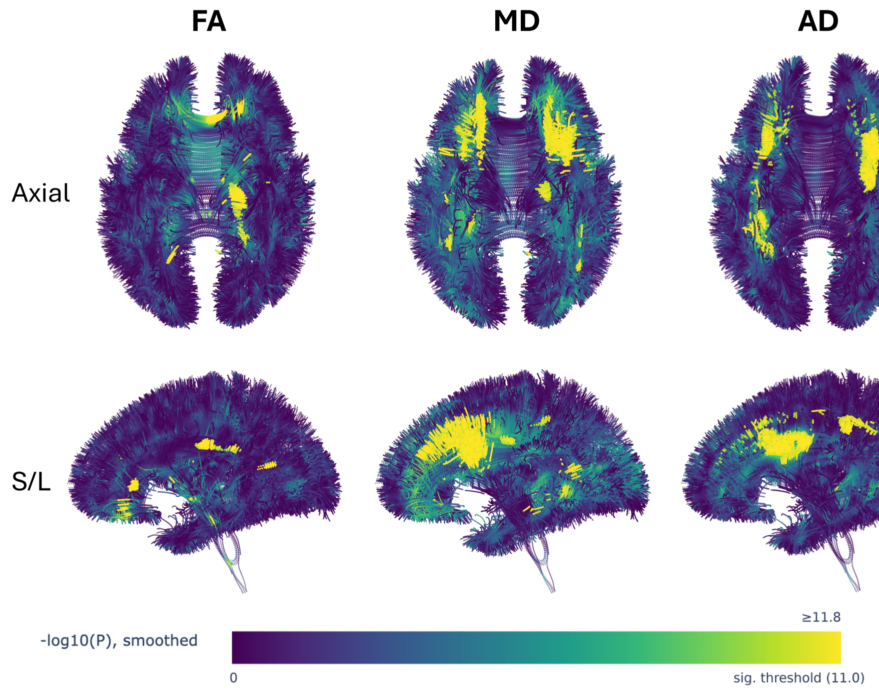

**Supplementary Fig. 35**: **Cross-modality voxel-level association map at rs9262135 (6p21).** FA, MD, and AD voxel-level $-\log_{10}(P)$ maps (smoothed; see Methods) for the FA-discovered, colocalization-prioritized locus at rs9262135, which colocalizes with bipolar disorder (PP.H4= 0.921; Supplementary Table 8). FA associates across 11 JHU tracts (3,145 discovery voxels, $P = 6.51 \times 10^{-19}$); MD and AD are independently genome-wide significant within 250 kb of the FA lead SNP (MD lead rs3130117, 110 kb away, 9 tracts, 13,957 voxels, $P = 4.17 \times 10^{-18}$; AD lead rs9262141, 25 kb away, 12 tracts, 18,426 voxels, $P = 1.03 \times 10^{-20}$). Top row: superior (top-down) view; bottom row: left lateral view. Color scale: smoothed $-\log_{10}(P)$; yellow indicates voxels exceeding the voxel-effective significance threshold ($P < 9.13 \times 10^{-12}$).

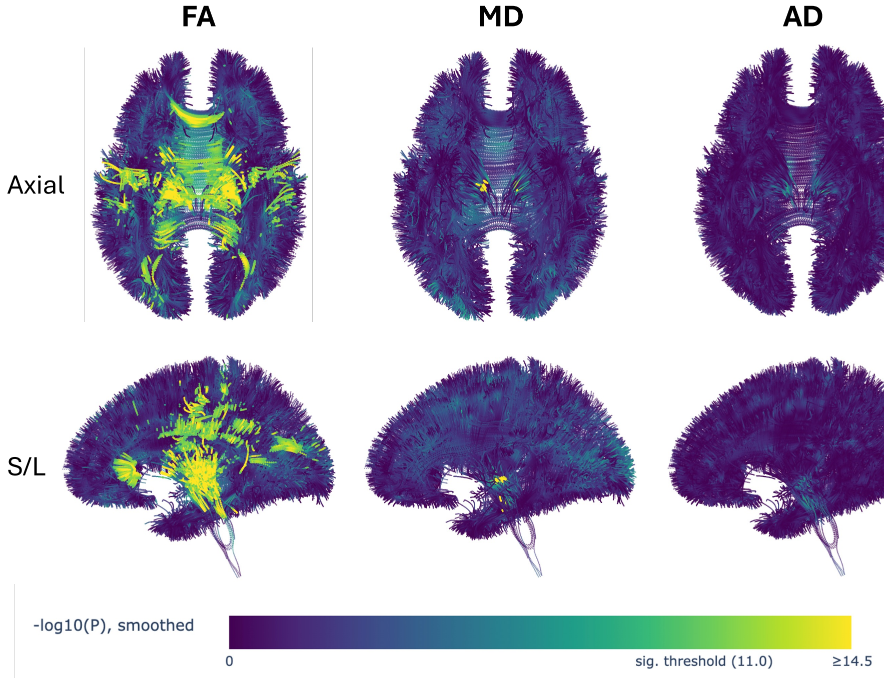

**Supplementary Fig. 36**: **Cross-modality voxel-level association map at rs6062264 (20q13.12).** FA and MD voxel-level $-\log_{10}(P)$ maps (smoothed) for the FA-discovered, colocalization-prioritized locus at rs6062264, which colocalizes with reaction time (PP.H4= 0.912; Supplementary Table 8). FA associates across 13 JHU tracts (12,589 discovery voxels, $P = 8.22 \times 10^{-26}$); MD is independently genome-wide significant 10 kb away (lead rs12479469; 2 tracts, CP/SCR; 105 voxels, $P = 1.48 \times 10^{-13}$), occupying largely distinct voxels from the FA signal (FA–MD Dice = 0.0009; see Section 1.14). AD shows no significant signal within 250 kb of this locus. Top row: superior (top-down) view; bottom row: left lateral view. Color scale as above.

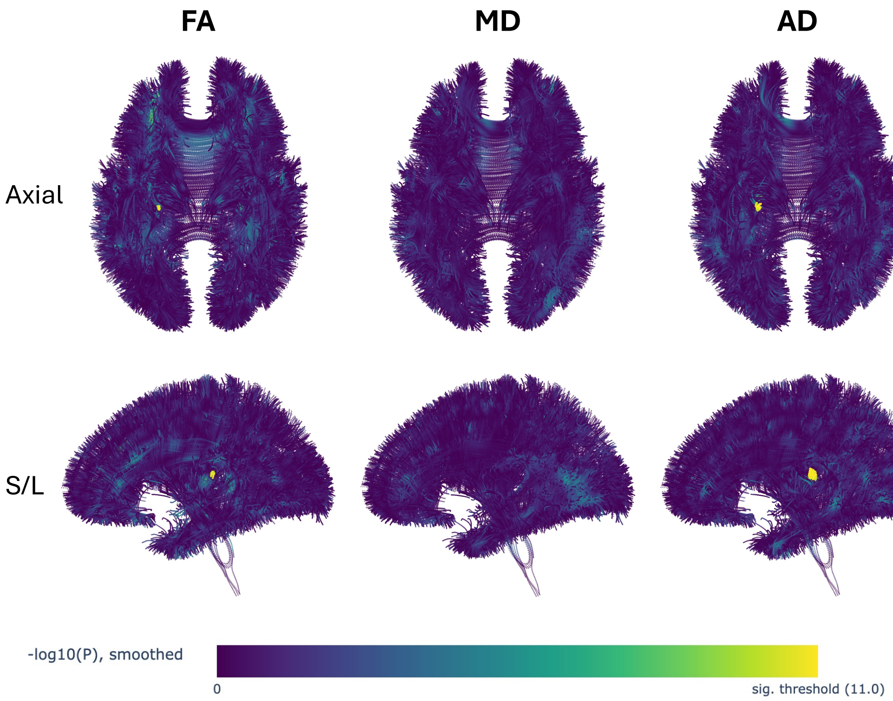

**Supplementary Fig. 37**: **Cross-modality voxel-level association map at rs1747223 (13q14.11).** FA and AD voxel-level $-\log_{10}(P)$ maps (smoothed) for the FA-discovered, colocalization-prioritized locus at rs1747223, which colocalizes with stroke (PP.H4$=$ 0.930; Supplementary Table 8). FA associates across 3 JHU tracts (GCC, PCR, PTR; 51 discovery voxels, $P = 1.04 \times 10^{-12}$); AD is independently genome-wide significant at the same lead SNP position (1 tract, PCR; 117 voxels, $P = 9.27 \times 10^{-16}$). MD shows no significant signal within 250 kb of this locus. Top row: superior (top-down) view; bottom row: left lateral view. Color scale as above.

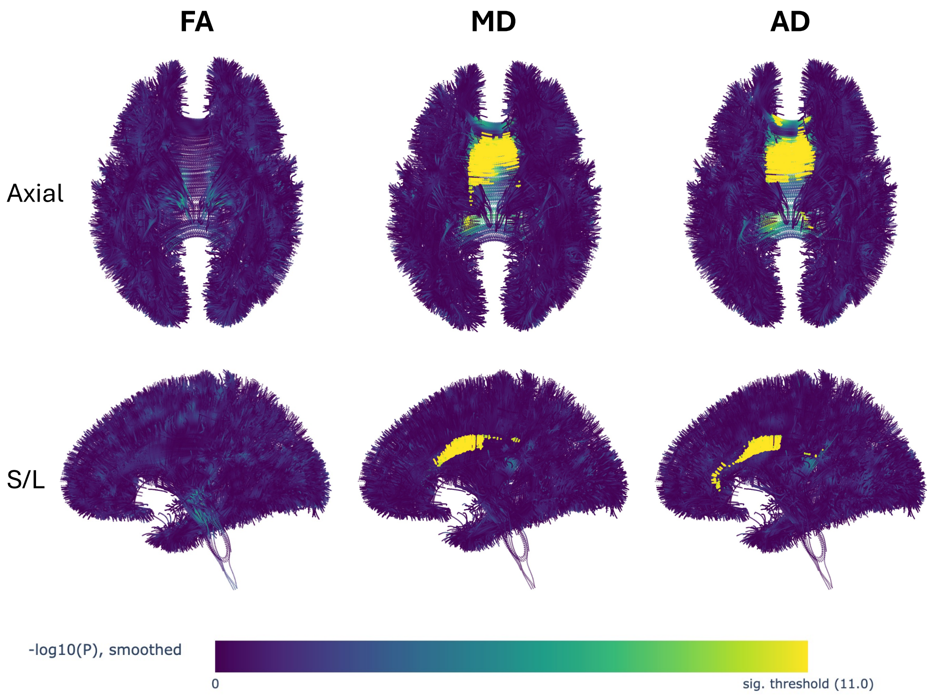

**Supplementary Fig. 38**: **Cross-modality voxel-level association map at rs2306811 (18p11.21).** MD and AD voxel-level $-\log_{10}(P)$ maps (smoothed) at rs2306811. MD (2 tracts, BCC/SCR; 2,339 discovery voxels, $P = 1.97 \times 10^{-19}$) and AD (3 tracts, BCC/GCC/SCR; 2,500 discovery voxels, $P = 4.46 \times 10^{-21}$) are both independently genome-wide significant; FA shows no significant signal within 250 kb of this locus. This locus is not part of the 47-locus, colocalization-prioritized set and has not been evaluated for disease-trait colocalization. Top row: superior (top-down) view; bottom row: left lateral view. Color scale as above.

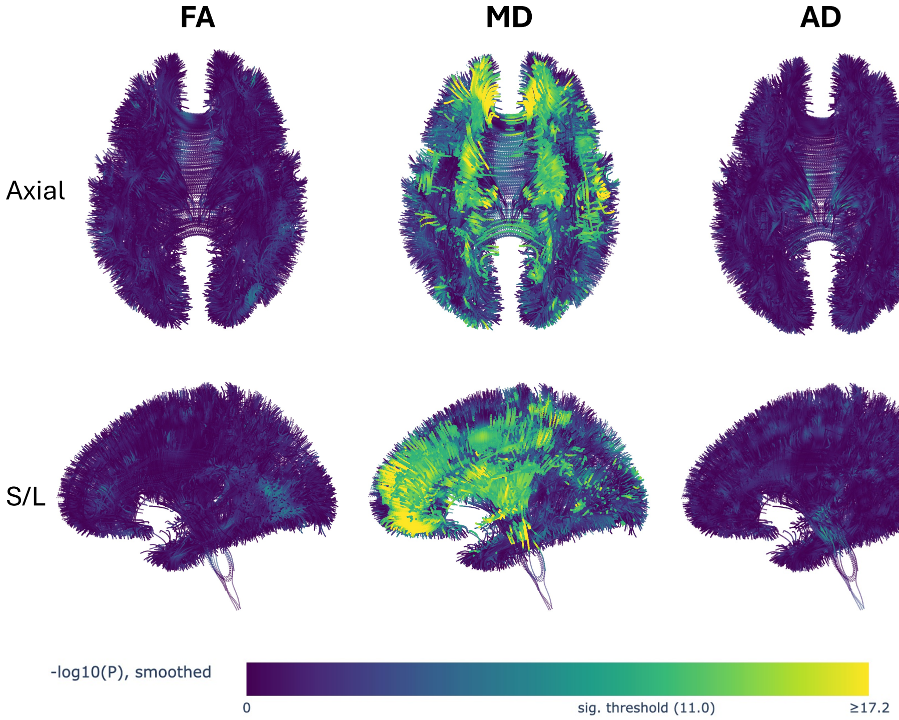

**Supplementary Fig. 39**: **Cross-modality voxel-level association map at rs78903072 (17q21.31).** MD voxel-level $-\log_{10}(P)$ map (smoothed) at rs78903072 (15 tracts, 140,058 discovery voxels, $P = 3.2 \times 10^{-29}$), one of the largest single loci identified in any of the three diffusion atlases. FA and AD show no significant signal within 250 kb of this locus. This locus is not part of the 47-locus, colocalization-prioritized set and has not been evaluated for disease-trait colocalization. Top row: superior (top-down) view; bottom row: left lateral view. Color scale as above.

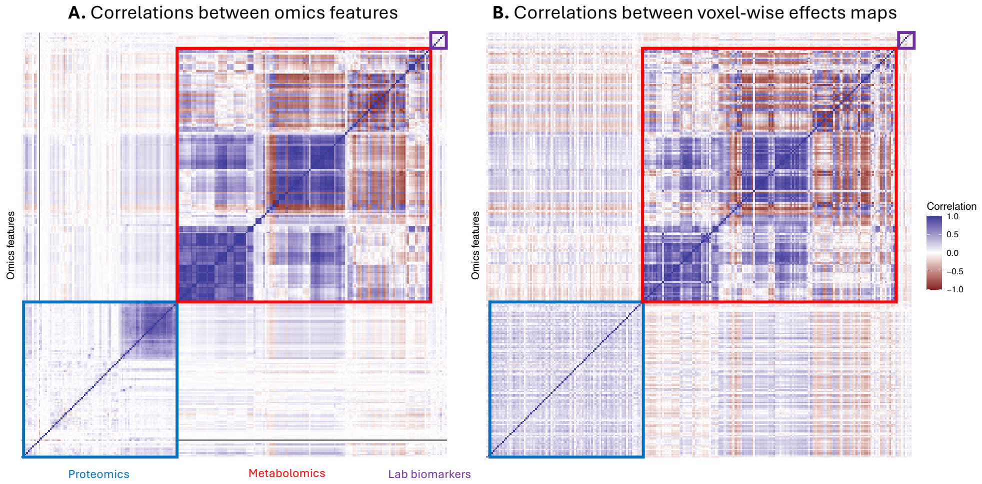

**Supplementary Fig. 40**: **Spatial association maps cluster by omics layer, and spatial concordance dissociates from phenotypic correlation.** Heatmaps of pairwise correlations across proteomic, metabolomic, and clinical biomarker features associated with white-matter microstructure. **(A)** Phenotypic correlations between features measured in the same individuals. **(B)** Correlations between voxel-wise effect maps, reflecting similarity in the spatial patterns of omics associations across fiber pathways; feature pairs with low phenotypic correlation can nonetheless share highly concordant effect maps, and vice versa. Colored annotation boxes group features by omics category (proteomic, metabolomic, biomarker); within each category, features are hierarchically clustered on phenotypic correlations. Color scale: Pearson $r$, $-1$ to 1.

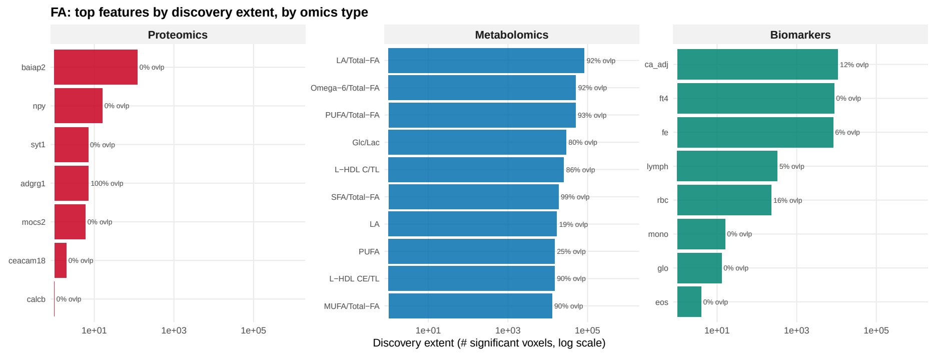

(a) Top discovered features per omics type (proteomics, metabolomics, biomarkers), ranked by validation overlap.

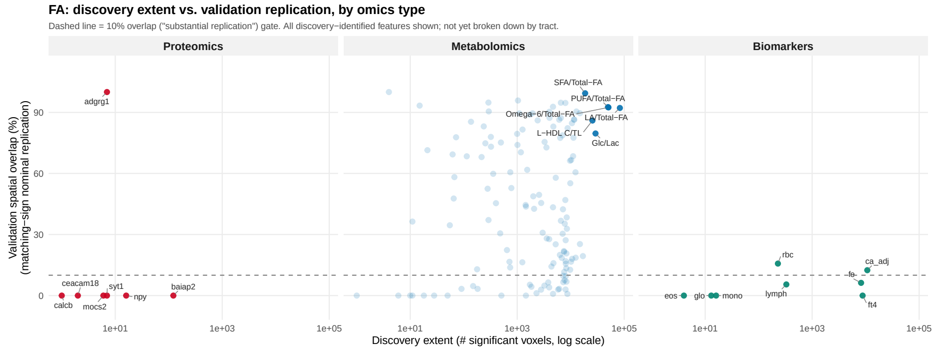

(b) Discovery extent (number of significant voxels at the Bonferroni threshold) versus validation overlap (%), by omics type.

**Supplementary Fig. 41**: Multi-omics top-feature screening and validation-replication summary for fractional anisotropy (FA). Validation replication differed markedly by omics layer: only 1 of 7 proteomic features (14%) and 2 of 8 biomarkers (25%) cleared the 10% discovery-validation overlap threshold, whereas 108 of 137 metabolomic features (79%) did so, led by the linoleic acid, omega-6, and polyunsaturated fatty acid ratios (each >90% voxel-wise overlap between discovery and validation).

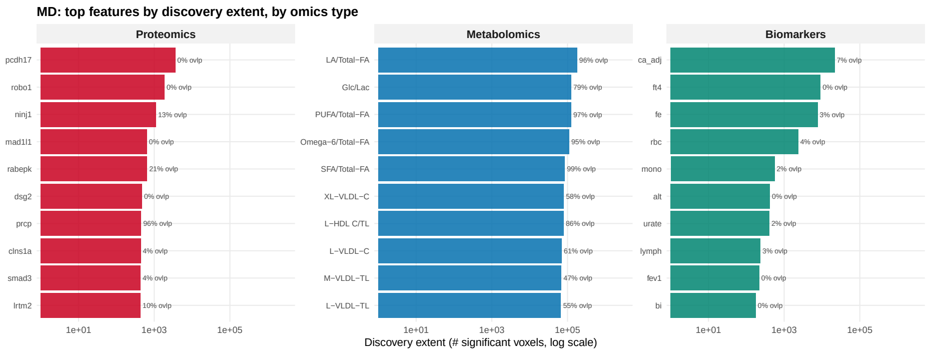

(a) Top discovered features per omics type (proteomics, metabolomics, biomarkers), ranked by validation overlap.

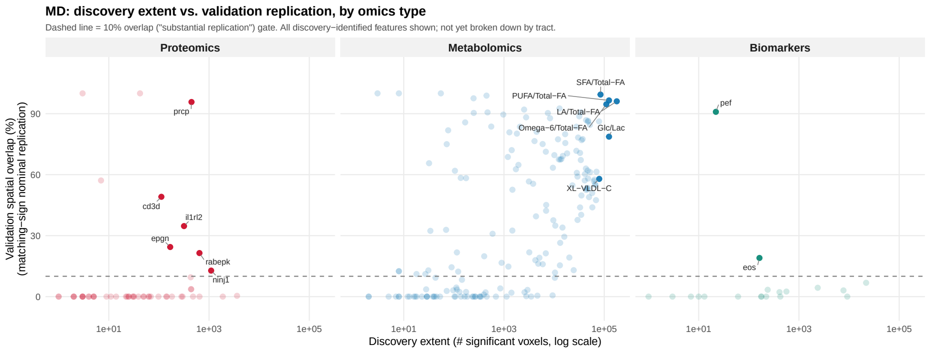

(b) Discovery extent (number of significant voxels at the Bonferroni threshold) versus validation overlap (%), by omics type.

**Supplementary Fig. 42**: Multi-omics top-feature screening and validation-replication summary for mean diffusivity (MD). As with FA, metabolomic features replicated most consistently (124 of 179, 69%), again led by fatty-acid ratio traits (>96% overlap) alongside the glycolysis-related glucose-lactate ratio (79% overlap); proteomic and biomarker replication was sparser overall (9 of 52 proteins, 17%; 2 of 19 biomarkers, 11%), though isolated features such as PRCP (96% overlap) and peak expiratory flow (91% overlap) replicated nearly completely.

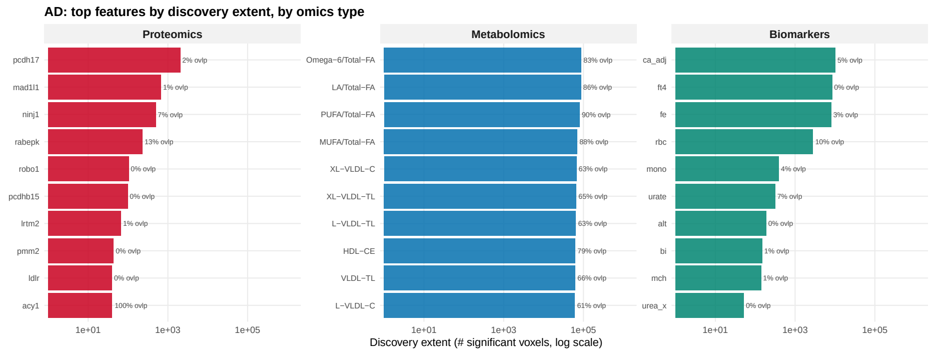

(a) Top discovered features per omics type (proteomics, metabolomics, biomarkers), ranked by validation overlap.

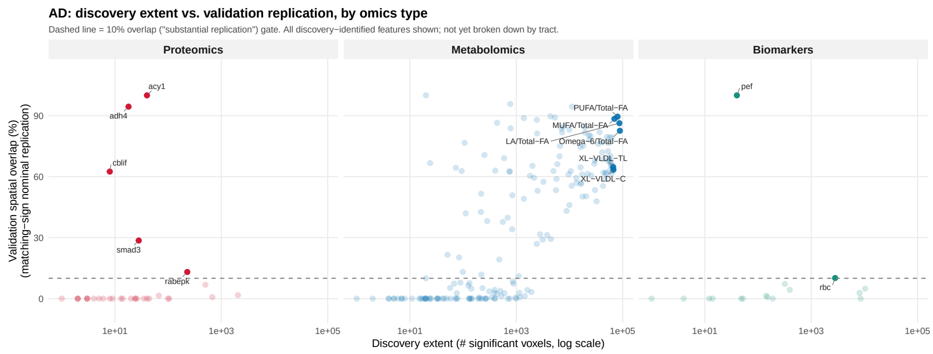

(b) Discovery extent (number of significant voxels at the Bonferroni threshold) versus validation overlap (%), by omics type.

**Supplementary Fig. 43**: Multi-omics top-feature screening and validation-replication summary for axial diffusivity (AD). The same pattern held for AD, with metabolomic features – again dominated by fatty-acid ratio traits – replicating in 112 of 186 cases (60%) at >80% voxel overlap, compared to markedly sparser replication among proteomic (5 of 32, 16%) and biomarker (2 of 16, 13%) features, though isolated proteins (e.g., ACY1, 100% overlap) and biomarkers (peak expiratory flow, 100% overlap) replicated completely.

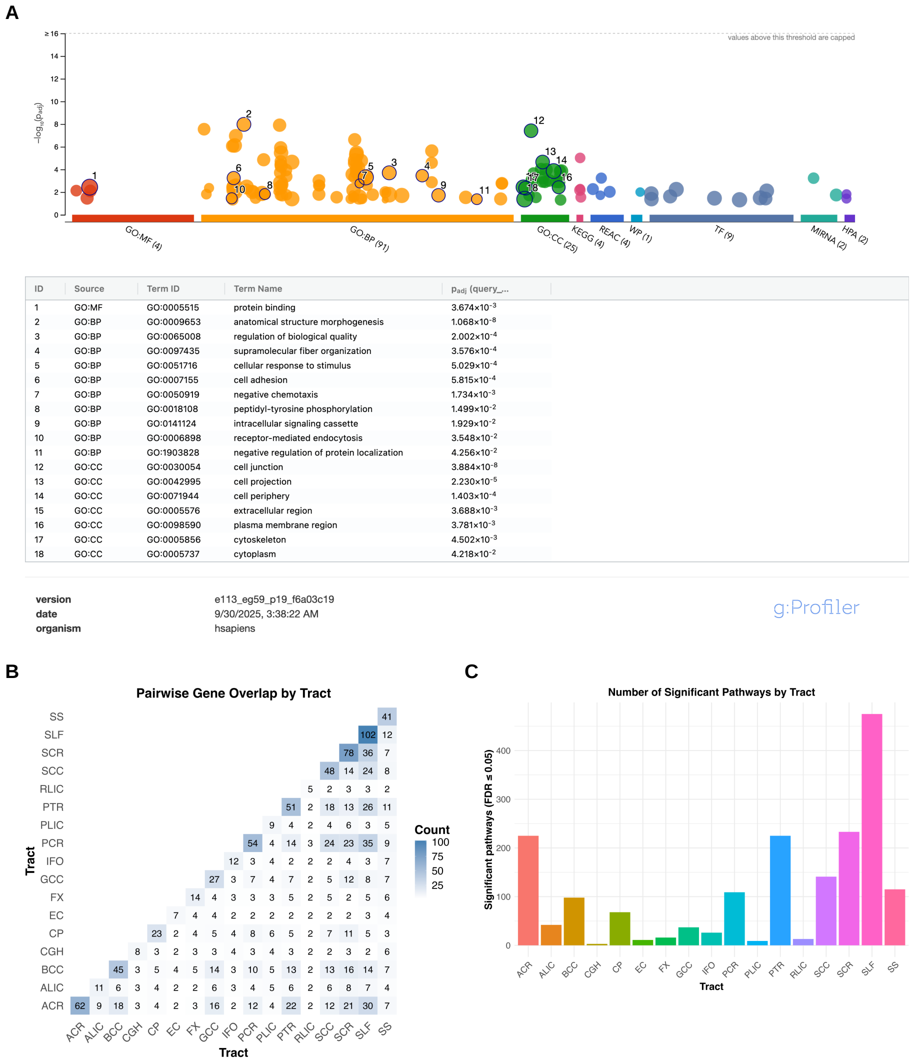

**Supplementary Fig. 44**: **Tract-level gene-set enrichment of the genes at FA loci. (A)** g:Profiler (v0.2.2; GO, KEGG, Reactome and the other databases listed on the panel) enrichment of the genes mapped to FA loci, shown as g:SCS-adjusted $-\log_{10} P$ per term, with the numbered terms listed on the right (protein binding, anatomical-structure morphogenesis, supramolecular-fiber organization, cell adhesion, negative chemotaxis, cell junction and cell projection among them). **(B)** Pairwise overlap of locus genes between the 17 JHU white-matter tracts; the diagonal gives the number of genes assigned to each tract (SLF 102, SCR 78, ACR 62, PCR 54, PTR 51) and off-diagonal cells the number of genes shared by two tracts. **(C)** Number of gene sets significant at FDR $\leq 0.05$ when enrichment is run separately on each tract's gene list; the SLF, ACR, PTR and SCR carry the most enriched pathways, as expected for the tracts that carry the most loci (main Fig. 5a), whereas the CGH, PLIC, EC and RLIC carry few. Per-tract gene lists and enrichment tables are in Supplementary Table 11.

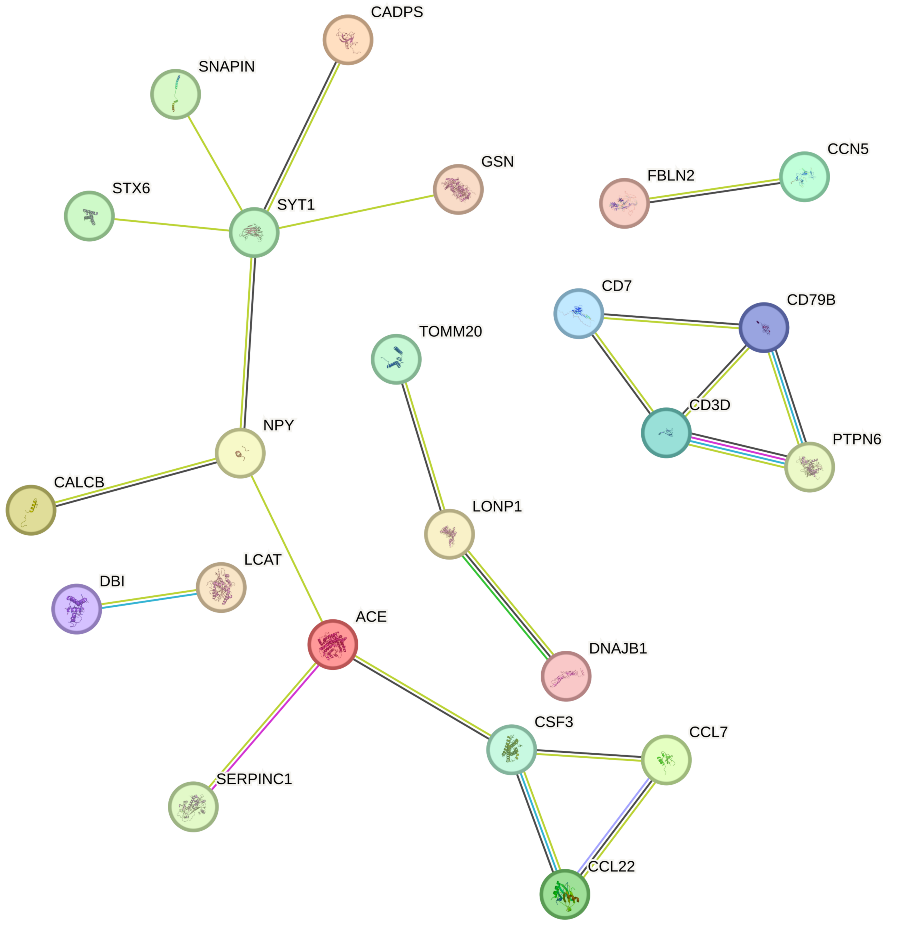

**Supplementary Fig. 45**: **STRING protein–protein interaction network of the FA-associated plasma proteins.** STRING v12.0 network (medium confidence, interaction score > 0.4; edge colours denote evidence channels) for the plasma proteins associated with FA; unconnected proteins are not shown. Connected components include a synaptic-vesicle/neuropeptide module (SYT1 with CADPS, SNAPIN, STX6 and GSN; NPY with CALCB), an ACE-centred module linking SERPINC1, LCAT and the chemokine/colony-stimulating module CSF3–CCL7–CCL22, a T-/B-cell-receptor signalling module (CD3D, CD7, CD79B, PTPN6), a mitochondrial/chaperone chain (TOMM20–LONP1–DNAJB1) and the pairs FBLN2–CCN5 and DBI–LCAT. The network is enriched for extracellular, secreted and receptor-binding proteins (Supplementary Table 11).

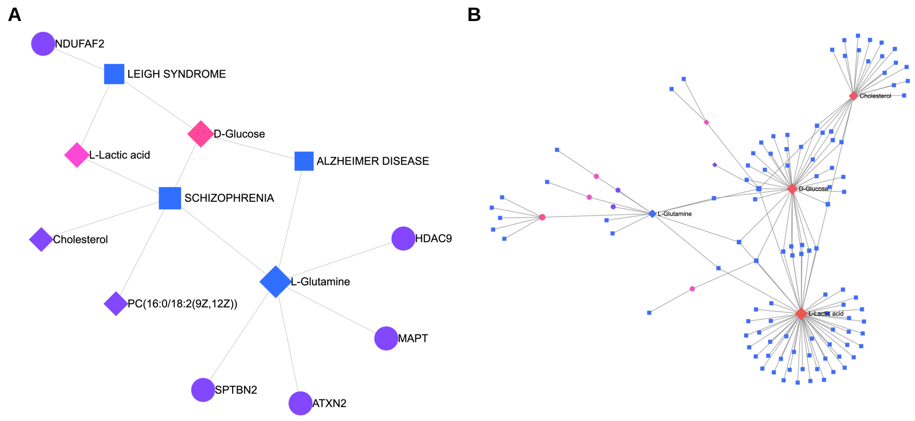

**Supplementary Fig. 46**: **Gene–metabolite–disease network linking the genetic, metabolomic and disease layers. (A)** Highly interconnected subnetwork from the MetaboAnalyst 5.0 Network Analysis module (gene–metabolite relations from HMDB; disease annotations from MetaboAnalyst's ontology; no pathway pre-filtering). Circles are genes (NDUFAF2, HDAC9, MAPT, SPTBN2, ATXN2), diamonds are FA-associated metabolites (L-glutamine, D-glucose, L-lactic acid, cholesterol and total phosphatidylcholines; the Nightingale panel reports total phosphatidylcholines rather than individual species, and the node labelled PC(16:0/18:2) is the HMDB representative to which MetaboAnalyst maps that measure) and squares are diseases (schizophrenia, Alzheimer disease, Leigh syndrome); L-glutamine and D-glucose connect the locus genes to schizophrenia and Alzheimer disease, and L-lactic acid and D-glucose to Leigh syndrome. The Leigh syndrome node arises from the database annotation of the glycolytic metabolites and of the complex-I assembly gene NDUFAF2, and is a database artefact of no relevance to white-matter microstructure. **(B)** The full gene–metabolite–disease network from which the subnetwork in (A) is extracted; the hub metabolites cholesterol, D-glucose, L-lactic acid and L-glutamine (labelled) connect to large sets of disease annotations (squares) and genes (circles). Because disease annotations are curated from the literature, the network summarizes prior knowledge about the implicated genes rather than providing independent evidence of shared mechanism.

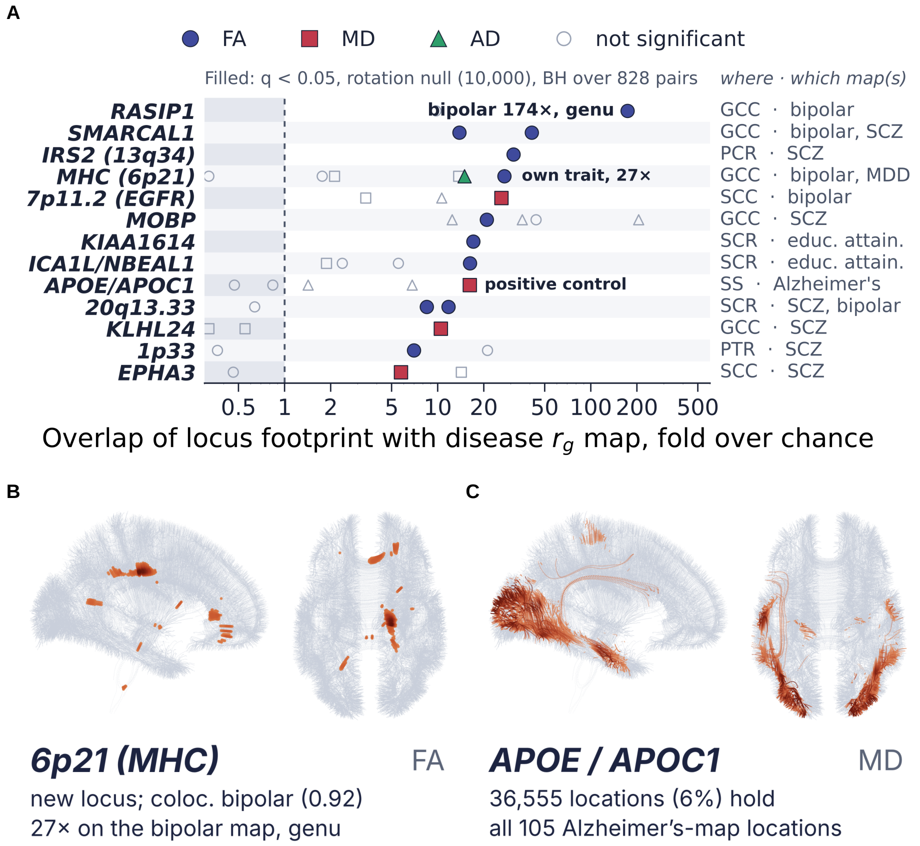

**Supplementary Fig. 47**: **Spatial closure: locus footprints that overlap disease genetic-correlation maps more than expected under a rotation null.** **(A)** For every testable locus × disease-map pair (828 pairs across the 47 colocalized FA loci; locus footprints at $P < 9.12 \times 10^{-12}$, disease $r_g$ maps at $P < 9.12 \times 10^{-6}$), the overlap between footprint and map expressed as fold over chance (log scale). Significance was assessed with a 10,000-draw rotation null (circular shift along each fiber plus fiber permutation within tract; §1.16) and Benjamini–Hochberg correction over all 828 pairs; filled symbols (FA circles, MD squares, AD triangles) mark the 16 pairs with $q < 0.05$, which fall in 13 loci (rows); hollow grey symbols are non-significant pairs; the right-hand column gives the tract in which the overlap lies and the map(s) involved. RASIP1 shows the largest fold (174-fold on the bipolar disorder map, in the genu; 66 of 127 footprint voxels; tied at the null floor, $P = 10^{-4}$, with 11 other pairs); the MHC locus (6p21; rs9262135, colocalized with bipolar disorder, PP.H4 = 0.92) is the only own-trait pair to pass (27-fold on the bipolar disorder map, genu), with the caveat that colocalization in the MHC is unreliable (§2.8); and APOE/APOC1 × Alzheimer's disease (MD) serves as a positive control (the Alzheimer's map includes the APOE region; §2.8). **(B)** FA footprint of the MHC locus (6p21) on the white-matter atlas (sagittal and axial views), concentrated in the genu of the corpus callosum. **(C)** MD footprint of the APOE/APOC1 locus (36,555 voxels, 6% of the skeleton), which contains all 105 voxels of the Alzheimer's disease MD $r_g$ map (16-fold, the maximum attainable for a footprint of this size). All pairs are listed in Supplementary Table 13.

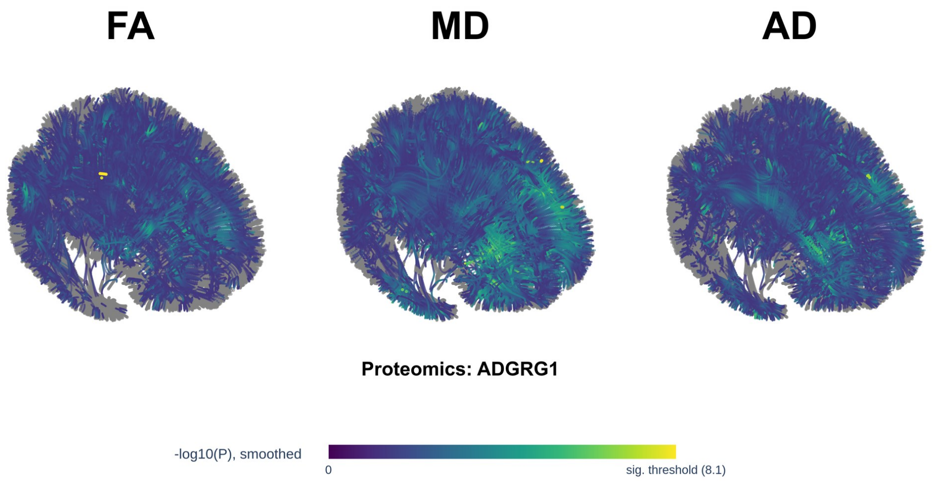

**Supplementary Fig. 48**: Discovery-phase white matter locus atlas for the plasma protein ADGRG1 (adhesion G protein-coupled receptor G1 / GPR56), showing smoothed $-\log_{10}(P)$ association with fractional anisotropy (FA), mean diffusivity (MD), and axial diffusivity (AD) along the white matter fiber atlas. Voxels with nominal $P > 0.05$ are shown in gray; all voxels with nominal $P < 0.05$ are colored by their smoothed $-\log_{10}(P)$ value on a single color scale shared across all three modalities (capped at the 99.5th percentile of these voxels), with the Bonferroni-corrected discovery-phase significance threshold marked as a labeled tick on the colorbar; voxels clearing that threshold are drawn at full opacity, and nominally significant but sub-threshold voxels are drawn dimmed.

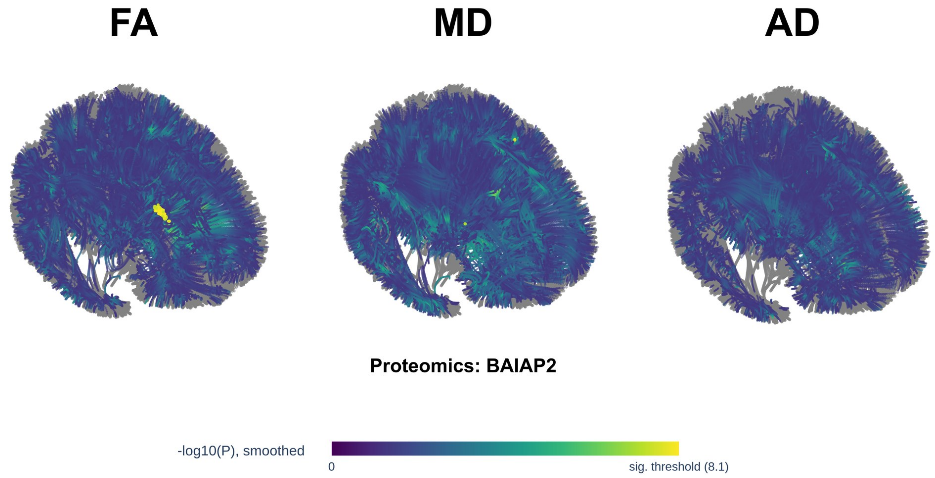

**Supplementary Fig. 49**: Discovery-phase white matter locus atlas for the plasma protein BAIAP2 (BAI1-associated protein 2 / IRSp53), showing smoothed $-\log_{10}(P)$ association with FA, MD, and AD along the white matter fiber atlas. Voxels with nominal $P > 0.05$ are shown in gray; all voxels with nominal $P < 0.05$ are colored by their smoothed $-\log_{10}(P)$ value on a single color scale shared across all three modalities (capped at the 99.5th percentile of these voxels), with the Bonferroni-corrected discovery-phase significance threshold marked as a labeled tick on the colorbar; voxels clearing that threshold are drawn at full opacity, and nominally significant but sub-threshold voxels are drawn dimmed.

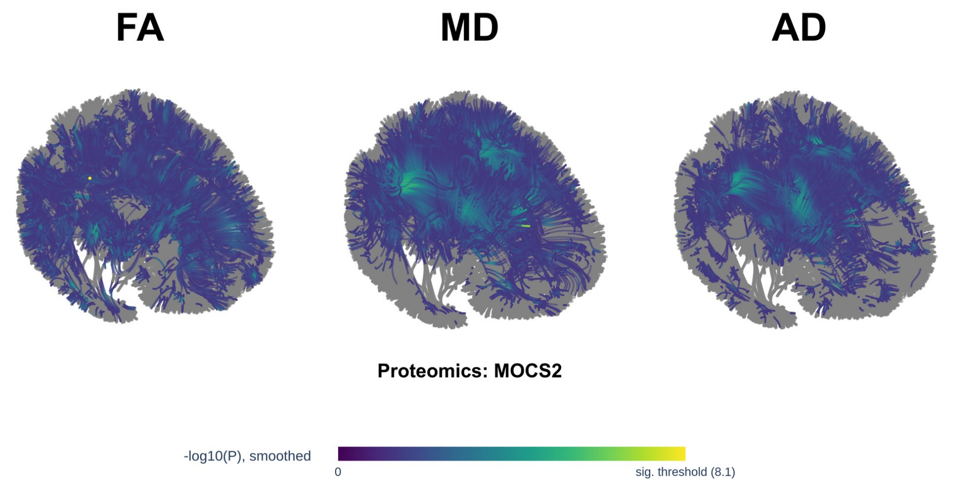

**Supplementary Fig. 50**: Discovery-phase white matter locus atlas for the plasma protein MOCS2 (molybdenum cofactor synthesis 2), showing smoothed $-\log_{10}(P)$ association with FA, MD, and AD along the white matter fiber atlas. Voxels with nominal $P > 0.05$ are shown in gray; all voxels with nominal $P < 0.05$ are colored by their smoothed $-\log_{10}(P)$ value on a single color scale shared across all three modalities (capped at the 99.5th percentile of these voxels), with the Bonferroni-corrected discovery-phase significance threshold marked as a labeled tick on the colorbar; voxels clearing that threshold are drawn at full opacity, and nominally significant but sub-threshold voxels are drawn dimmed.

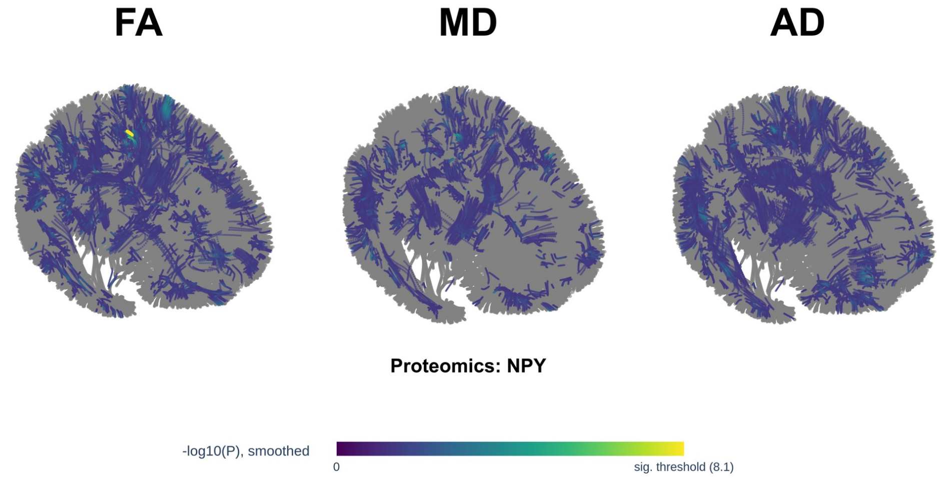

**Supplementary Fig. 51**: Discovery-phase white matter locus atlas for the plasma protein NPY (neuropeptide Y), showing smoothed $-\log_{10}(P)$ association with FA, MD, and AD along the white matter fiber atlas. Voxels with nominal $P > 0.05$ are shown in gray; all voxels with nominal $P < 0.05$ are colored by their smoothed $-\log_{10}(P)$ value on a single color scale shared across all three modalities (capped at the 99.5th percentile of these voxels), with the Bonferroni-corrected discovery- phase significance threshold marked as a labeled tick on the colorbar; voxels clearing that threshold are drawn at full opacity, and nominally significant but sub-threshold voxels are drawn dimmed.

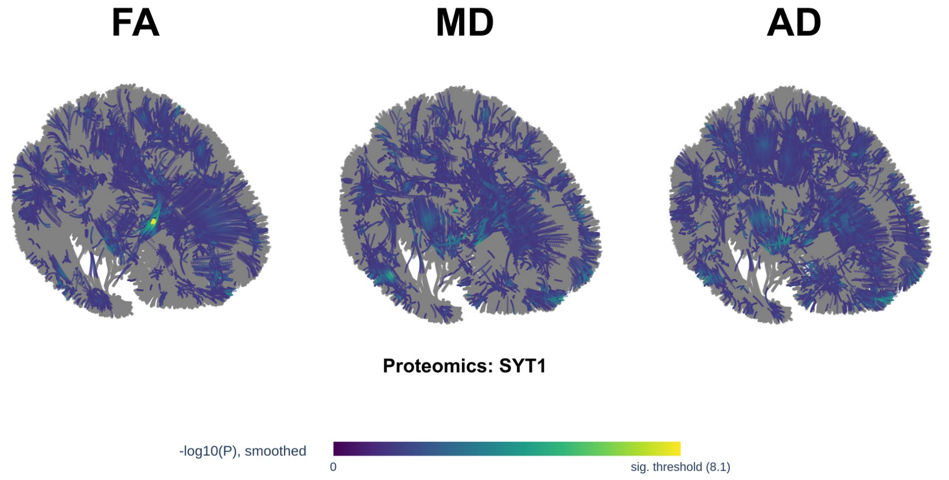

**Supplementary Fig. 52**: Discovery-phase white matter locus atlas for the plasma protein SYT1 (synaptotagmin 1), showing smoothed $-\log_{10}(P)$ association with FA, MD, and AD along the white matter fiber atlas. Voxels with nominal $P > 0.05$ are shown in gray; all voxels with nominal $P < 0.05$ are colored by their smoothed $-\log_{10}(P)$ value on a single color scale shared across all three modalities (capped at the 99.5th percentile of these voxels), with the Bonferroni-corrected discovery- phase significance threshold marked as a labeled tick on the colorbar; voxels clearing that threshold are drawn at full opacity, and nominally significant but sub-threshold voxels are drawn dimmed.

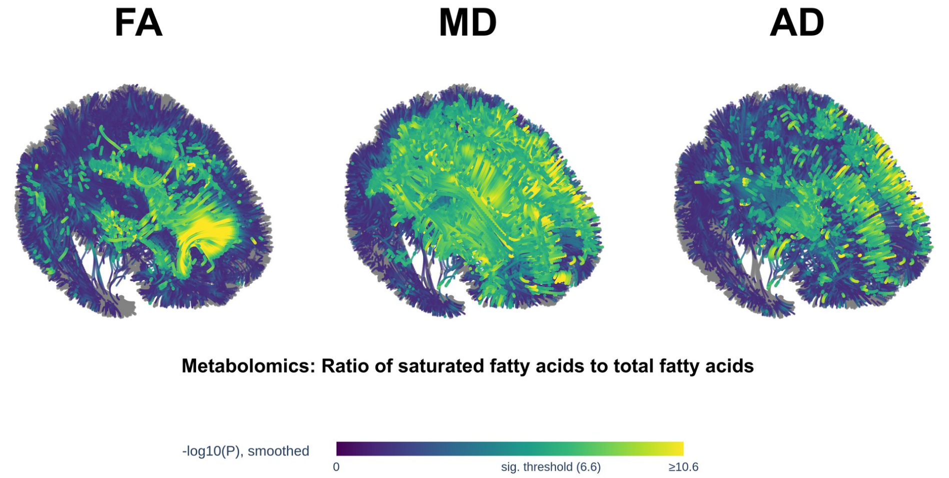

**Supplementary Fig. 53**: Discovery-phase white matter locus atlas for the NMR metabolomics measure *ratio of saturated fatty acids to total fatty acids* (UK Biobank field 23455), showing smoothed $-\log_{10}(P)$ association with FA, MD, and AD along the white matter fiber atlas. Voxels with nominal $P > 0.05$ are shown in gray; all voxels with nominal $P < 0.05$ are colored by their smoothed $-\log_{10}(P)$ value on a single color scale shared across all three modalities (capped at the 99.5th percentile of these voxels), with the Bonferroni-corrected discovery- phase significance threshold marked as a labeled tick on the colorbar; voxels clearing that threshold are drawn at full opacity, and nominally significant but sub-threshold voxels are drawn dimmed.

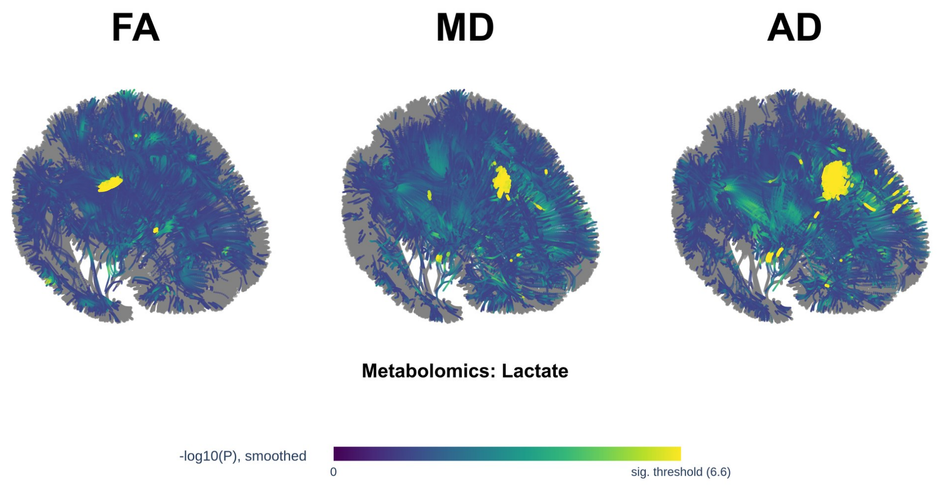

**Supplementary Fig. 54**: Discovery-phase white matter locus atlas for the NMR metabolomics measure *lactate* (UK Biobank field 23471), showing smoothed $-\log_{10}(P)$ association with FA, MD, and AD along the white matter fiber atlas. Voxels with nominal $P > 0.05$ are shown in gray; all voxels with nominal $P < 0.05$ are colored by their smoothed $-\log_{10}(P)$ value on a single color scale shared across all three modalities (capped at the 99.5th percentile of these voxels), with the Bonferroni-corrected discovery-phase significance threshold marked as a labeled tick on the colorbar; voxels clearing that threshold are drawn at full opacity, and nominally significant but sub-threshold voxels are drawn dimmed.

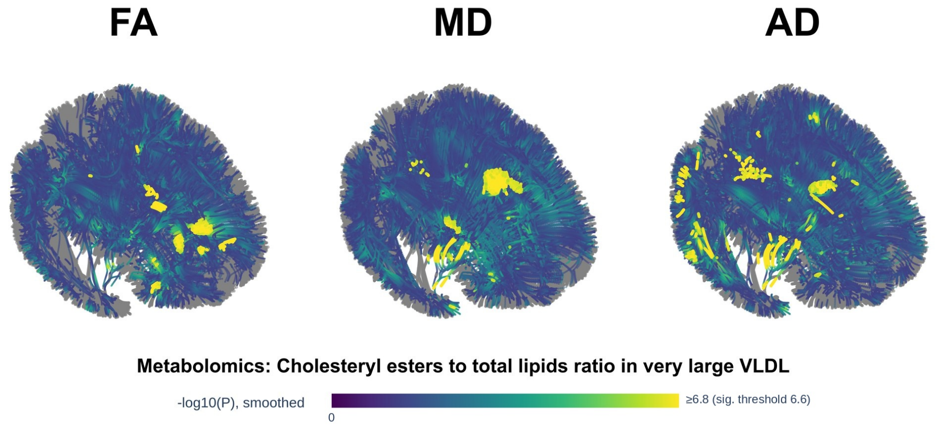

**Supplementary Fig. 55**: Discovery-phase white matter locus atlas for the NMR metabolomics measure *cholesteryl esters to total lipids ratio in very large VLDL* (UK Biobank field 23586), showing smoothed $-\log_{10}(P)$ association with FA, MD, and AD along the white matter fiber atlas. Voxels with nominal $P > 0.05$ are shown in gray; all voxels with nominal $P < 0.05$ are colored by their smoothed $-\log_{10}(P)$ value on a single color scale shared across all three modalities (capped at the 99.5th percentile of these voxels), with the Bonferroni-corrected discovery-phase significance threshold marked as a labeled tick on the colorbar; voxels clearing that threshold are drawn at full opacity, and nominally significant but sub-threshold voxels are drawn dimmed.

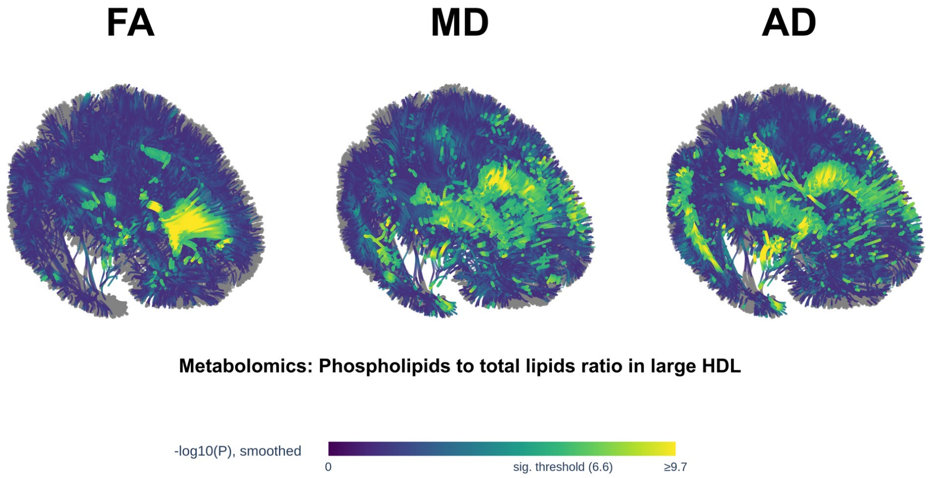

**Supplementary Fig. 56**: Discovery-phase white matter locus atlas for the NMR metabolomics measure *phospholipids to total lipids ratio in large HDL* (UK Biobank field 23634), showing smoothed $-\log_{10}(P)$ association with FA, MD, and AD along the white matter fiber atlas. Voxels with nominal $P > 0.05$ are shown in gray; all voxels with nominal $P < 0.05$ are colored by their smoothed $-\log_{10}(P)$ value on a single color scale shared across all three modalities (capped at the 99.5th percentile of these voxels), with the Bonferroni-corrected discovery- phase significance threshold marked as a labeled tick on the colorbar; voxels clearing that threshold are drawn at full opacity, and nominally significant but sub-threshold voxels are drawn dimmed.

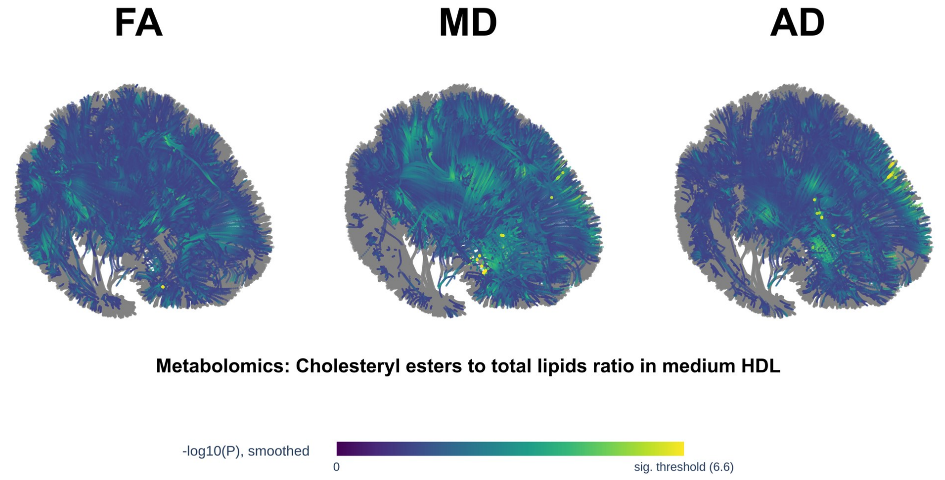

**Supplementary Fig. 57**: Discovery-phase white matter locus atlas for the NMR metabolomics measure *cholesteryl esters to total lipids ratio in medium HDL* (UK Biobank field 23641), showing smoothed $-\log_{10}(P)$ association with FA, MD, and AD along the white matter fiber atlas. Voxels with nominal $P > 0.05$ are shown in gray; all voxels with nominal $P < 0.05$ are colored by their smoothed $-\log_{10}(P)$ value on a single color scale shared across all three modalities (capped at the 99.5th percentile of these voxels), with the Bonferroni-corrected discovery- phase significance threshold marked as a labeled tick on the colorbar; voxels clearing that threshold are drawn at full opacity, and nominally significant but sub-threshold voxels are drawn dimmed.

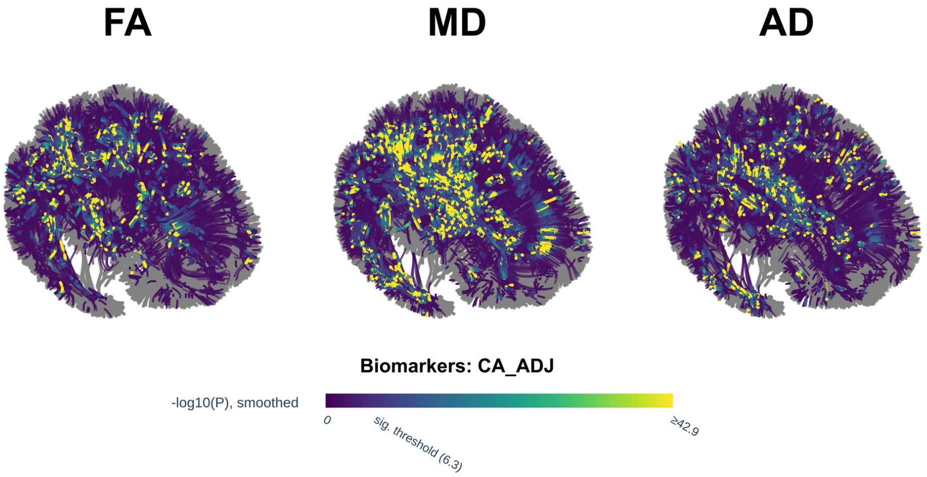

**Supplementary Fig. 58**: Discovery-phase white matter locus atlas for albumin-adjusted calcium (UK Biobank biomarker `ca_adj`), showing smoothed $-\log_{10}(P)$ association with FA, MD, and AD along the white matter fiber atlas. Voxels with nominal $P > 0.05$ are shown in gray; all voxels with nominal $P < 0.05$ are colored by their smoothed $-\log_{10}(P)$ value on a single color scale shared across all three modalities (capped at the 99.5th percentile of these voxels), with the Bonferroni-corrected discovery-phase significance threshold marked as a labeled tick on the colorbar; voxels clearing that threshold are drawn at full opacity, and nominally significant but sub-threshold voxels are drawn dimmed.

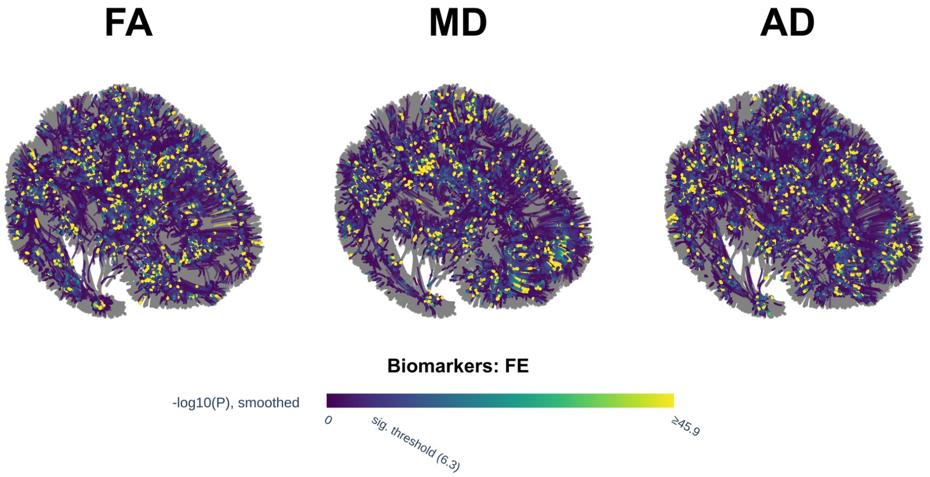

**Supplementary Fig. 59**: Discovery-phase white matter locus atlas for serum iron (UK Biobank biomarker `fe`), showing smoothed $-\log_{10}(P)$ association with FA, MD, and AD along the white matter fiber atlas. Voxels with nominal $P > 0.05$ are shown in gray; all voxels with nominal $P < 0.05$ are colored by their smoothed $-\log_{10}(P)$ value on a single color scale shared across all three modalities (capped at the 99.5th percentile of these voxels), with the Bonferroni-corrected discovery- phase significance threshold marked as a labeled tick on the colorbar; voxels clearing that threshold are drawn at full opacity, and nominally significant but sub-threshold voxels are drawn dimmed.

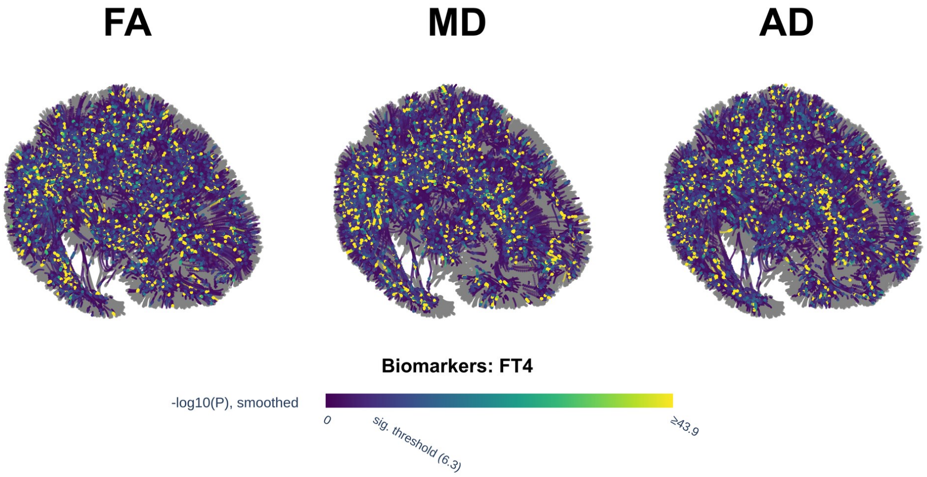

**Supplementary Fig. 60**: Discovery-phase white matter locus atlas for free thyroxine (FT4; UK Biobank biomarker `ft4`), showing smoothed $-\log_{10}(P)$ association with FA, MD, and AD along the white matter fiber atlas. Voxels with nominal $P > 0.05$ are shown in gray; all voxels with nominal $P < 0.05$ are colored by their smoothed $-\log_{10}(P)$ value on a single color scale shared across all three modalities (capped at the 99.5th percentile of these voxels), with the Bonferroni-corrected discovery-phase significance threshold marked as a labeled tick on the colorbar; voxels clearing that threshold are drawn at full opacity, and nominally significant but sub-threshold voxels are drawn dimmed.

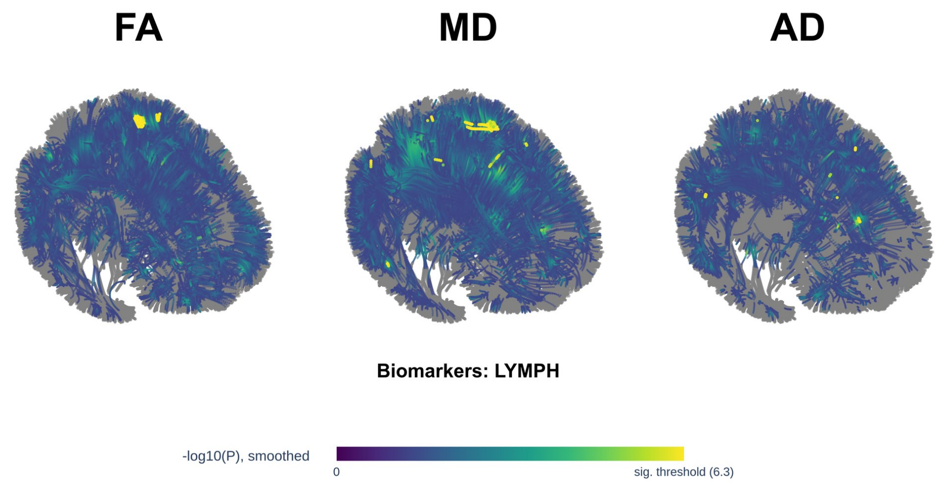

**Supplementary Fig. 61**: Discovery-phase white matter locus atlas for lymphocyte count (UK Biobank biomarker `lymph`), showing smoothed $-\log_{10}(P)$ association with FA, MD, and AD along the white matter fiber atlas. Voxels with nominal $P > 0.05$ are shown in gray; all voxels with nominal $P < 0.05$ are colored by their smoothed $-\log_{10}(P)$ value on a single color scale shared across all three modalities (capped at the 99.5th percentile of these voxels), with the Bonferroni-corrected discovery-phase significance threshold marked as a labeled tick on the colorbar; voxels clearing that threshold are drawn at full opacity, and nominally significant but sub-threshold voxels are drawn dimmed.

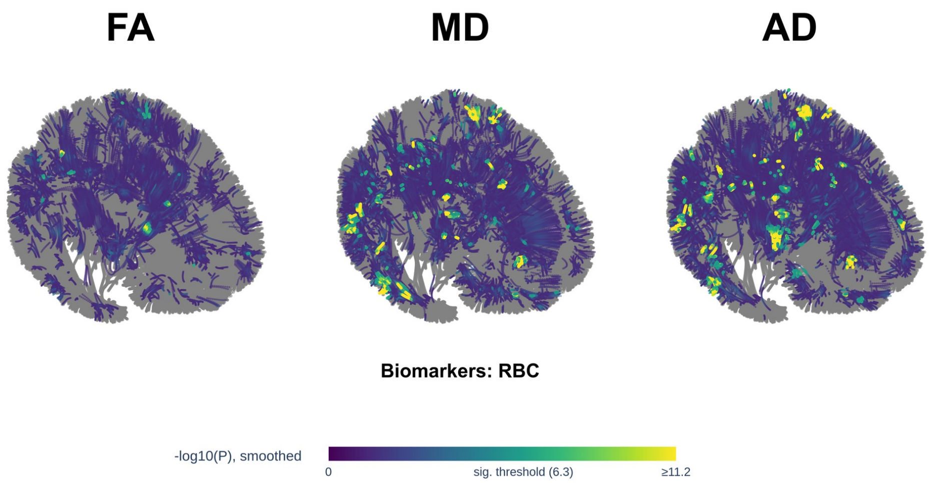

**Supplementary Fig. 62**: Discovery-phase white matter locus atlas for red blood cell (erythrocyte) count (UK Biobank biomarker `rbc`), showing smoothed $-\log_{10}(P)$ association with FA, MD, and AD along the white matter fiber atlas. Voxels with nominal $P > 0.05$ are shown in gray; all voxels with nominal $P < 0.05$ are colored by their smoothed $-\log_{10}(P)$ value on a single color scale shared across all three modalities (capped at the 99.5th percentile of these voxels), with the Bonferroni-corrected discovery-phase significance threshold marked as a labeled tick on the colorbar; voxels clearing that threshold are drawn at full opacity, and nominally significant but sub-threshold voxels are drawn dimmed.
